# Multi-ancestry gene-trait connection landscape using electronic health record (EHR) linked biobank data

**DOI:** 10.1101/2021.10.21.21265225

**Authors:** Binglan Li, Yogasudha Veturi, Anastasia Lucas, Yuki Bradford, Shefali S. Verma, Anurag Verma, Joseph Park, Wei-Qi Wei, Qiping Feng, Bahram Namjou, Krzysztof Kiryluk, Iftikhar Kullo, Yuan Luo, Milton Pividori, Hae Kyung Im, Casey S. Greene, Marylyn D. Ritchie

## Abstract

Understanding genetic factors of complex traits across ancestry groups holds a key to improve the overall health care quality for diverse populations in the United States. In recent years, multiple electronic health record-linked (EHR-linked) biobanks have recruited participants of diverse ancestry backgrounds; these biobanks make it possible to obtain phenome-wide association study (PheWAS) summary statistics on a genome-wide scale for different ancestry groups. Moreover, advancement in bioinformatics methods provide novel means to accelerate the translation of basic discoveries to clinical utility by integrating GWAS summary statistics and expression quantitative trait locus (eQTL) data to identify complex trait-related genes, such as transcriptome-wide association study (TWAS) and colocalization analyses. Here, we combined the advantages of multi-ancestry biobanks and data integrative approaches to investigate the multi-ancestry, gene-disease connection landscape. We first performed a phenome-wide TWAS on Electronic Medical Records and Genomics (eMERGE) III network participants of European ancestry (N = 68,813) and participants of African ancestry (N = 12,658) populations, separately. For each ancestry group, the phenome-wide TWAS tested gene-disease associations between 22,535 genes and 309 curated disease phenotypes in 49 primary human tissues, as well as cross-tissue associations. Next, we identified gene-disease associations that were shared across the two ancestry groups by combining the ancestry-specific results via meta-analyses. We further applied a Bayesian colocalization method, fastENLOC, to prioritize likely functional gene-disease associations with supportive colocalized eQTL and GWAS signals. We replicated the phenome-wide gene-disease analysis in the analogous Penn Medicine BioBank (PMBB) cohorts and sought additional validations in the PhenomeXcan UK Biobank (UKBB) database, PheWAS catalog, and systematic literature review. Phenome-wide TWAS identified many proof-of-concept gene-disease associations, e.g. *FTO*-obesity association (p = 7.29e-15), and numerous novel disease-associated genes, e.g. association between *GATA6-AS1* with pulmonary heart disease (p = 4.60e-10). In short, the multi-ancestry, gene-disease connection landscape provides rich resources for future multi-ancestry complex disease research. We also highlight the importance of expanding the size of non-European ancestry datasets and the potential of exploring ancestry-specific genetic analyses as these will be critical to improve our understanding of the genetic architecture of complex disease.

## Introduction

Human genomics research has achieved great progress in identifying complex disease associated genetic variants via genome-wide association studies (GWAS) and achieving a better understanding of the genetic architecture of many complex human diseases. However, the majority of complex disease genetic studies focus on populations descending from Europe [1-3]. In addition, translating GWAS discoveries to clinical care has been difficult due to the obstacles in mapping genetic variants to their downstream affected genes.

Under-representation of non-European ancestry populations in complex disease research is well known and can potentially exacerbate the disparity in health outcomes [4]. Non-European ancestry populations, especially those of African ancestry, carry variants private to their own ancestry subpopulation [5]. These private variants likely contribute to disease susceptibility in an ancestry-specific way. On the other hand, understanding of these private variants may unravel general biological insights of complex diseases and thus, improve clinical care beyond only the ancestry-defined subpopulations in which they are identified [6,7]. Inclusion of individuals from diverse ancestry groups will enrich and expand the discoveries of complex disease susceptibility genetic factors. A survey of the NHGRI-EBI GWAS Catalog [8], one of the most comprehensive data repositories of GWAS data, found that while African ancestry individuals only comprised 2.4% of the overall study participants, they contributed approximately three times as many genetic associations (7%) as the expected proportions [1]. Still, it is unclear how many yet undiscovered genetic associations are ancestry-specific or shared across European and African ancestry populations; and how these may vary across a variety of complex human diseases and traits.

Electronic health record (EHR) linked biobanks provide opportunities to perform phenome-wide studies for European and African ancestry populations. EHR-linked biobanks, such as Penn Medicine BioBank (PMBB) [9] and the Electronic Medical Records and Genomics (eMERGE) network phase III (hereafter referred to as eMERGE III) [10], recruit individuals of diverse ancestry backgrounds from their respective health systems to participate in the institutional EHR-linked biobanks. These biobanks include access to the EHR for the participants along with DNA samples, which can be used for GWAS and other molecular studies. Included in the EHR data is a documentation of each participant’s clinical visits over time using disease diagnosis codes. These diagnosis codes, typically ICD-9 or ICD-10 codes (International Classification of Disease, version 9 or 10), are used to document diagnoses at each visit and to bill insurance providers. These disease diagnosis codes capture an individual’s interaction with the health care system and can be used to depict health conditions and diseases in retrospect. In summary, eMERGE III and PMBB datasets each provide a wide range of diseases and traits (also known as phenotypes) through the use of EHR data that provides an opportunity to conduct a phenome-wide study across various complex diseases. Due to the ancestral diversity of the patient-participants recruited into the PMBB and into the multiple EHR-linked biobanks included in eMERGE III, this phenome-wide study can be conducted in both European ancestry and African ancestry populations.

The identification of a greater number of disease-associated genes has been enabled by the growing quantity of functional genomics data and data integrative bioinformatics algorithms. The Genotype-Tissue Expression (GTEx) consortium released the v8 reference transcriptome data for 49 primary human tissues or cell lines from samples of multiple ancestry groups, including individuals of European ancestry (85.3%) and African ancestry (12.3%) [11]. Transcriptome-wide association studies (TWAS) and colocalization analyses provide complementary means to identify transcriptionally regulated genes that lead to differential disease risks among individuals. TWAS integrates expression quantitative trait locus (eQTL) information with either individual-level genotype data or GWAS summary statistics to estimate if a gene is transcriptionally regulated by one or multiple eQTLs to result in a certain complex disease [12-15]. Alternatively, colocalization analyses evaluate if a GWAS signal alters the risk of a certain disease by influencing gene expression levels through statistical probabilistic models.

Taking advantage of multi-ancestry EHR-linked biobanks through eMERGE III and PMBB, we investigated ancestry-specific and cross-ancestry complex disease associated genes on a phenome-wide scale for European ancestry and African ancestry populations. Here, based on our previous work [16], we designed a GWAS summary statistics-based phenome-wide TWAS framework that performed a combination of tissue-specific and cross-tissue TWAS to comprehensively capture gene-disease connections. We first applied phenome-wide TWAS to the eMERGE III individuals of European ancestry (N = 68,813) and to individuals of African ancestry (N = 12,658), separately, to investigate ancestry-specific gene-disease associations. Then, meta-analyses combined ancestry-specific GWAS summary statistics to allow TWAS inspection of cross-ancestry gene-disease associations. Moreover, we applied fastENLOC [56], a computationally efficient Bayesian colocalization algorithm, to prioritize functional gene-disease associations that were further supported by colocalized eQTL and GWAS signals. Subsequently, we attempted to replicate the eMERGE III statistically significant gene-disease associations in the PMBB European ancestry population (N = 6,967) and African ancestry population (N = 8,506), respectively. We also sought additional validation against centralized genetics databases including PhenomeXcan [17], PheWAS catalog [18], GWAS catalog [8], and through systematic literature review. Overall, this study identified and replicated ancestry-specific and cross-ancestry gene-disease associations for both European ancestry and African ancestry populations on a phenome-wide scale and provided resources for future multi-ancestry studies.

## Methods and materials

### Design and overview of this study

In order to depict a multi-ancestry, gene-disease connection landscape, we designed and implemented a high-throughput phenome-wide TWAS framework based on our previous evaluation of multiple representative TWAS approaches [16] (**Figure 1**). As individual-level data phenome-wide TWAS takes a substantial amount of computing resources, so much that the analyses would be nearly impossible to finish, our phenome-wide TWAS framework used GWAS summary statistics as input. We first conducted genome-wide phenome-wide association studies (PheWAS) using Regenie [19]. Regenie is an efficient GWAS tool that adopts linear mixed models; and can properly account for population structure and unbalanced case/control ratios commonly seen in EHR-based phenome-wide studies. Regenie produced phenome-wide GWAS summary statistics between 309 phecode-derived case/control-based disease phenotypes (**Supplementary Table 1**) and ∼6M single nucleotide polymorphism (SNP) loci for the eMERGE III individuals of European ancestry (N = 67,885), and ∼11M SNPs for the eMERGE III individuals of African ancestry (N = 12,658), separately. The eMERGE III individuals of African ancestry had similar case/control ratios in comparison to the individuals of European ancestry for the majority of phenotypes, but the African ancestry subset had lower sample sizes overall (**Figures 2 and 3**). The eMERGE III subset of individuals of African ancestry had much lower power and detected only a few genome-wide significant GWAS signals (p < 5e-8) likely due to the limited sample size (described in **Results** and depicted in **Figures 4 and 5**).

**Figure 1.**
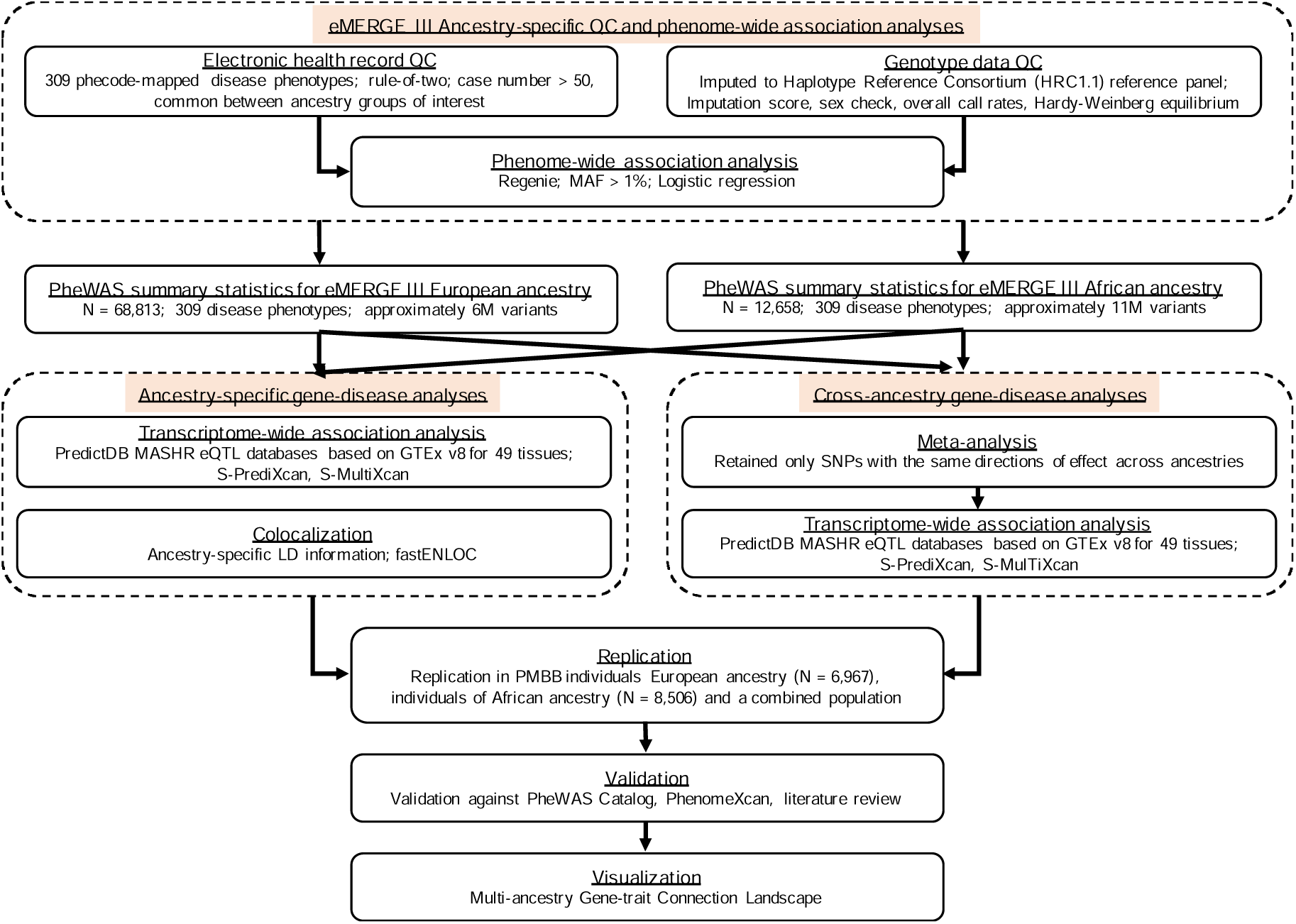
Analytic framework for multi-ancestry gene-disease connection landscape.

**Figure 2.**
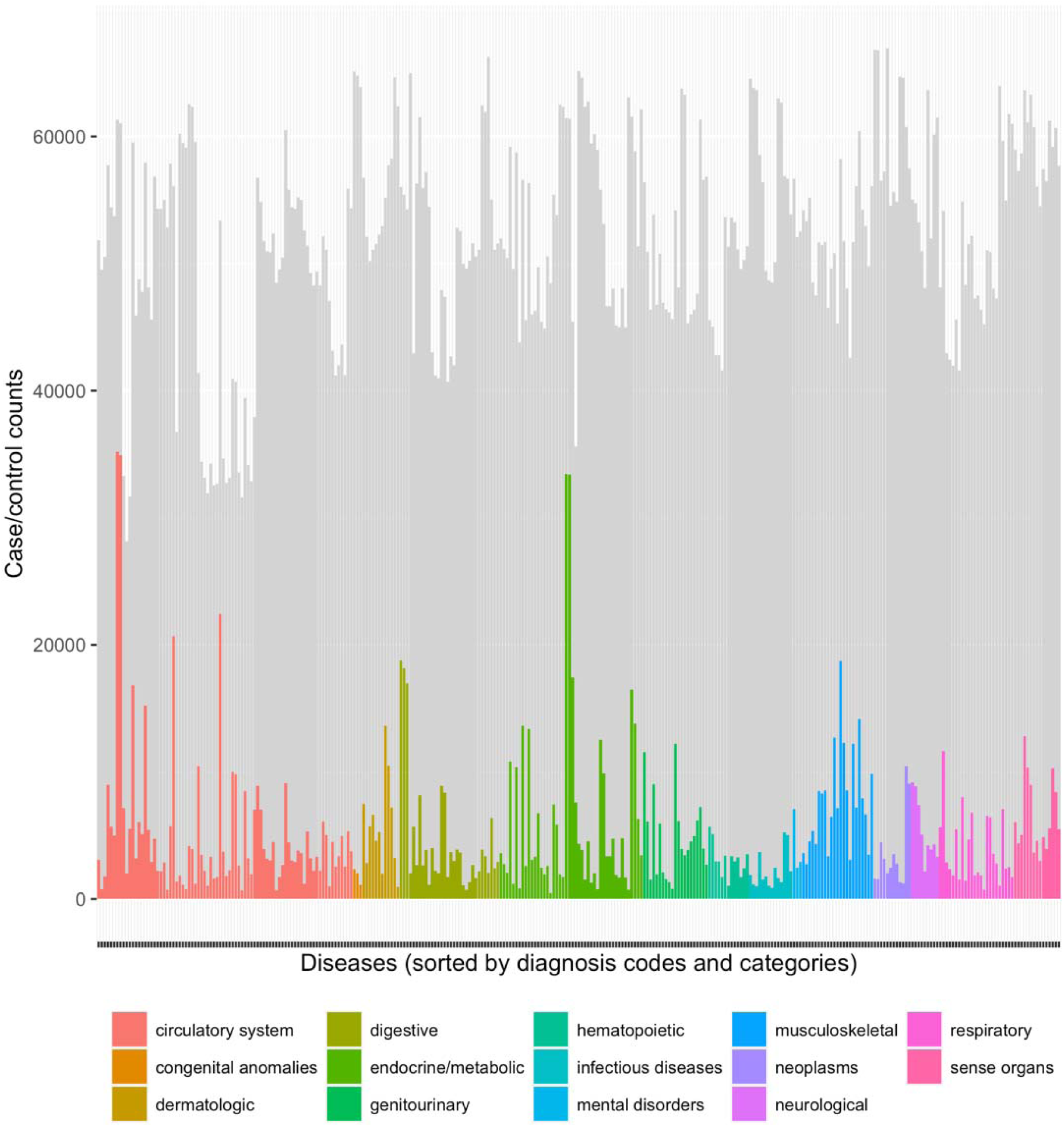
Sample size distribution of 309 disease phecodes for the eMERGE III individuals of European ancestry.Diseases were sorted according the phecodes. Colored area represented cases and greyed area represented controls. Different colors corresponded to different categories of diseases.

**Figure 3.**
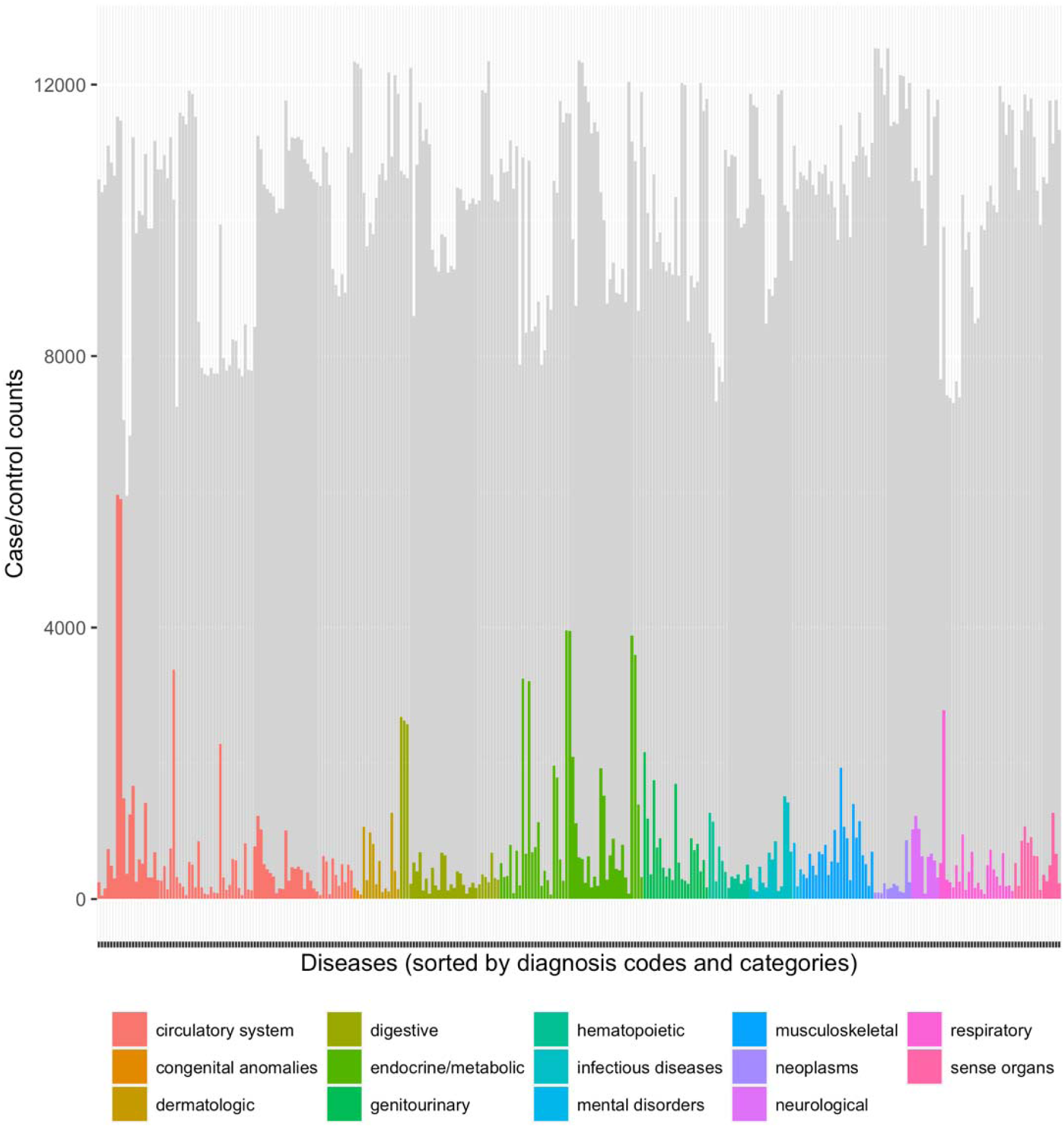
Sample size distribution of 309 disease phecodes for the eMERGE III individuals of African ancestry. Diseases were sorted according the phecodes. Colored area represented cases and greyed area represented controls. Different colors corresponded to different categories of diseases. The African ancestry population had less numbers of samples (both cases and controls) for all categories of diseases in eMERGE III.

**Figure 4.**
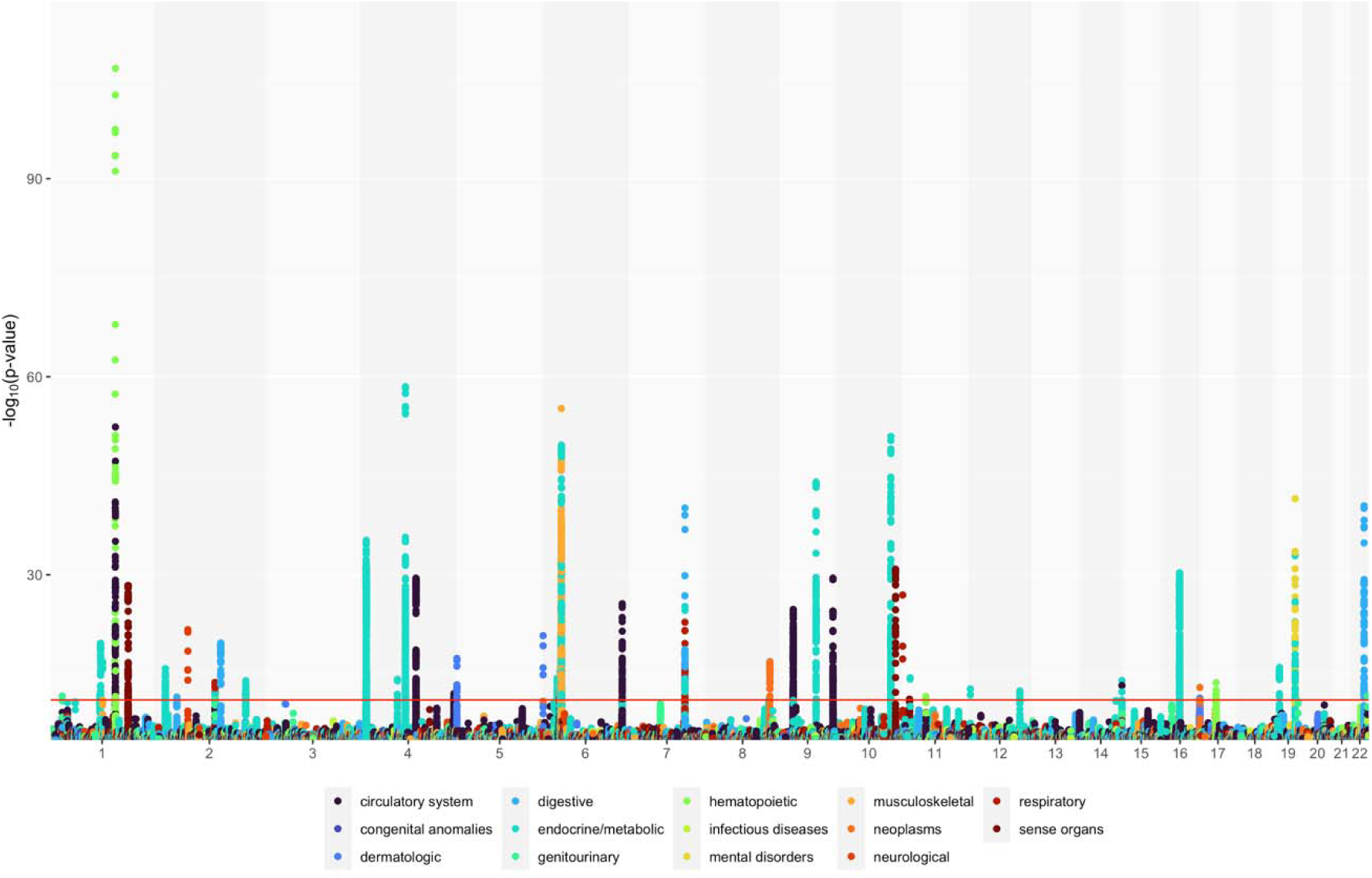
Variant-based PheWAS of 309 diseases in the eMERGE III European ancestry population (total N = 67,885). X-axis represented chromosome positio. Y-axis was negative log10 transformed p-values. Red horizontal line was the Bonferroni significance threshold (p = 8.96e-12). Colors represented different disease categories.

**Figure 5.**
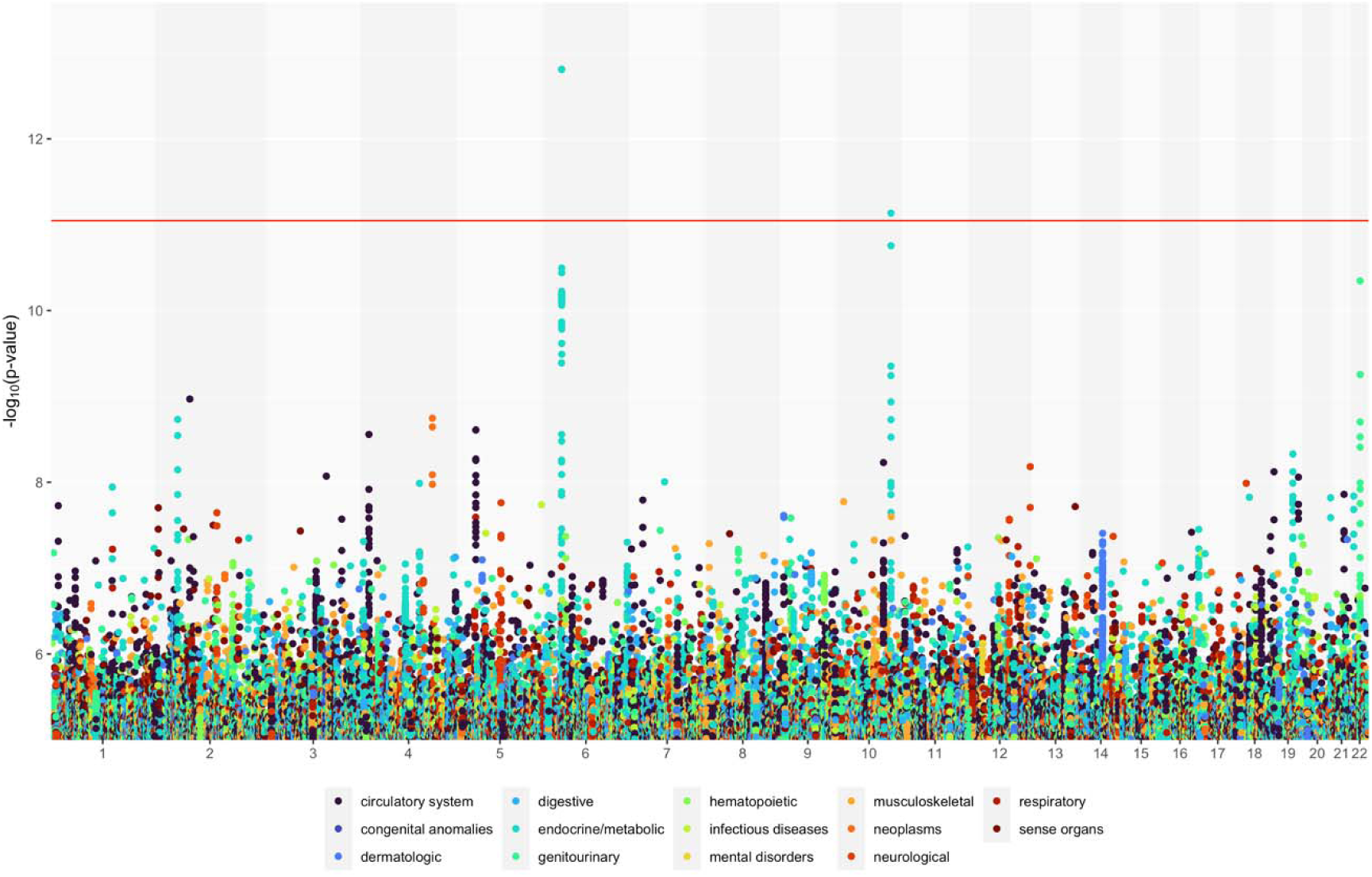
Variant-based PheWAS of 309 diseases in the eMERGE III African ancestry population (total N = 12,658). X-axis represented chromosome position. Y-axis was negative log10 transformed p-values. Red horizontal line was the Bonferroni significance threshold (p = 8.96e-12). Colors represented different disease categories. Only a few GWAS signals exceeded the Bonferroni significance threshold likely due to multiple testing burden and reduced power due to limited sample sizes.

Ancestry-specific phenome-wide GWAS summary statistics, combined with the MASHR-based GTEx v8 eQTL models, were input into S-PrediXcan [20] to estimate the gene-disease associations for the eMERGE III European ancestry and African ancestry datasets, respectively. For each ancestry-based subset, S-PrediXcan tested for the gene-disease associations among 309 diseases phenotypes and 22,535 distinct genes in each of the 49 primary human tissues or cell lines independently. S-MultiXcan [14] followed S-PrediXcan to aggregate association results across all tissues to identify cross-tissue gene-disease associations for the eMERGE III European ancestry and African ancestry datasets, respectively. We further conducted ancestry-specific colocalization analyses in eMERGE III to further prioritize likely functional genes with a locus regional colocalization probability (locus RCP).

For cross-ancestry analyses, we first meta-analyzed the ancestry-specific phenome-wide GWAS summary statistics using Stouffer’s Z-score method that was implemented METAL [21]. To ensure interpretability of meta-analysis results, variants were retained only if they were present in both eMERGE datasets and had same direction of effect for each variant in in both subsets. We then implemented S-PrediXcan, followed by S-MultiXcan, on the meta-analyzed GWAS summary statistics to identify cross-ancestry gene-disease associations, respectively.

Statistically significant discoveries from the eMERGE III phenome-wide TWAS (p < 1.47e-10) were replicated in the individuals from PMBB that were also subset into two ancestry groups: individuals of European ancestry (N = 6,967) and individuals of African ancestry (N = 8,506). We also performed ancestry-specific colocalization analyses using fastENLOC [17]. Second, we validated the multi-ancestry gene-disease association against multiple publicly accessible, centralized genetics research databases, including PheWAS Catalog [18] and PhenomeXcan [56] on the UK Biobank individuals of European ancestry [22] as well as systematic literature review.

To control false positives due to multiple testing, we considered a gene-disease association significance in the eMERGE III discovery analyses only when it exceeded the Bonferroni-corrected statistical significance threshold (p < 1.47e-10). This significance threshold accounted for the total number of possible tests across 22,535 genes, 309 diseases, and 49 primary human tissues. Associations were considered a statistically significant replication in the independent PMBB replication cohorts if they exceeded the replication significance threshold (p ≤ 6.39e-5), which accounted for the total number of replications performed in the PMBB. A gene-disease association was considered a statistically significant validation in the independent PhenomeXcan data if the association was statistically significant in the original PhenomeXcan analyses, i.e. PhenomeXcan p ≤ 5.49e-10.

### eMERGE network

The eMERGE network is a National Human Genome Research Institute (NHGRI) funded multi-site consortium created to facilitate large-scale genomic research by exploring the utility of DNA biorepositories linked with EHR data collected within health care systems. Here, we used eMERGE network phase III data (hereafter referred to as eMERGE III data) as the discovery dataset [23]. eMERGE III consists of nine study sites, two sequencing and genotyping facilities, and a coordinating center. The eMERGE III data used in this study were updated on February 13, 2020, comprising 105,109 participants recruited into the respective biorepositories via clinics within health systems or community settings. eMERGE III participants were from different genetic ancestry backgrounds (77.7% European ancestry, 16.1% African ancestry, and 6.2% others).

#### Penn Medicine BioBank (PMBB)

The PMBB is a single-institutional research program that recruits participants throughout the University of Pennsylvania Health System by enrolling at the time of outpatient visits. Patients participate by completing a questionnaire, donating a blood sample, allowing researchers access to their EHR information, and agreeing to future recontact [9]. The PMBB has recruited approximately 90,000 participants by October 2021 and is still actively recruiting new participants. 16,867 unique participants have been genotyped in one batch by the Illumina Global Screening Array Version 2 (GSA_V2) chip, which are the data used for this study.

#### Genotype data and quality control in the eMERGE discovery dataset

eMERGE III DNA samples were genotyped across multiple genotyping platforms. Genotype data across different genotyping platforms were combined after imputation to the Haplotype Reference Consortium (HRC1.1) that contains the 1000 Genomes globally diverse sample set as well as a large balance of European ancestry samples using the minimac3 algorithm implemented on the Michigan Imputation Server (MIS) [24,25]. The eMERGE III imputed dataset consisted of in total 39,131,578 genetic variants, 73,384 European ancestry (82.9%) and 12,658 African ancestry (17.1%) out of the 88,518 eMERGE III adult participants (age >= 19). QC of the eMERGE III imputed data followed the suggested steps by Verma *et al*. [26], including imputation score filtering (R^2^ > 0.3), removal of palindromic variants, biallelic variant check, genotype call rate (> 99%) and sample call rate (>99%) filtering, minor allele frequency filtering (MAF > 1%), Hardy-Weinberg equilibrium test (p-value > 1e-8). The related samples were kept in the final QC’d imputed data as Regenie can properly account for relatedness using linear mixed models. Subsequent principal component analysis (EIGENSOFT [27]) projected all individuals that passed QC onto the 1000 Genomes Project Phase 3 [28] sample space to generate principal components to adjust for population stratification. Based on the percent of variance explained, the first four and eight principal components (PCs) estimated by SmartPCA in EIGENSOFT were used as covariates to adjust for population structure in the subsequent analyses for eMERGE III African ancestry and European ancestry populations, respectively. Approximately six million SNPs and 67,885 eMERGE III individuals of European ancestry survived the post-imputation QC. Approximately eleven million SNPs and 12,658 eMERGE III individuals of African ancestry survived the post-imputation QC. For subsequent phenome-wide association analyses, we further performed phenotype-specific QC on the genotype data, including MAF > 1% and Hardy-Weinberg Equilibrium tests (p-value > 1e-8) to ensure satisfactory SNP quality for each interrogated phenotype and avoid biases in this study.

### Phenotype data and quality control in the eMERGE discovery dataset

We used disease diagnosis codes from eMERGE III to classify individuals into binary case and control groups for each disease phenotype. eMERGE III contained a variety of health data, including International Classification of Diseases, Ninth and Tenth Revision, Clinical Modification (ICD-9-CM, ICD-10-CM) codes, clinical lab variables, medications, demographics etc. ICD-9-CM (mainly before 2015) and ICD-10-CM (2015 and onward) codes describe patient disease diagnoses and procedures observed and entered into the EHR during clinical practice. To better define disease phenotypes and obtain groups of clear controls for the phenome-wide study, we first mapped ICD-CM codes to phecodes and then determined case/control status for EHR-linked biobank participants across a variety of diseases using phecodes and the *phewas* R package [29]. ICD-10-CM codes were mapped to ICD-9-CM using a combination of the Center for Medicare and Medicaid Services 2017 General Equivalency Mappings information (https://www.cms.gov/Medicare/Coding/ICD10/2017-ICD-10-CM-and-GEMs.html) and manual curation. Phecodes [29] are the manually curated medical codes that aggregate one or more related ICD-CM codes into distinct diseases or traits. Phecodes additionally develop exclusion criteria to identify clinically reasonable, clean controls to reduce confounding effects. Phenotypes for each eMERGE participant were determined by mapping ICD-9 codes to distinct phecodes via Phecode Map 1.2 [18] embedded in the R package “*PheWAS*” [30] under rule-of-two. Rule-of-two considered an individual to be a case of a certain disease when they have two or more incidents of the corresponding phecode. Controls were those who did not have any incidents of the phecode and met the phecode inclusion criteria of controls as described above. Individuals with one incident of the phecode were considered to have a “missing phenotype” and thus, excluded from any subsequent statistical analyses. To ensure sufficient statistical power [31], all statistical analyses in this study only considered disease phenotypes that had case numbers greater than 50 in either the eMERGE III individuals of European ancestry or the eMERGE III individuals of African ancestry. Similarly, the disease phenotypes were required to also have greater than 50 cases in the PMBB individuals of European ancestry or individuals of African ancestry to guarantee sufficient sample sizes for replication. In addition, subsequent association analyses excluded phecodes that represent non-specific diseases, symptoms, or injuries after manual curation. The number of phecode-derived disease phenotypes came down to 309 after thorough phenotype QC (**Supplementary Table 1**).

### Genome-wide PheWAS and harmonization of summary statistics

We adopted Regenie for the genome-wide PheWAS analyses in the eMERGE III individuals of European ancestry and the individuals of African ancestry, respectively. Regenie is a whole-genome association tool developed by Mbatchou *et al*. [19] that adopts linear mixed regression models and runs orders of magnitude faster than other equivalent tools. Regenie proceeds in two main steps. Step 1 fits ridge regression models to a set of consecutive blocks of SNPs across the whole genome to account for local LD structures. Step 2 uses the prediction from the step 1 and is completely decoupled from step 1 to facilitate phenome-wide analyses [19].

#### Regenie in eMERGE III

Step 1 used LD pruned whole-genome imputed data that reduced the number of variants to about 50K [19]. LD pruning on QC’d eMERGE III European ancestry imputed data using ‘– indep-pairwise 1000 100 0.3’ in PLINK yielded 563,320 variants and 68,813 samples for Regenie Step 1. Similarly, LD pruning on QC’d eMERGE III African ancestry imputed data using ‘–indep-pairwise 1000 100 0.15’ in PLINK yielded 544,686 variants and 12,658 samples for Regenie Step 1. We ran Regenie Step 1 for each disease phenotype separately to account for the differential case/control numbers in different diseases. Covariates included sex, decade of birth, first four PCs for the eMERGE III individuals of African ancestry (first eight PCs for the eMERGE III individuals of European ancestry), and nine eMERGE adult site codes (one for each eMERGE site). Step 1 also used a shrinkage parameter (block size = 1000) and leave-one out cross validation scheme. Different from step 1, step 2 of Regenie uses the whole QC’d imputed data, but the same set of covariates as in step 1, the same shrinkage parameter (block size = 1000) and leave-one out cross validation scheme. We also adopted Firth likelihood ratio test as a fallback for p-values less than 0.01. Overall, Regenie interrogated genetic associations between six million SNPs in the eMERGE European ancestry population (eleven million in the eMERGE African ancestry population) and 309 disease phecodes.

#### Harmonization of GWAS summary statistics

Harmonization of genome-wide phenome-wide GWAS summary statistics followed the examples provided at the GitHub WiKi page (https://github.com/hakyimlab/summary-gwas-imputation/wiki/GWAS-Harmonization-And-Imputation). Exemplary and reference datasets were downloaded from https://zenodo.org/record/3569954#.XiiurVOQHRb. The LD blocks were defined based on the results of Berisa and Pickrell, 2015 [32], and are ancestry-specific. Files of the Berisa-Pickrell LD blocks were downloaded from https://bitbucket.org/nygcresearch/ldetect-data/src/master/. The downloaded LD blocks were based on GRCh37. Liftover of LD files from GRCh37 to GRCh38 were conducted using the UCSC liftover tool (https://genome.ucsc.edu/cgi-bin/hgLiftOver). We first harmonized eMERGE ancestry-specific phenome-wide GWAS summary statistics onto the format expected by TWAS methods using *gwas_parsing*.*py* available at https://github.com/hakyimlab/summary-gwas-imputation. eMERGE variant positions were based on GRCh37. *gwas_parsing*.*py* also performed liftover for eMERGE from GRCh37 to GRCh38 provided a proper chain file downloaded from UCSC (hg19ToHg38.over.chain.gz).

#### Imputation of GWAS summary statistics

Certain eQTLs may be missing in the genotype data due to various reasons. To improve TWAS power, it was critical to impute GWAS summary statistics for the eQTLs that were missing in the genotype data. Instruction of GWAS summary statistics imputation was provided at the GitHub (https://github.com/hakyimlab/MetaXcan/wiki/Tutorial:-GTEx-v8-MASH-models-integration-with-a-Coronary-Artery-Disease-GWAS). Our imputation used variants and a covariance matrix generated from the1000 Genomes Project data as the reference panel, and ancestry-specific LD blocks defined by Berisa and Pickrell (appropriately lifted over from GRCh37 to GRCh38 using UCSC) [32]. We used a Ridge-like regularization when inverting the covariance matrix (regularization = 0.1) and only retained variants with minor allele frequency > 1%. After GWAS summary statistics imputation, we compiled the imputed and the original GWAS summary statistics into one dataset for downstream TWAS analyses.

### Ancestry-specific TWAS using genome-wide phenome-wide GWAS summary statistics

We performed gene-based association analyses using S-PrediXcan [20] and S-MultiXcan [14] on the eMERGE III individuals of European ancestry and the individuals of African ancestry, respectively. S-PrediXcan is a tissue-specific TWAS method that integrates GWAS summary statistics and eQTL databases to estimate each gene’s effect on a trait of interest in a tissue-specific way [20]. S-MultiXcan is a cross-tissue TWAS method that leverages gene-disease association information across all tissue to improve TWAS capability of identifying potential complex trait-related genes [14]. Both S-PrediXcan and S-MultiXcan are available under the MetaXcan GitHub repository (https://github.com/hakyimlab/MetaXcan). The repository was forked on April 21^st^, 2020. The eQTL models for 49 primary human tissues that used the latest GTEx v8 reference transcriptomic data were downloaded and available from a Zenodo repository (https://zenodo.org/record/3518299) [33]. These eQTL models computed a putative eQTL effect size on gene expression levels using MASHR, which incorporates biological information, on DAP-G fine-mapped variables. Covariances among SNPs in each specific primary human tissue are also available at the same Zenodo repository.

Both S-PrediXcan and S-MultiXcan used ancestry-specific GWAS summary statistics derived from the eMERGE III individuals of European ancestry and the individuals of African ancestry, separately, to perform ancestry-specific TWAS. For each population, we ran S-PrediXcan to identify associations between 309 phecodes and 22,255 genes, tissue by tissue, for all the 49 primary human tissues and cell lines. Then, S-MultiXcan took in GWAS summary statistics, all 49 eQTL databases, and all tissue specific S-PrediXcan results on a certain disease to estimate the joint effect of a gene’s expression levels across tissues on a trait. At the end of this step, we obtained tissue-specific and cross-tissue gene-disease associations across 22,255 genes and 309 phecodes for each of the eMERGE III ancestry-based population.

### African ancestry-specific TWAS using individual-level data

We carried out PrediXcan for individuals of African ancestry within eMERGE III. PrediXcan used the LD structure indigenous to the testing populations, thus, it can better account for the LD structure for the study cohort of interest than S-PrediXcan.

PrediXcan required additional QC. We removed related samples (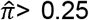; one individual was dropped from each related pair) and performed another round of QC on the imputed data to ensure proper genotype data quality. In the eMERGE III AA population, 11,177,244 genetic variants and 12,290 unrelated individuals passed QC.

We ran PrediXcan on eMERGE AA population across 309 phecodes, 22,255 total genes, and 49 primary human tissues and cell lines, using MASHR-based eQTL models based on GTEx v8 reference data [11,33]. To speed up PrediXcan, we used *Predict*.*py* provided in the MetaXcan GitHub repository (https://github.com/hakyimlab/MetaXcan) to predict gene expression levels of 22,255 genes in 49 primary human tissues separately for eMERGE individuals of African ancestry and PMBB individuals of African ancestry. Then, we used *glm* function, coupled with *doParallel* (https://cran.r-project.org/web/packages/doParallel/index.html) and *foreach* (https://cran.r-project.org/web/packages/foreach/index.html) packages, to achieve fast, parallel computing in R.

### Cross-ancestry TWAS

We combined the eMERGE III ancestry-specific genome-wide phenome-wide GWAS summary statistics in a meta-analysis implemented METAL [21]. METAL combined Z-scores for each allele across the two populations in a weighted sum, with weights proportional to the square-root of sample sizes in each ancestry using Stouffer’s Z-score method. Only variants that were present in both ancestry populations and had the same direction of effect were retained for cross-ancestry analysis. At the end of this step, we combined eMERGE European ancestry-specific and African ancestry-specific genome-wide phenome-wide GWAS summary statistics for 309 phecodes.

S-PrediXcan and S-MultiXcan used meta-analyzed genome-wide phenome-wide GWAS summary statistics to identify gene-disease associations that are shared across the eMERGE III individuals of European ancestry and the individuals of African ancestry. Similar to ancestry-specific TWAS analyses, S-PrediXcan integrated meta-analyzed GWAS summary statistics with MASHR-based eQTL models [33] for 309 phecodes and 49 primary human tissues and cell lines. Then, S-MultiXcan took the same inputs of the S-PrediXcan, plus S-PrediXcan tissue-specific gene-disease association results across all tissues, to identify the cross-ancestry and cross-tissue gene-disease associations. At the end of this step, we obtained tissue-specific and cross-tissue gene-disease association signals that were shared between the eMERGE III individuals of European ancestry and the eMERGE III individuals of African ancestry.

### Colocalization analyses

We performed statistical fine-mapping of likely causal variants and calculated GWAS posterior inclusion probability (PIP) by applying a Bayesian method, TORUS [34], on the eMERGE III ancestry-specific phenome-wide GWAS summary statistics. Then, we estimated the probability of colocalization between each of the GWAS and cis-eQTL signals using fastENLOC developed by Pividori *et al*. [56]. Ancestry-specific haplotype blocks were provided by Berisa and Pickrell after properly being lifted over to GRCh38 [32], in concordance with the genome build of the GTEx v8 eQTLs. We interrogated a total of 6,463,721 and 11,189,509 observed and imputed GWAS signals for the eMERGE III individuals of European ancestry and the eMERGE III individuals of African ancestry, respectively. The locus regional colocalization probability (RCP) for each signal cluster of interest was calculated automatically by fastENLOC. Given multiple tissues considered in this study, the maximum colocalization probability across all tissues was used, following PhenomeXcan [17]. Locus RCP > 0.1 provided additional functional evidence for putative causal gene prioritization.

### Replication in Penn Medicine BioBank (PMBB)

#### Genotype QC

Different from eMERGE III, PMBB DNA samples were all genotyped using the same Illumina Global Screening Array Version 2 (GSA_V2) platform in 2020. We performed genotype imputation for the PMBB data using Eagle2 [35] and Minimac [36] software on the TOPMed Imputation Server (MIS) [25]. Imputation was performed for all autosomes, with TOPMed version R2 on GRCh38 reference panel [37]. Cosmopolitan post-imputation QC included imputation score filtering (R^2^ > 0.7), removal of palindromic variants, biallelic variant check, sex check, genotype call rate (> 99%) and sample call rate (>99%) filtering, minor allele frequency filtering (MAF > 1%), Hardy-Weinberg equilibrium test (p-value > 1e-8) [26]. We generated PCAs to adjust for population structure and to identify genetically informed ancestry (GIA) from EIGENSOFT [27]; this process classified 8,506 PMBB samples as individuals of African ancestry and 6,967 individuals as individuals of European Ancestry. Ancestry-specific QC followed the same QC parameters used in the cosmopolitan PMBB QC. Given the percent of variance explained, the first two and five PCs estimated by SmartPCA in EIGENSOFT were used as covariates to adjust for population structure in the subsequent analyses for PMBB AA and EA populations, respectively. At the end, approximately nine million SNPs and 6,967 PMBB participants of European ancestry survived the post-imputation QC. Approximately fifteen million SNPs and 8,506 PMBB participants of African ancestry survived the post-imputation QC.

#### Phenotype data QC

PMBB phenotype data quality control followed the same procedures as that on the eMERGE III data. The same 309 disease phecodes as in the eMERGE III survived the phenotype data QC in the PMBB.

#### Regenie in PMBB

Regenie in PMBB was similar to that in eMERGE III for the purpose of proper replication. Step 1 used LD pruned whole-genome imputed data. LD pruning on QC’d PMBB individuals of European ancestry imputed data using ‘–indep-pairwise 1000 100 0.3’ in PLINK yielded 483,787 variants and 6,967 samples for Regenie Step 1. Similarly, LD pruning on QC’d PMBB individuals of African ancestry imputed data using ‘–indep-pairwise 1000 100 0.15’ in PLINK yielded 648,134 variants and 8,506 samples for Regenie Step 1. Regenie Step 1 fit regression models on each disease phenotype separately. Covariates included sex, age, first two PCs for PMBB individuals of African ancestry (first five PCs for PMBB individuals of European ancestry). Step 1 also used shrinkage parameter (block size = 1000) and leave-one out cross validation scheme. Distinguished from step 1, step 2 of Regenie used the whole QC’d imputed data, but the same set of covariates as in step 1, shrinkage parameter (block size = 1000) and leave-one out cross validation scheme. We also adopted Firth likelihood ratio test in step 2 as fallback for p-values less than 0.01. Overall, Regenie interrogated genetic associations between nine million SNPs in PMBB individuals of European ancestry (fifteen million in PMBB individuals of African ancestry) and 309 phecodes.

#### Harmonization and post-GWAS process of ancestry-specific genome-wide phenome-wide GWAS summary statistics

This step followed the same procedures as that on the eMERGE III data.

#### Replication of statistical analyses on PMBB

. We replicated the whole phenome-wide TWAS and colocalization analyses in PMBB. Ancestry-specific S-PrediXcan and S-MultiXcan used the genome-wide phenome-wide GWAS summary statistics as described above and estimated tissue-specific and cross-tissue gene-disease associations across 309 phecodes, 22,255 genes for PMBB individuals of European ancestry (N = 6,967) and individuals of African ancestry (N = 8,506) population, respectively. African ancestry-specific PrediXcan that used individual-level PMBB genotype data processed approximately eleven million genetic variants and 7,978 unrelated individuals. For cross-ancestry TWAS, we applied S-PrediXcan and S-MultiXcan on meta-analyzed PMBB European ancestry-specific and African ancestry-specific genome-wide phenome-wide GWAS summary statistics (total N = 15,473). At the end, we obtained ancestry-specific and cross-ancestry gene-disease associations across the same 309 phecodes and 22,255 genes in PMBB.

### Validation using PhenomeXcan, PheWAS Catalog, and systemic literature search

We validated the statistically significant gene-disease associations that we found in eMERGE III European ancestry-specific, African ancestry-specific, and cross-ancestry analyses by seeking supportive evidence in PhenomeXcan (a gene-level phenome-wide research using UK Biobank health data) [17], PheWAS Catalog (a PheWAS catalog on the Vanderbilt BioVU biobank) [18], and systemic literature review.

PhenomeXcan performed comparative phenome-wide gene-level genomics analyses (including TWAS and colocalization) on UK Biobank, a population-based biobank with a larger sample size than eMERGE III, nevertheless made up by a majority of European ancestry, healthy individuals. The UK Biobank phenotype data comes from a combination of health care data and enrollment surveys [22]. PhenomeXcan includes gene-based associations generated by TWAS for 4091 traits and 22,535 genes in 49 primary human tissues or cell lines using the MASHR-based eQTL models derived from the GTEx v8 reference transcriptome panel. We downloaded the summary PhenomeXcan results from https://doi.org/10.5281/zenodo.3530669 on October 9th, 2020. UK Biobank adopts the ICD-10 coding system, different from the ICD-10-CM version adapted in the US. Descriptions of ICD-10 and its adaption, ICD-10-CM, to US medical coding system can be found at the CDC website (https://www.cdc.gov/nchs/icd). The 3-digit and 5-digit ICD-10 codes in the form of long titles were available and downloaded from ftp://ftp.cdc.gov/pub/Health_Statistics/NCHS/Publications/ICD10 at October 8^th^, 2020. To match phenotypes, phecodes were first mapped to the ICD-10 using the mapping file provided by PheWAS catalog at https://phewascatalog.org/phecodes_icd10 [38]. We manually curated the phecode to ICD-10 mapping list by cross-referencing medical code descriptions and the presence of corresponding ICD-10 codes in the PhenomeXcan results. Since PhenomeXcan only looked into the three-digit ICD-10 codes, we only retained the corresponding three-digit phecodes unless a leaf phecode mapped to another specific ICD-10 three-digit category. There were in total 236 matching phecode-PhenomeXcan ICD-10 pairs, encompassing 127 unique three-digit phecodes to 212 unique three-digit ICD-10 codes (**Supplementary Table 2**). An association was validated by PhenomeXcan if the gene-disease pair reached the original statistical significance threshold in the PhenomeXcan study (p < 5.49e-10).

The PheWAS Catalog [18] hosted associations for 1,358 phecode-defined disease phenotypes in 13,835 individuals of European ancestry in the Vanderbilt DNA biobank, BioVU. As BioVU is one of the eMERGE III study sites, we expected that many of the samples in the PheWAS catalog were also included in the eMERGE III. Thus, we only used PheWAS Catalog as a proof-of concept validation set to test for the effectiveness of our analytic framework. We considered our analytic framework effective if our analytic framework were able to identify the PheWAS Catalog associations with a greater statistical significance.

### Statistical correction

To account for multiple tests and control for type I error rates, we adapted a Bonferroni-corrected significance threshold for the discovery analysis in the eMERGE III data. Bonferroni-corrected significance threshold was corrected by the total possible number of tests among 22,535 genes, 309 disease phenotypes and 49 primary human tissues or cell lines, which resulted in 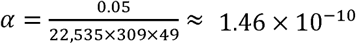. The replication significance threshold was corrected for the number of statistically significant associations to be replicated (N = 803) in the PMBB data, which gave 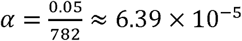.

## Results

### Overview of phenome-wide gene-disease analyses in eMERGE III

We found a total of 782 statistically significant unique gene-disease associations with a p-value less than the Bonferroni-corrected significance threshold (p < 1.47e-10), encompassing 296 unique genes and 239 disease phenotypes (**Figure 6**). The 782 statistically significant gene-disease associations included results from the ancestry-specific analyses and cross-ancestry analysis. 74 (9.5%) of the 782 statistically significant gene-disease associations were found exclusively by the cross-ancestry analysis. 82 (10.5%) of the 782 were found only in the eMERGE III individuals of European ancestry. 394 (50.4%) of the 782 were found only in the eMERGE III individuals of African ancestry. The rest 232 (29.7%) gene-disease associations were found in both ancestry-specific or cross-ancestry analyses, most overlapping between the European ancestry-specific analysis and the cross-ancestry analysis.

**Figure 6.**
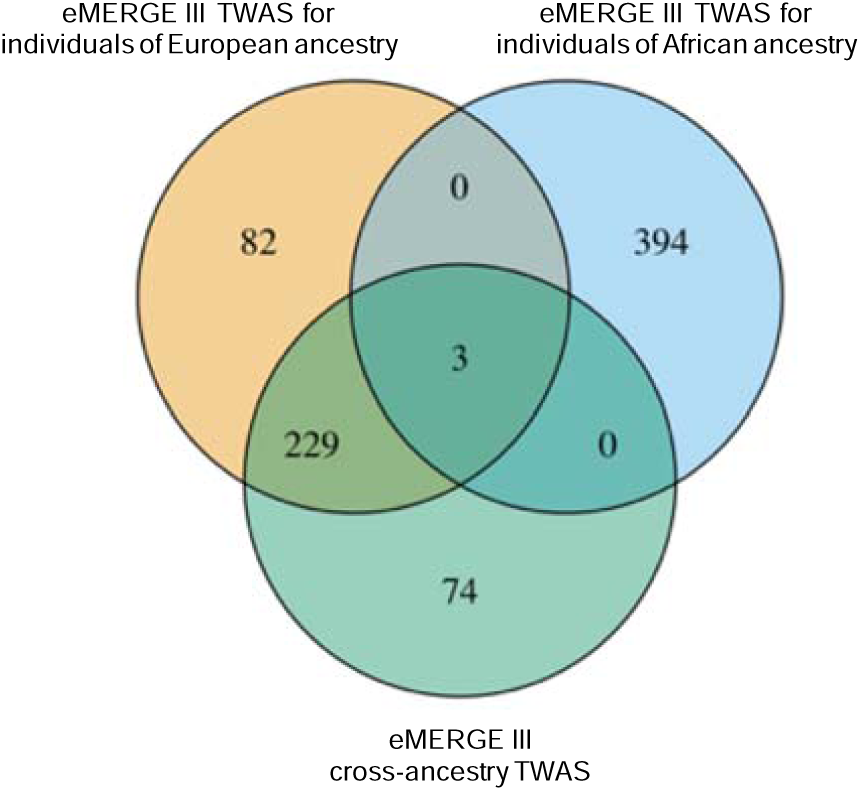
Overlaps of statistically significant gene-disease associations (p < 1.47e-10) among the eMERGE III ancestry-specific TWAS and cross-ancestry TWAS.

eMERGE III African ancestry-specific analysis contributed over half (50.4%) of the unique statistically significant associations, while the sample size was about 19% of that of the eMERGE III individuals of European ancestry. Moreover, eMERGE III African ancestry-specific analysis unearthed associations with 218 different phecode-defined disease phenotypes, which was more than triple the amount of that on the eMERGE III European ancestry-specific analysis (64 significant phecode-defined disease phenotypes).

### Gene-disease analyses for individuals of European ancestry

Phenome-wide TWAS found in total 314 statistically significant gene-disease associations in the eMERGE III individuals of European ancestry (p < 1.47e-10) (**Figure 7**; **Supplementary Table 3**) where 36 (10.8%) of the 314 associations had locus regional colocalization probability (RCP) > 0.1. The 314 statistically significant associations comprised 120 unique genes and 64 unique phecode-defined disease phenotypes in 11 disease categories. 82 (26.1%) of the total 314 were unique to the eMERGE III individuals of European ancestry. 229 (72.9%) were replicated in the cross-ancestry analysis. And only 3 were shared with the eMERGE III individuals of African ancestry.

**Figure 7.**
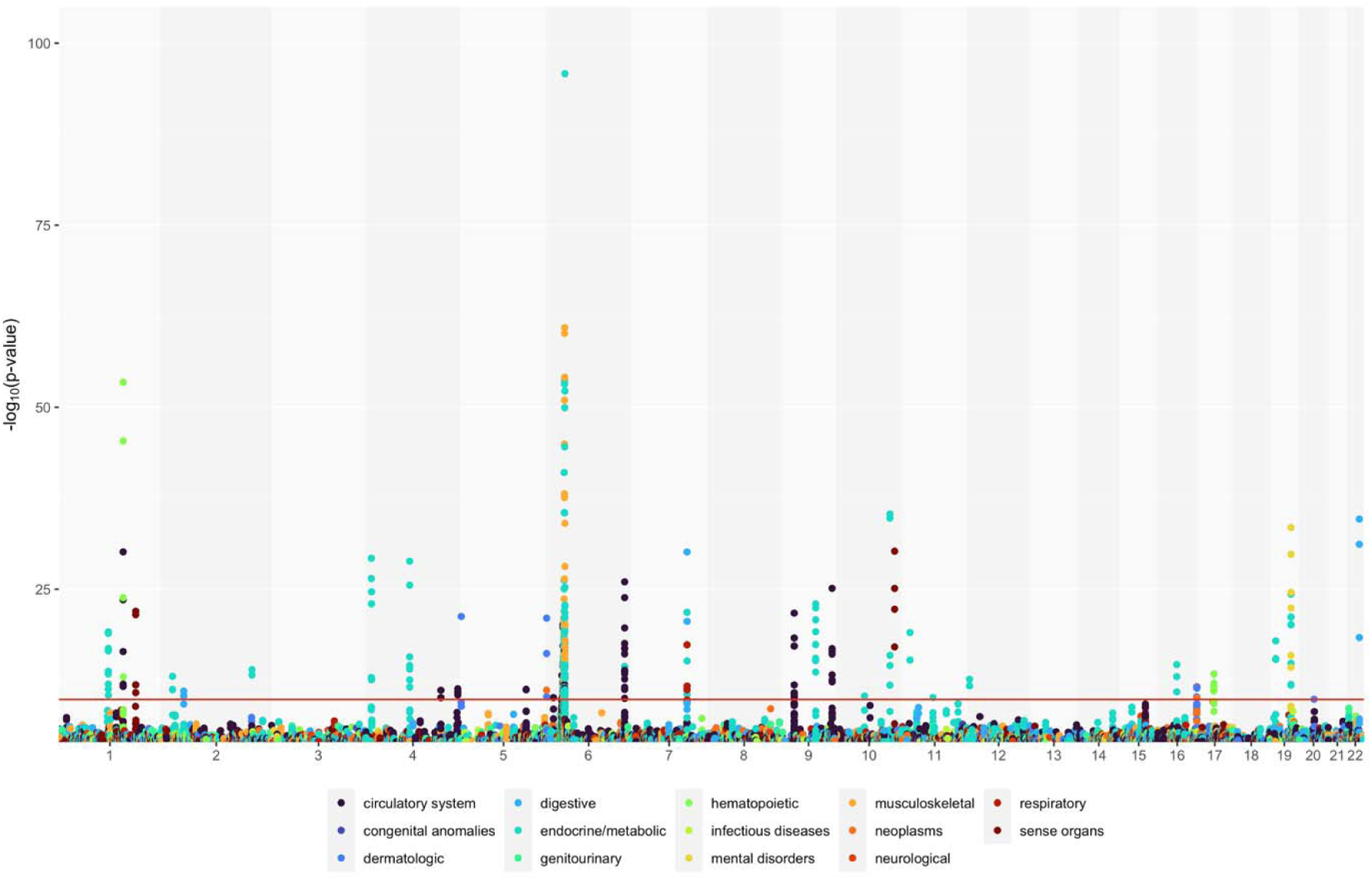
Gene-disease associations for the eMERGE III individuals of European ancestry. X-axis represented chromosome position. Y-axis was negative log10 transformed p-values. Red horizontal line was the Bonferroni significance threshold (p = 1.47e-10). Colors represented varied categories of diseases.

We first looked for validation against the PheWAS catalog [18]. The PheWAS catalog reports associations between 3,144 SNPs (previously associated with complex human diseases in the GWAS catalog [39]) and 1,358 phecode-derived disease phenotypes in 13,835 individuals of European ancestry from the Vanderbilt BioVU [18]. Due to the limited number of inspected SNPs, PheWAS catalog did not have association information for 227 of the 314 statistically significant associations that we found in the eMERGE III European ancestry-specific analysis. The remaining 87 gene-disease associations were present in the PheWAS catalog with at least nominal statistical significance (PheWAS catalog p < 0.05). Almost all of these 87 gene-disease associations, except the one between *ABCG2* and gout, were assigned greater statistical significance in the eMERGE III dataset. We expected that many of the samples in the PheWAS catalog were also included in the eMERGE III as the Vanderbilt BioVU is one of the participating sites. Therefore, we considered these validations a strong proof-of-concept that our analytic framework worked effectively.

We then sought validations using the independent PhenomeXcan results [17] (**Table 1)**. PhenomeXcan tested for gene-disease associations across 4,091 traits and 22,535 genes in individuals of European ancestry (N = 361,194) from the UK Biobank. Only 127 of the total 309 phecodes were successfully mapped to 212 ICD-10 diagnosis codes in UK Biobank after automatic mapping and manual curation (see **Methods**), amassing to 236 phenotype matching eMERGE-PhenomeXcan disease pairs. As a result, only 179 statistically significant associations from the eMERGE III European ancestry-specific analyses had corresponding ICD-10 disease phenotypes in the PhenomeXcan. Among these 179 gene-disease associations, we successfully replicated 95 (53.1%) unique associations in the PhenomeXcan at phenomexcan.org (PhenomeXcan Bonferroni-corrected significance threshold at p ≤5.49e-10 [17]). Some of the PhenomeXcan validated associations have also been reported in previous studies. The most significant replication was observed between *HLA-DQA2* and type 1 diabetes (p = 4.94e-86) (**Supplementary Table 3**). *HLA* genes has long been associated with type 1 diabetes. However, due to complexity of the polymorphisms in the *HLA* region, the association between *HLA-DQA2* and type 1 diabetes needs further validation perhaps with analysis of *HLA* haplotypes [40]. Another example was the association between *PHACTR1* and coronary atherosclerosis (p = 1.82e-11) [41], which was further supported by a locus RCP at 0.9998 in eMERGE III (**Supplementary Table 3**).

**Table 1.**
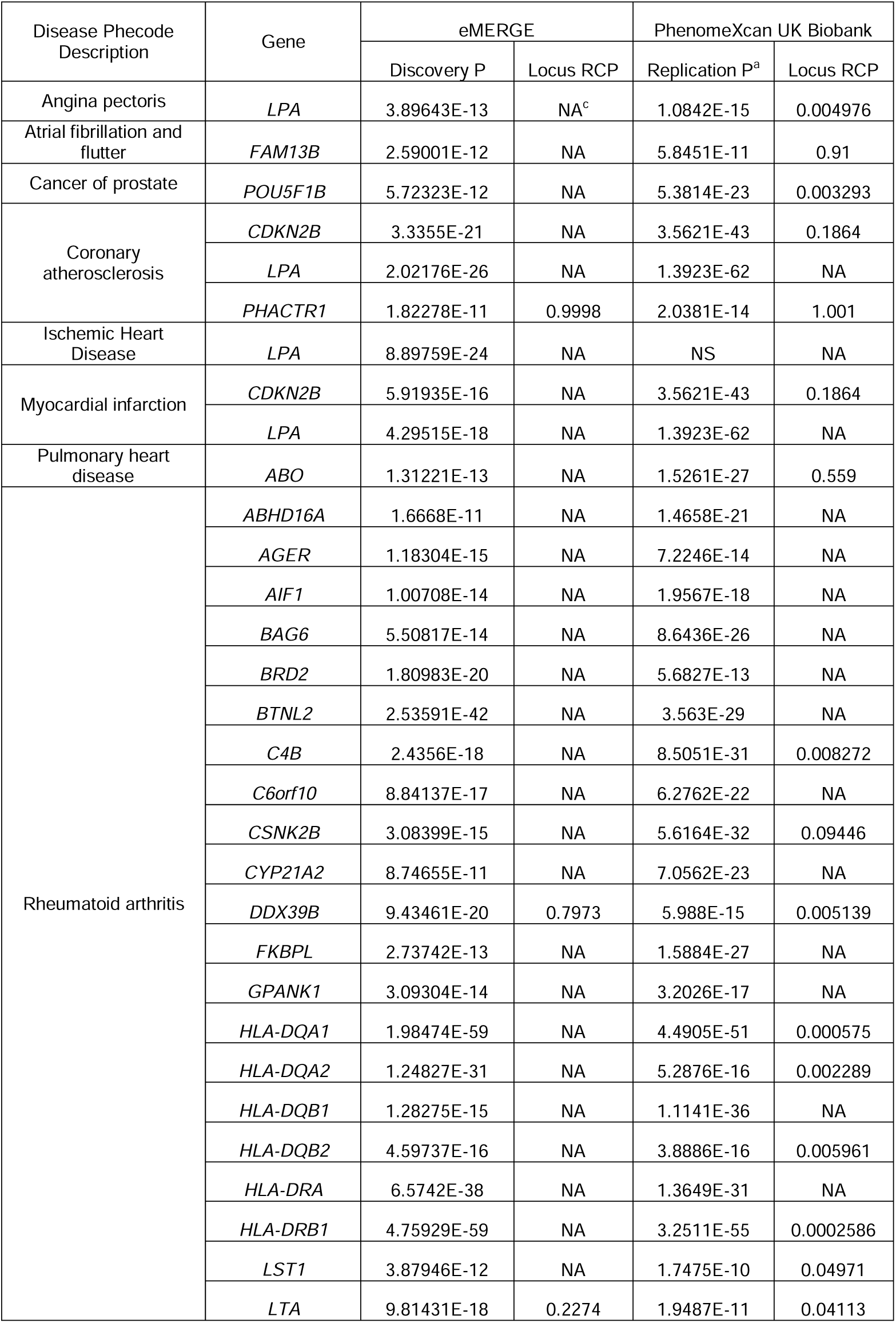
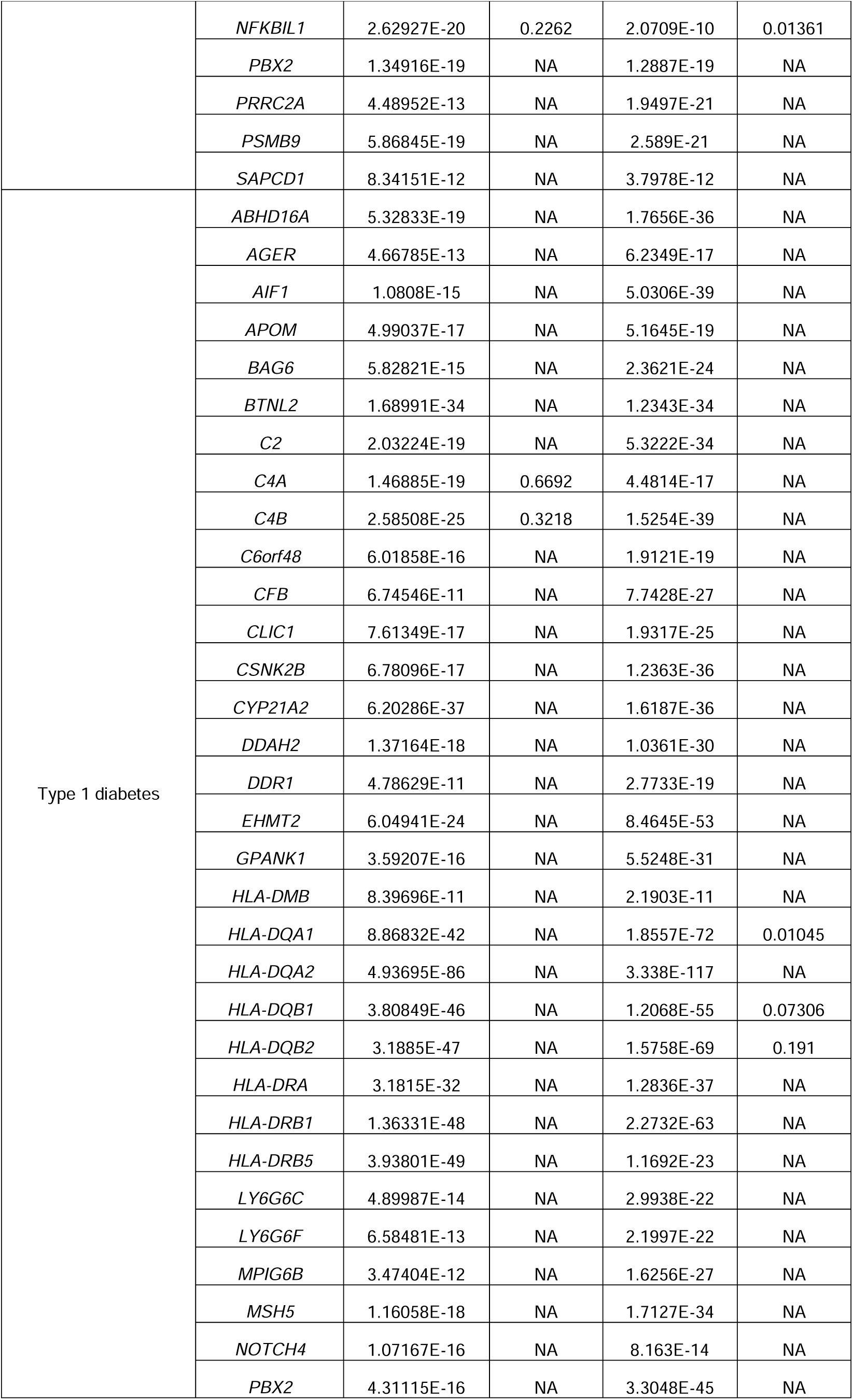
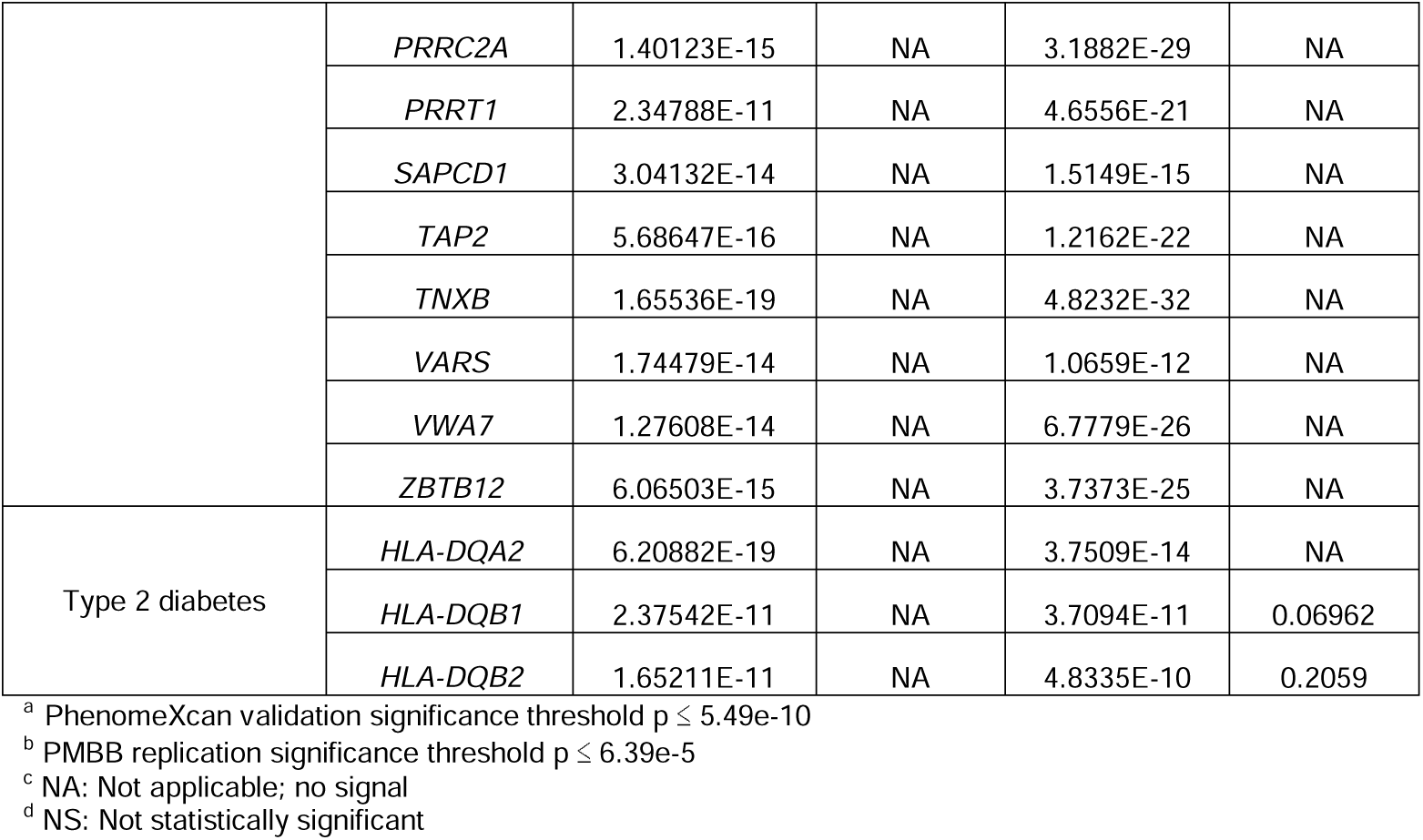
Top statistically significant genes from the eMERGE III European ancestry-specific analysis that replicated in PMBB participants of European ancestry, or were validated in PhenomeXcan UK Biobank participants of European ancestry.

We found novel associations that have not been reported elsewhere besides by the PhenomeXcan [17]. Several novel associations were further supported with a colocalization signal between eQTLs and GWAS signals in the eMERGE III (locus RCP > 0.1). One example was the association between rheumatoid arthritis and *DDX39B* (p = 9.43e-20, locus RCP = 0.797; PhenomeXcan validation p = 5.99e-15) (**Supplementary Table 3**). Another example was the association between rheumatoid arthritis and *LTA* (p = 9.81e-18, locus RCP = 0.227; PhenomeXcan validation p = 1.95e-11) (**Supplementary Table 3**). *LTA* has been associated with myocardial infarction [42] and psoriatic arthritis [43], but not rheumatoid arthritis in any previous research except in the PhenomeXcan [17].

### Gene-disease analyses for individuals of African ancestry

Genes were associated with more unique phenotypes in the eMERGE III individuals of African ancestry. In total, 397 gene-disease associations were statistically significant in the eMERGE III individuals of African ancestry, comprising 144 genes and 218 phecode-defined diseases in all 14 disease categories (**Figure 8**; **Supplementary Table 4**). Only 2 of the 390 significant associations had locus RCP > 0.1. The majority (99.2%) of the 397 gene-disease associations were only significant in the eMERGE III individuals of African ancestry. Three associations were found to be statistically significant in both European ancestry-specific analysis and cross-ancestry analysis. These included associations between *HLA-DRB5* (p = 1.56e-11) and type 1 diabetes, *TOMM40* with lipid disorders (phecode 272; p = 2.05e-13) and hyperlipidemia (phecode 272.1; p = 7.37e-13).

**Figure 8.**
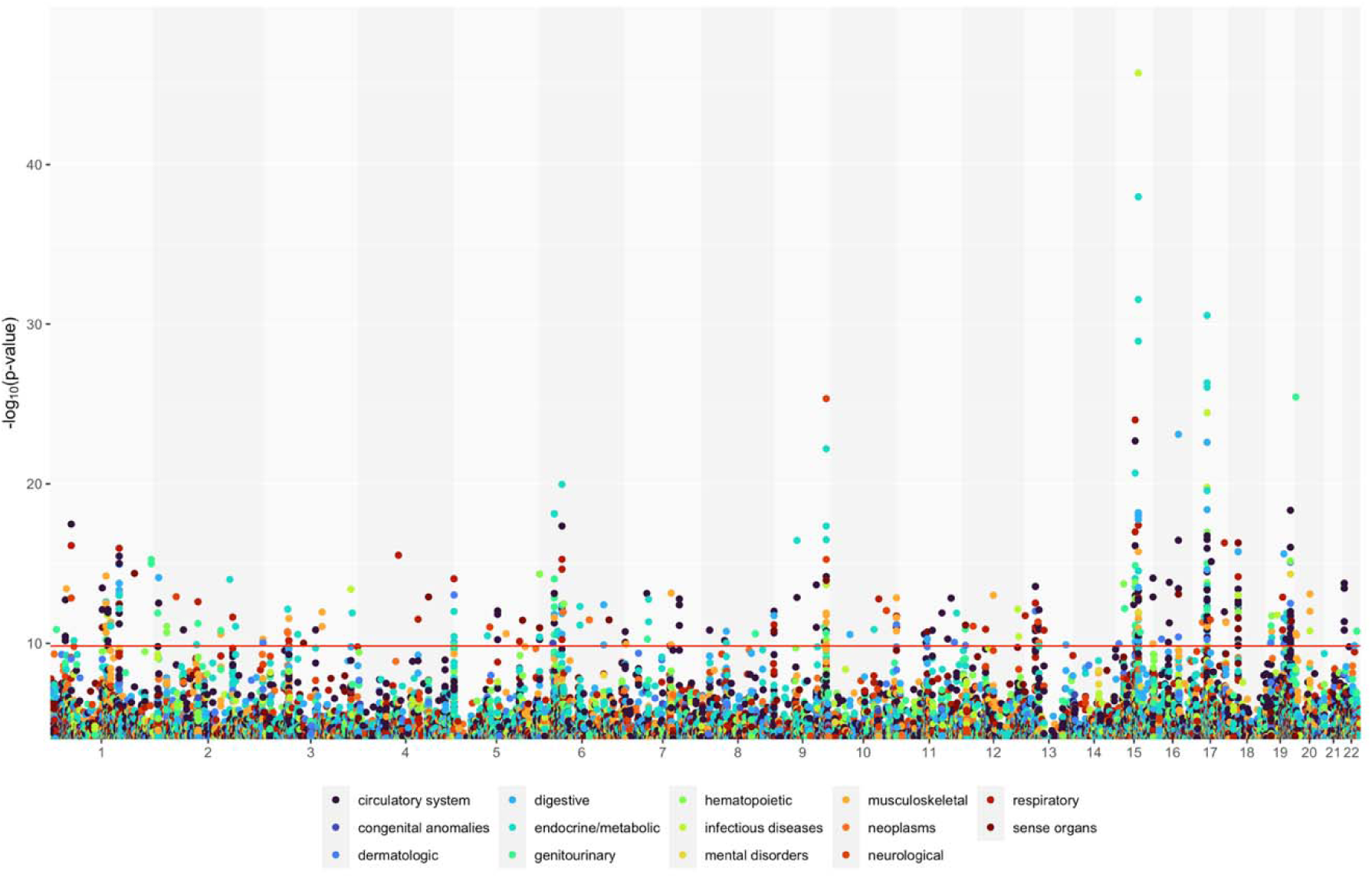
Gene-disease associations for the eMERGE III individuals of African ancestry. X-axis represented chromosome position. Y-axis was negative log10 transformed p-values. Red horizontal line was the Bonferroni significance threshold (p = 1.47e-10). Colors represented varied categories of diseases. 28 (7.0%) gene-disease associations were successfully replicated in the analogous PMBB dataset which included individuals of African ancestry with a replication significance threshold at p ≤ 6.39e-5.

### Cross-ancestry TWAS

Cross-ancestry analysis of the eMERGE III individuals identified a total of 306 statistically significant gene-disease associations (**Figure 9**). A large proportion (75.8%) of the 306 genes were also significant in the eMERGE III ancestry-specific analysis. 74 (24.2%) gene-disease associations were unique to the cross-ancestry analysis (**Supplementary Table 5**).

**Figure 9.**
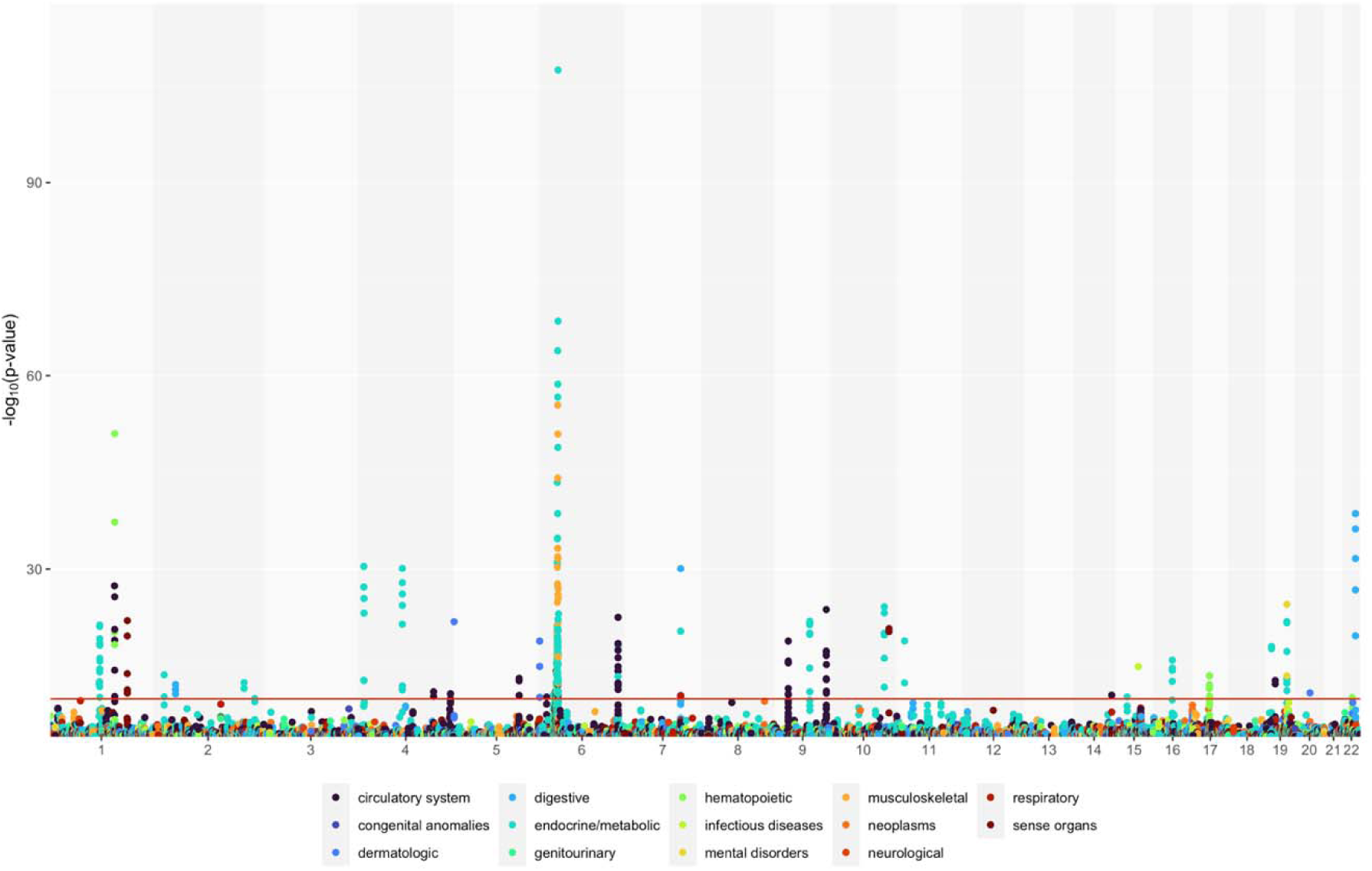
Gene-disease associations for the eMERGE III cross-ancestry analysis. X-axis represented chromosome position. Y-axis was negative log10 transformed p-values. Red horizontal line was the Bonferroni significance threshold (p = 1.47e-10). Colors represented varied categories of diseases. The majority of the cross-ancestry statistically significant associations were also found in the European ancestry-specific analysis.

We did not find replication of the eMERGE III cross-ancestry analysis in the PMBB cross-ancestry analysis; this is probably due to the limited sample size of PMBB in comparison to the eMERGE III cohort. Interestingly, several associations were validated in the PhenomeXcan UK Biobank data (**Table 3**). We found the association between *AGPAT1* and rheumatoid arthritis (p = 4.33e-17) which has been reported in previous literature besides PhenomeXcan [58]. On the other hand, we were the first besides PhenomeXcan to report the associations of type 1 diabetes to *ATP6V1G2* (p = 6.52e-20) and *TAP1* (p = 1.18e-14), and of atrial fibrillation to *RP11-325L7*.*2* (p = 7.03e-11).

**Table 2.** Top statistically significant genes from the eMERGE III African ancestry-specific analysis that replicated in the PMBB participants of African ancestry.

| Disease Phecode Description | Gene | eMERGE III<br>African Ancestry |  |
| --- | --- | --- | --- |
|  |  | Discovery P | Locus RCP |
| Acute bronchitis and bronchiolitis | *CTC-304I17.5* | 4.22E-13 | NA<sup>b</sup> |
| Acute pulmonary heart disease | *ASH1L* | 9.06E-13 | NA |
|  | *GOLGA6B* | 3.11E-11 | NA |
| Atherosclerosis | *GOLGA6B* | 1.98E-13 | NA |
| Cancer of urinary organs | *LINC00563* | 8.07E-11 | NA |
| Cardiomegaly | *RP11-49P4.7* | 1.32E-18 | NA |
| Disturbance of skin sensation | *LINC01001* | 1.28E-12 | NA |
| Dysphagia | *CTC-304I17.5* | 1.46E-11 | NA |
|  | *GOLGA6B* | 8.64E-13 | NA |
| End stage renal disease | *GOLGA6B* | 4.91E-13 | NA |
| Essential hypertension | *GOLGA6B* | 1.51E-11 | NA |
| Frequency of urination and polyuria | *TGM3* | 2.92E-23 | NA |
| Hypertension | *GOLGA6B* | 4.15E-12 | NA |
| Inflammation of the eye | *OBP2B* | 4.00E-14 | NA |
| Insomnia | *OBP2B* | 2.22E-26 | NA |
| Intestinal infection | *CTC-304I17.5* | 4.36E-12 | NA |
| Nonspecific chest pain | *NPHS2* | 3.30E-17 | NA |
|  | *RP11-552E20.4* | 1.37E-16 | NA |
| Occlusion and stenosis of precerebral arteries | *HIST1H1B* | 6.60E-12 | NA |
| Osteoarthritis; localized | *OBP2B* | 5.12E-11 | NA |
| Osteopenia or other disorder of bone and cartilage | *CTC-304I17.5* | 2.07E-12 | NA |
| Pleurisy; pleural effusion | *CTC-304I17.5* | 8.54E-11 | NA |
| Pulmonary embolism and infarction,<br>acute | *ASH1L* | 7.33E-13 | NA |
| Respiratory failure | *AC109826.1* | 5.07E-13 | NA |
| Symptoms and disorders of the joints | *CTC-304I17.5* | 3.87E-13 | NA |
| Type 1 diabetes | *HLA-DRB5* | 1.56E-11 | NA |
| Urinary tract infection | *GOLGA6B* | 4.06E-13 | NA |
| Viral hepatitis C | *CTC-304I17.5* | 1.26E-20 | NA |

<sup>a</sup> PMBB replication significance threshold $p \leq 6.39\text{e-}5$
<sup>b</sup> NA: not applicable; no signal

**Table 3.** Statistically significant genes that were exclusively identified by the eMERGE III cross-ancestry analysis and were validated by PhenomeXcan.

| Disease Phecode Description | Gene | eMERGE III<br>Cross-ancestry | PhenomeXcan<br>European Ancestry |
| --- | --- | --- | --- |
|  |  | Discovery P | Validation P<sup>a</sup> |
| Type 1 diabetes | *XXbac-BPG181B23.7* | 4.17E-12 | 8.94E-24 |
|  | *ATP6V1G2* | 6.52E-20 | 1.43E-22 |
|  | *HLA-B* | 9.57E-13 | 1.64E-20 |
|  | *MICB* | 1.46E-10 | 4.12E-17 |
|  | *TAP1* | 1.18E-14 | 3.46E-15 |
|  | *DXO* | 1.87E-17 | 7.04E-11 |
|  | *MICA* | 4.72E-11 | 9.51E-11 |
| Rheumatoid arthritis and other inflammatory | *AGPAT1* | 4.33E-17 | 1.35E-13 |
| polyarthropathies |  |  |  |
| Atrial fibrillation and flutter | *RP11-325L7.2* | 7.03E-11 | 3.14E-11 |

<sup>a</sup> PhenomeXcan validation significance threshold $p \leq 5.49\text{e-}10$

Some statistically significant gene-disease associations that we identified in the cross-ancestry analysis were proof-of-concept or well-studied associations, even though they did not replicate in the PMBB or were not validated in the PhenomeXcan (**Supplementary Table 5)**. For example, the association between *SORT1* and hypercholesterolemia (p = 4.29e-14) [59,60]. Another example was the association between *SAMM50* and chronic liver diseases (p = 3.31e-24), which has been reported in the literature in multiple East Asian ancestry groups (Chinese Han, Korean, Japan) and admixed Latino populations [61-64].

## Discussion

We investigated ancestry-specific and cross-ancestry gene-disease associations across 309 phecode-defined disease phenotypes and 22,535 genes for European ancestry, African ancestry, and a cross-ancestry analysis through meta-analysis. Given the vast volume of data, we designed a phenome-wide TWAS (gene-based) framework that integrates phenome-wide GWAS (SNP-based) summary statistics with the latest GTEx v8 eQTL data to estimate gene-disease relationships. We further supported ancestry-specific TWAS gene-disease associations with colocalization analyses to improve the precision of disease-related gene identification [33]. The discovery analyses were performed on the eMERGE III network participants. Statistically significant gene-disease associations (p < 1.47e-10) were replicated in the equivalent ancestry subset from PMBB. We further validated the results against the PhenomeXcan UK Biobank results [17], PheWAS catalog [18] or via systematic literature review. We identified a total of 782 statistically significant unique gene-disease associations in the eMERGE III network cohort. 74 (9.5%) of the 782 statistically significant gene-disease associations were exclusive to the cross-ancestry analysis. 82 (10.5%) of the 782 associations were exclusive to the eMERGE III individuals of European ancestry. 394 (50.4%) of the 782 were found only in the eMERGE III individuals of African ancestry. The remaining 232 (29.7%) gene-disease associations were found in ancestry-specific and/or cross-ancestry analyses. A good number of the 782 gene-disease associations were successfully replicated in the independent PMBB complementary analyses; some were validated by PhenomeXcan on the UK Biobank individuals of European ancestry, or by previous genomics research in the literature. Gene-disease associations that were successfully replicated in the PMBB included both proof-of-concept associations and novel gene-disease connections that are worth further investigation in additional datasets.

Gene-level analyses on individuals of African ancestry revealed a vast pool of unique gene-disease associations that the European ancestry-specific or cross-ancestry analyses did not identify. While the number of African ancestry individuals was only 19% of the number of European ancestry individuals in the eMERGE III data, African ancestry-specific analysis contributed nearly half (394 of 782; 50.4%) of the total unique gene-disease associations. 28 (7.1%) of the 397 African ancestry-specific gene-disease associations replicated in the PMBB individuals of African ancestry. In addition, the number of African ancestry-exclusive associations were five times more than those of European ancestry-exclusive associations (394 vs 82). The abundant number of associations identified in the analysis from individuals of African ancestry indicate the value of ancestry-specific genomic analyses. That said, due to the smaller sample size of the African ancestry subset of the eMERGE III dataset, it is possible that many of these associations are false positive associations. Because both the eMERGE III and PMBB African ancestry datasets are modest in sample size, we want to be careful regarding the interpretations of the association results. However, we feel it is extremely important to publish these putative associations into the public domain so that other researchers with African ancestry datasets can attempt to replicate these signals. Until the genome-wide association study datasets in individuals of African ancestry, as well as other ancestry groups, are as large as those of European ancestry, the field will be limited in identifying association signals that are robust, replicated, and believed with confidence. Our results demonstrate an overwhelming need to increase the size and availability of genome-wide datasets in individuals of African ancestry. Coupled with clinical data, ancestry-specific discoveries may fuel the application of disease risk prediction, such as polygenic risk scores (PRSs), and therapeutics management for a wider population that is currently present in the EHR-linked biobanks.

Several associations from individuals of African ancestry are worthy of pursuit for additional replication and validation as they may be excellent candidates to explore potential disease mechanisms. Some have been previously investigated but our study was the first to report some of these associations, for example, the *GATA6-AS1* connection to heart diseases. *GATA6-AS1* is known to be co-expressed with *GATA6* in several human tissues, including heart, to regulate cardiomyocyte differentiation [56]. *GATA6-AS1* is also able to interact with *LOXL2*, an epigenetic regulatory, to regulate endothelial gene expression via changes in histone methylation [57], which can have dreadful consequences, such as vascular diseases, heart diseases, etc. [65]. Some of the replicated associations were novel and have not been previously reported by other studies, for example, the connection between *NPHS2* and chest pain. The pathological connection is not clear, but *NPHS2* may moderate risks of chronic diseases, such as end-stage renal disease, which can develop chest pain as a syndrome. Indeed, heterozygous *NPHS2* mutation carriers had a high chance of later onset steroid-resistant nephrotic syndrome and progression of end-stage renal disease [66]. Chest pain is one of the common syndromes as chronic kidney disease progresses to end-stage renal disease. Overall, individuals of African ancestry from two independent EHR-linked biobanks provided a list of 29 replicated gene-disease connections for future disease mechanism studies.

While there are many interesting associations uncovered by our study, there are several limitations that are important to note. First, the sample size is certainly limited for some combinations of ancestry group and low frequency phenotypes (rare diseases). For example, we see examples where the number of cases was limited for rare diseases among individuals of African ancestry, leading to limited power to understand disease mechanisms in this specific genetic ancestry group. In addition, we were not able to replicate any of the 74 cross-ancestry-exclusive gene-disease associations in the analogous PMBB cross-ancestry analysis; this was likely due to a lower sample size and reduced power in the PMBB (N = 15,473) than that in the eMERGE III network (N = 81,576) for both ancestry-based groups. Second, we observed that several cross-ancestry associations in the eMERGE III dataset (N = 81,576) replicated in the PhenomeXcan analysis in the UK Biobank individuals of European ancestry (N = 361,194). These replicated associations do not necessarily implicate a gene-disease connection that is ancestry-specific, but instead most likely demonstrate associations that are shared by individuals of European and African ancestry. We speculate, however, that these are genes with moderate to small effect sizes and therefore they require a much greater sample size to reach statistical significance. Third, our phenome-wide TWAS framework focused only on identification of transcriptionally regulated genes behind complex diseases. Investigations of other possible disease mechanism (such as methylation, alternative splicing, or translational regulation) demands alternative methodological designs beyond TWAS. Fourth, due to the precise but conservative nature of colocalization approaches [33], we used locus RCP only as a supportive functional evidence for gene prioritization, additional to TWAS statistical significance and not a hard-line requirement. Indeed, several of our gene-disease associations did not exceed the pre-defined colocalization signal threshold (locus RCP > 0.1) but were, nonetheless, proof-of-concept associations, such as the association between *SORT1* and hypercholesterolemia (p = 4.29e-14). We defer the comparison of TWAS and colocalization approaches to future researchers.

In summary, we described a multi-ancestry gene-disease connection landscape for individuals of European and African ancestry. Our study revealed a large number of African ancestry-specific gene-disease associations and provided a resource of ancestry-specific and cross-ancestry gene-disease connections for future multi-ancestry complex disease studies. Our results demonstrate the overwhelming need for larger datasets of African ancestry genome-wide SNP data along with phenotypes across the phenome so that the gene-disease associations identified herein can be further replicated in larger, more robust datasets.

## Data Availability

All data produced in the present work are contained in the manuscript

## Notes

### Competing Interest Statement

MDR is on the scientific advisory board for Cipherome.

### Funding Statement

eMERGE Network (Phase III). This phase of the eMERGE Network was initiated and funded by the NHGRI through the following grants: U01HG8657 (Group Health Cooperative/University of Washington); U01HG8685 (Brigham and Womens Hospital); U01HG8672 (Vanderbilt University Medical Center); U01HG8666 (Cincinnati Childrens Hospital Medical Center); U01HG6379 (Mayo Clinic); U01HG8679 (Geisinger Clinic); U01HG8680 (Columbia University Health Sciences); U01HG8684 (Childrens Hospital of Philadelphia); U01HG8673 (Northwestern University); U01HG8701 (Vanderbilt University Medical Center serving as the Coordinating Center); U01HG8676 (Partners Healthcare/Broad Institute); and U01HG8664 (Baylor College of Medicine). Penn Medicine BioBank (PMBB). The PMBB is funded by the Perelman School of Medicine at the University of Pennsylvania, a gift from the Smilow family, and the National Center for Advancing Translational Sciences of the National Institutes of Health under CTSA Award Number UL1TR001878. We thank D. Birtwell, H. Williams, P. Baumann and M. Risman for informatics support regarding the PMBB. We thank the staff of the Regeneron Genetics Center for whole-exome sequencing of DNA from PMBB participants.

### Author Declarations

IRB of the University of Pennsylvania gave ethnical approval for this work.

