## Supplemental Table 1 for "Multi-ancestry gene-trait connection landscape using electronic health record (EHR) linked biobank data"

Supplementary Table 1. Case and control counts for the 309 phecode-defined complex disease phenotypes in the eMERGE III and PMBB individuals. Counts were biobank and genetic ancestry group-specific.

| Phecode | Phenotype Description | eMERGE III African ancestry case | eMERGE III  African ancestry control | eMERGE III European ancestry case | eMERGE III European ancestry control | PMBB African ancestry case | PMBB  African ancestry control | PMBB European ancestry case | PMBB European ancestry control | Disease Category |
| --- | --- | --- | --- | --- | --- | --- | --- | --- | --- | --- |
| 008 | Intestinal infection | 281 | 11180 | 1639 | 57495 | 244 | 7350 | 81 | 3499 | infectious diseases |
| 008.5 | Bacterial enteritis | 134 | 11180 | 1024 | 57495 | 170 | 7350 | 61 | 3499 | infectious diseases |
| 008.52 | Intestinal infection due to C. difficile | 111 | 11180 | 893 | 57495 | 90 | 7350 | 58 | 3499 | infectious diseases |
| 038 | Septicemia | 474 | 9753 | 3172 | 50610 | 428 | 6346 | 190 | 2968 | infectious diseases |
| 038.3 | Bacteremia | 240 | 9753 | 1361 | 50610 | 143 | 6346 | 70 | 2968 | infectious diseases |
| 053 | Herpes zoster | 172 | 8069 | 1275 | 45057 | 148 | 5569 | 55 | 3148 | infectious diseases |
| 070 | Viral hepatitis | 686 | 8069 | 1008 | 45057 | 492 | 5569 | 58 | 3148 | infectious diseases |
| 070.3 | Viral hepatitis C | 581 | 8069 | 860 | 45057 | 414 | 5569 | 51 | 3148 | infectious diseases |
| 079 | Viral infection | 751 | 8069 | 1934 | 45057 | 335 | 5569 | 153 | 3148 | infectious diseases |
| 080 | Postoperative infection | 118 | 11305 | 1446 | 56142 | 159 | 7419 | 82 | 3372 | infectious diseases |
| 081 | Infection/inflammation of internal prosthetic device; implant; and graft | 184 | 11305 | 1186 | 56142 | 150 | 7419 | 85 | 3372 | infectious diseases |
| 110 | Dermatophytosis / Dermatomycosis | 1384 | 8450 | 4205 | 48249 | 456 | 5793 | 77 | 3287 | infectious diseases |
| 110.1 | Dermatophytosis | 1296 | 8450 | 4038 | 48249 | 410 | 5793 | 71 | 3287 | infectious diseases |
| 112 | Candidiasis | 675 | 8450 | 1884 | 48249 | 512 | 5793 | 75 | 3287 | infectious diseases |
| 165 | Cancer within the respiratory system | 96 | 11999 | 1456 | 59601 | 166 | 7826 | 79 | 3578 | neoplasms |
| 165.1 | Cancer of bronchus; lung | 94 | 11999 | 1422 | 59601 | 161 | 7826 | 77 | 3578 | neoplasms |
| 173 | Neoplasm of uncertain behavior of skin | 87 | 11726 | 4443 | 47699 | 114 | 7589 | 235 | 3189 | neoplasms |
| 185 | Cancer of prostate | 234 | 11184 | 2815 | 49815 | 382 | 6906 | 118 | 2935 | neoplasms |
| 189 | Cancer of urinary organs (incl. kidney and bladder) | 152 | 11947 | 1835 | 59353 | 214 | 7770 | 89 | 3575 | neoplasms |
| 195 | Cancer, suspected or other | 161 | 10799 | 2250 | 47790 | 174 | 7203 | 63 | 3419 | neoplasms |
| 198 | Secondary malignant neoplasm | 224 | 10799 | 3243 | 47790 | 245 | 7203 | 78 | 3419 | neoplasms |
| 199 | Neoplasm of uncertain behavior | 194 | 10799 | 2629 | 47790 | 223 | 7203 | 71 | 3419 | neoplasms |
| 202 | Cancer of other lymphoid, histiocytic tissue | 107 | 11591 | 1221 | 57962 | 72 | 7716 | 57 | 3498 | neoplasms |
| 202.2 | Non-Hodgkins lymphoma | 94 | 11591 | 1134 | 57962 | 71 | 7716 | 55 | 3498 | neoplasms |
| 208 | Benign neoplasm of colon | 868 | 10340 | 8597 | 47097 | 694 | 6468 | 164 | 3328 | neoplasms |
| 216 | Benign neoplasm of skin | 239 | 11356 | 7903 | 45345 | 121 | 7551 | 178 | 3393 | neoplasms |
| 241 | Nontoxic nodular goiter | 532 | 9961 | 3158 | 44684 | 579 | 6370 | 106 | 2803 | endocrine/metabolic |
| 241.1 | Nontoxic uninodular goiter | 321 | 9961 | 2441 | 44684 | 363 | 6370 | 75 | 2803 | endocrine/metabolic |
| 241.2 | Nontoxic multinodular goiter | 337 | 9961 | 1781 | 44684 | 266 | 6370 | 52 | 2803 | endocrine/metabolic |
| 244 | Hypothyroidism | 790 | 9961 | 9720 | 44684 | 545 | 6370 | 530 | 2803 | endocrine/metabolic |
| 244.2 | Acquired hypothyroidism | 85 | 9961 | 1155 | 44684 | 196 | 6370 | 84 | 2803 | endocrine/metabolic |
| 244.4 | Hypothyroidism NOS | 706 | 9961 | 9284 | 44684 | 474 | 6370 | 512 | 2803 | endocrine/metabolic |
| 249 | Secondary diabetes mellitus | 200 | 7263 | 795 | 40273 | 439 | 3814 | 192 | 2132 | endocrine/metabolic |
| 250 | Diabetes mellitus | 3238 | 7263 | 12258 | 40273 | 2476 | 3814 | 950 | 2132 | endocrine/metabolic |
| 250.1 | Type 1 diabetes | 662 | 7263 | 2450 | 40273 | 224 | 3814 | 77 | 2132 | endocrine/metabolic |
| 250.2 | Type 2 diabetes | 3199 | 7263 | 12031 | 40273 | 2452 | 3814 | 923 | 2132 | endocrine/metabolic |
| 250.22 | Type 2 diabetes with renal manifestations | 693 | 7263 | 2611 | 40273 | 668 | 3814 | 135 | 2132 | endocrine/metabolic |
| 250.24 | Type 2 diabetes with neurological manifestations | 763 | 7263 | 2963 | 40273 | 444 | 3814 | 97 | 2132 | endocrine/metabolic |
| 250.4 | Abnormal glucose | 1122 | 7263 | 4947 | 40273 | 1758 | 3814 | 305 | 2132 | endocrine/metabolic |
| 250.41 | Impaired fasting glucose | 196 | 7263 | 1811 | 40273 | 245 | 3814 | 123 | 2132 | endocrine/metabolic |
| 250.6 | Polyneuropathy in diabetes | 412 | 7263 | 1675 | 40273 | 132 | 3814 | 67 | 2132 | endocrine/metabolic |
| 260 | Protein-calorie malnutrition | 271 | 8306 | 2026 | 44420 | 245 | 5384 | 144 | 2980 | endocrine/metabolic |
| 260.2 | severe protein-calorie malnutrition | 65 | 8306 | 407 | 44420 | 116 | 5384 | 52 | 2980 | endocrine/metabolic |
| 261 | Vitamin deficiency | 1926 | 8306 | 6965 | 44420 | 1431 | 5384 | 161 | 2980 | endocrine/metabolic |
| 261.4 | Vitamin D deficiency | 1748 | 8306 | 5583 | 44420 | 1334 | 5384 | 144 | 2980 | endocrine/metabolic |
| 269 | Proteinuria | 574 | 10749 | 1605 | 55537 | 565 | 6991 | 86 | 3445 | endocrine/metabolic |
| 270 | Disorders of protein plasma/amino-acid transport and metabolism | 268 | 10749 | 1595 | 55537 | 140 | 6991 | 58 | 3445 | endocrine/metabolic |
| 272 | Disorders of lipoid metabolism | 3943 | 7208 | 29244 | 26648 | 3297 | 4020 | 2239 | 929 | endocrine/metabolic |
| 272.1 | Hyperlipidemia | 3932 | 7208 | 29195 | 26648 | 3285 | 4020 | 2238 | 929 | endocrine/metabolic |
| 272.11 | Hypercholesterolemia | 2087 | 7208 | 14138 | 26648 | 1080 | 4020 | 715 | 929 | endocrine/metabolic |
| 272.13 | Mixed hyperlipidemia | 1113 | 7208 | 7481 | 26648 | 1325 | 4020 | 770 | 929 | endocrine/metabolic |
| 274 | Gout and other crystal arthropathies | 612 | 11299 | 3771 | 55858 | 551 | 7257 | 307 | 3247 | endocrine/metabolic |
| 274.1 | Gout | 588 | 11299 | 3394 | 55858 | 535 | 7257 | 301 | 3247 | endocrine/metabolic |
| 274.11 | Gouty arthropathy | 238 | 11299 | 1333 | 55858 | 223 | 7257 | 58 | 3247 | endocrine/metabolic |
| 275 | Disorders of mineral metabolism | 623 | 10684 | 3807 | 53605 | 735 | 6518 | 226 | 3108 | endocrine/metabolic |
| 275.3 | Disorders of magnesium metabolism | 173 | 10684 | 1004 | 53605 | 309 | 6518 | 121 | 3108 | endocrine/metabolic |
| 275.5 | Disorders of calcium/phosphorus metabolism | 326 | 10684 | 1621 | 53605 | 300 | 6518 | 84 | 3108 | endocrine/metabolic |
| 275.53 | Disorders of phosphorus metabolism | 190 | 10684 | 666 | 53605 | 222 | 6518 | 62 | 3108 | endocrine/metabolic |
| 276 | Disorders of fluid, electrolyte, and acid-base balance | 1909 | 8100 | 10584 | 40312 | 1547 | 4993 | 608 | 2211 | endocrine/metabolic |
| 276.1 | Electrolyte imbalance | 1512 | 8100 | 8303 | 40312 | 1272 | 4993 | 500 | 2211 | endocrine/metabolic |
| 276.12 | Hyposmolality and/or hyponatremia | 281 | 8100 | 2882 | 40312 | 313 | 4993 | 183 | 2211 | endocrine/metabolic |
| 276.13 | Hyperpotassemia | 643 | 8100 | 2971 | 40312 | 518 | 4993 | 230 | 2211 | endocrine/metabolic |
| 276.14 | Hypopotassemia | 891 | 8100 | 3728 | 40312 | 697 | 4993 | 207 | 2211 | endocrine/metabolic |
| 276.4 | Acid-base balance disorder | 446 | 8100 | 1693 | 40312 | 359 | 4993 | 110 | 2211 | endocrine/metabolic |
| 276.41 | Acidosis | 423 | 8100 | 1526 | 40312 | 337 | 4993 | 96 | 2211 | endocrine/metabolic |
| 276.5 | Hypovolemia | 787 | 8100 | 3933 | 40312 | 457 | 4993 | 152 | 2211 | endocrine/metabolic |
| 276.6 | Fluid overload | 313 | 8100 | 1453 | 40312 | 265 | 4993 | 138 | 2211 | endocrine/metabolic |
| 277.7 | Dysmetabolic syndrome X | 76 | 11542 | 632 | 57386 | 57 | 7549 | 59 | 3481 | endocrine/metabolic |
| 278 | Overweight, obesity and other hyperalimentation | 3748 | 7008 | 13872 | 42470 | 3057 | 3924 | 544 | 2813 | endocrine/metabolic |
| 278.1 | Obesity | 3471 | 7008 | 11782 | 42470 | 2895 | 3924 | 496 | 2813 | endocrine/metabolic |
| 278.11 | Morbid obesity | 1366 | 7008 | 5581 | 42470 | 1352 | 3924 | 114 | 2813 | endocrine/metabolic |
| 279 | Disorders involving the immune mechanism | 324 | 11140 | 3309 | 53462 | 362 | 7210 | 215 | 3380 | endocrine/metabolic |
| 280 | Iron deficiency anemias | 1233 | 6715 | 4786 | 37067 | 981 | 3916 | 220 | 2041 | hematopoietic |
| 280.1 | Iron deficiency anemias, unspecified or not due to blood loss | 1098 | 6715 | 4315 | 37067 | 903 | 3916 | 196 | 2041 | hematopoietic |
| 285.1 | Acute posthemorrhagic anemia | 266 | 6715 | 2524 | 37067 | 188 | 3916 | 120 | 2041 | hematopoietic |
| 285.2 | Anemia of chronic disease | 770 | 6715 | 2606 | 37067 | 486 | 3916 | 143 | 2041 | hematopoietic |
| 285.21 | Anemia in chronic kidney disease | 554 | 6715 | 1549 | 37067 | 353 | 3916 | 93 | 2041 | hematopoietic |
| 286 | Coagulation defects | 400 | 10210 | 2976 | 47206 | 251 | 6701 | 123 | 2636 | hematopoietic |
| 286.9 | Abnormal coagulation profile | 159 | 10210 | 800 | 47206 | 158 | 6701 | 60 | 2636 | hematopoietic |
| 287 | Purpura and other hemorrhagic conditions | 326 | 10210 | 2719 | 47206 | 381 | 6701 | 277 | 2636 | hematopoietic |
| 287.3 | Thrombocytopenia | 303 | 10210 | 2423 | 47206 | 371 | 6701 | 267 | 2636 | hematopoietic |
| 288 | Diseases of white blood cells | 357 | 9285 | 2802 | 44486 | 240 | 6277 | 110 | 3056 | hematopoietic |
| 288.1 | Decreased white blood cell count | 217 | 9285 | 1359 | 44486 | 188 | 6277 | 87 | 3056 | hematopoietic |
| 288.2 | Elevated white blood cell count | 280 | 9285 | 2109 | 44486 | 211 | 6277 | 101 | 3056 | hematopoietic |
| 289.4 | Lymphadenitis | 488 | 9285 | 3078 | 44486 | 335 | 6277 | 62 | 3056 | hematopoietic |
| 292 | Neurological disorders | 785 | 9916 | 5959 | 46691 | 576 | 6634 | 180 | 3208 | mental disorders |
| 292.3 | Memory loss | 184 | 9916 | 2094 | 46691 | 205 | 6634 | 64 | 3208 | mental disorders |
| 292.4 | Altered mental status | 434 | 9916 | 2549 | 46691 | 255 | 6634 | 82 | 3208 | mental disorders |
| 327 | Sleep disorders | 985 | 9232 | 7772 | 42278 | 946 | 4899 | 245 | 2580 | neurological |
| 327.3 | Sleep apnea | 1166 | 9232 | 7967 | 42278 | 1671 | 4899 | 666 | 2580 | neurological |
| 327.32 | Obstructive sleep apnea | 989 | 9232 | 6618 | 42278 | 1562 | 4899 | 594 | 2580 | neurological |
| 327.4 | Insomnia | 625 | 9232 | 4667 | 42278 | 599 | 4899 | 142 | 2580 | neurological |
| 327.7 | Sleep related movement disorders | 79 | 9232 | 1684 | 42278 | 110 | 4899 | 69 | 2580 | neurological |
| 340 | Migraine | 599 | 10924 | 3702 | 54584 | 488 | 6933 | 81 | 3509 | neurological |
| 345 | Epilepsy, recurrent seizures, convulsions | 629 | 9648 | 3538 | 44393 | 342 | 6577 | 73 | 3200 | neurological |
| 350.2 | Abnormality of gait | 553 | 10564 | 3382 | 52137 | 296 | 6844 | 83 | 3412 | neurological |
| 356 | Hereditary and idiopathic peripheral neuropathy | 316 | 11032 | 2797 | 53329 | 147 | 7198 | 67 | 3439 | neurological |
| 362 | Other retinal disorders | 523 | 9826 | 3731 | 50839 | 367 | 6482 | 109 | 3294 | sense organs |
| 362.2 | Degeneration of macula and posterior pole of retina | 194 | 9826 | 2620 | 50839 | 149 | 6482 | 66 | 3294 | sense organs |
| 365 | Glaucoma | 853 | 10048 | 3286 | 51518 | 733 | 6593 | 115 | 3345 | sense organs |
| 366 | Cataract | 1071 | 10351 | 9159 | 49002 | 1137 | 6337 | 170 | 3422 | sense organs |
| 366.2 | Senile cataract | 827 | 10351 | 7062 | 49002 | 1015 | 6337 | 121 | 3422 | sense organs |
| 367 | Disorders of refraction and accommodation; blindness and low vision | 854 | 10536 | 5572 | 52491 | 861 | 6369 | 97 | 3499 | sense organs |
| 368 | Visual disturbances | 619 | 10193 | 2694 | 53447 | 410 | 6681 | 78 | 3477 | sense organs |
| 371 | Inflammation of the eye | 594 | 9507 | 2581 | 49673 | 326 | 6593 | 54 | 3448 | sense organs |
| 374 | Other disorders of eyelids | 124 | 9507 | 1706 | 49673 | 128 | 6593 | 56 | 3448 | sense organs |
| 379 | Other disorders of eye | 355 | 9895 | 2654 | 50694 | 355 | 6451 | 69 | 3482 | sense organs |
| 379.2 | Disorders of vitreous body | 265 | 9895 | 2191 | 50694 | 293 | 6451 | 63 | 3482 | sense organs |
| 380.4 | Impacted cerumen | 452 | 10906 | 4017 | 52589 | 269 | 7212 | 52 | 3545 | sense organs |
| 386.9 | Dizziness and giddiness (Light-headedness and vertigo) | 1258 | 9443 | 8301 | 45974 | 1124 | 5810 | 458 | 2887 | sense organs |
| 389 | Hearing loss | 597 | 10783 | 6565 | 49354 | 452 | 7059 | 162 | 3377 | sense organs |
| 389.1 | Sensorineural hearing loss | 222 | 10783 | 4318 | 49354 | 198 | 7059 | 94 | 3377 | sense organs |
| 394 | Rheumatic disease of the heart valves | 243 | 9983 | 2879 | 44825 | 274 | 6536 | 403 | 1872 | circulatory system |
| 394.2 | Mitral valve disease | 51 | 9983 | 722 | 44825 | 102 | 6536 | 137 | 1872 | circulatory system |
| 394.7 | Disease of tricuspid valve | 156 | 9983 | 1700 | 44825 | 139 | 6536 | 187 | 1872 | circulatory system |
| 395 | Heart valve disorders | 730 | 9983 | 8109 | 44825 | 671 | 6536 | 1381 | 1872 | circulatory system |
| 395.1 | Nonrheumatic mitral valve disorders | 489 | 9983 | 5228 | 44825 | 412 | 6536 | 686 | 1872 | circulatory system |
| 395.2 | Nonrheumatic aortic valve disorders | 297 | 9983 | 4515 | 44825 | 294 | 6536 | 912 | 1872 | circulatory system |
| 401 | Hypertension | 5937 | 5159 | 31651 | 24152 | 4878 | 2563 | 2291 | 825 | circulatory system |
| 401.1 | Essential hypertension | 5878 | 5159 | 31366 | 24152 | 4779 | 2563 | 2176 | 825 | circulatory system |
| 401.2 | Hypertensive heart and/or renal disease | 1477 | 5159 | 6253 | 24152 | 1390 | 2563 | 641 | 825 | circulatory system |
| 401.21 | Hypertensive heart disease | 367 | 5159 | 1798 | 24152 | 406 | 2563 | 130 | 825 | circulatory system |
| 401.22 | Hypertensive chronic kidney disease | 1246 | 5159 | 4792 | 24152 | 1068 | 2563 | 525 | 825 | circulatory system |
| 411 | Ischemic Heart Disease | 1666 | 9124 | 15154 | 39121 | 1267 | 5998 | 1974 | 1089 | circulatory system |
| 411.1 | Unstable angina (intermediate coronary syndrome) | 252 | 9124 | 2642 | 39121 | 132 | 5998 | 155 | 1089 | circulatory system |
| 411.2 | Myocardial infarction | 580 | 9124 | 5353 | 39121 | 522 | 5998 | 681 | 1089 | circulatory system |
| 411.3 | Angina pectoris | 522 | 9124 | 4382 | 39121 | 174 | 5998 | 234 | 1089 | circulatory system |
| 411.4 | Coronary atherosclerosis | 1417 | 9124 | 13715 | 39121 | 1101 | 5998 | 1860 | 1089 | circulatory system |
| 411.8 | Other chronic ischemic heart disease, unspecified | 319 | 9124 | 5002 | 39121 | 267 | 5998 | 605 | 1089 | circulatory system |
| 414 | Other forms of chronic heart disease | 319 | 9124 | 2579 | 39121 | 330 | 5998 | 383 | 1089 | circulatory system |
| 415 | Pulmonary heart disease | 691 | 10047 | 3941 | 48030 | 801 | 6575 | 564 | 2697 | circulatory system |
| 415.1 | Acute pulmonary heart disease | 275 | 10047 | 1857 | 48030 | 307 | 6575 | 90 | 2697 | circulatory system |
| 415.11 | Pulmonary embolism and infarction, acute | 273 | 10047 | 1837 | 48030 | 306 | 6575 | 88 | 2697 | circulatory system |
| 415.2 | Chronic pulmonary heart disease | 482 | 10047 | 2367 | 48030 | 585 | 6575 | 506 | 2697 | circulatory system |
| 415.21 | Primary pulmonary hypertension | 140 | 10047 | 694 | 48030 | 147 | 6575 | 91 | 2697 | circulatory system |
| 416 | Cardiomegaly | 739 | 10047 | 5259 | 48030 | 178 | 6575 | 55 | 2697 | circulatory system |
| 418 | Nonspecific chest pain | 3347 | 6551 | 17765 | 33317 | 2266 | 4413 | 644 | 2594 | circulatory system |
| 418.1 | Precordial pain | 321 | 6551 | 1160 | 33317 | 207 | 4413 | 72 | 2594 | circulatory system |
| 420 | Carditis | 231 | 10928 | 1609 | 53488 | 194 | 6701 | 158 | 2361 | circulatory system |
| 420.2 | Pericarditis | 179 | 10928 | 1015 | 53488 | 128 | 6701 | 88 | 2361 | circulatory system |
| 420.3 | Endocarditis | 54 | 10928 | 584 | 53488 | 55 | 6701 | 61 | 2361 | circulatory system |
| 425 | Cardiomyopathy | 546 | 10928 | 3761 | 53488 | 836 | 6701 | 789 | 2361 | circulatory system |
| 425.1 | Primary/intrinsic cardiomyopathies | 506 | 10928 | 3531 | 53488 | 796 | 6701 | 696 | 2361 | circulatory system |
| 425.2 | Secondary/extrinsic cardiomyopathies | 170 | 10928 | 1182 | 53488 | 237 | 6701 | 283 | 2361 | circulatory system |
| 426 | Cardiac conduction disorders | 841 | 7293 | 8424 | 29122 | 871 | 4144 | 689 | 985 | circulatory system |
| 426.2 | Atrioventricular [AV] block | 172 | 7293 | 2935 | 29122 | 188 | 4144 | 292 | 985 | circulatory system |
| 426.21 | First degree AV block | 80 | 7293 | 1861 | 29122 | 88 | 4144 | 84 | 985 | circulatory system |
| 426.24 | Atrioventricular block, complete | 62 | 7293 | 892 | 29122 | 78 | 4144 | 197 | 985 | circulatory system |
| 426.3 | Bundle branch block | 175 | 7293 | 2705 | 29122 | 236 | 4144 | 337 | 985 | circulatory system |
| 426.31 | Right bundle branch block | 92 | 7293 | 1292 | 29122 | 100 | 4144 | 125 | 985 | circulatory system |
| 426.32 | Left bundle branch block | 88 | 7293 | 1454 | 29122 | 130 | 4144 | 219 | 985 | circulatory system |
| 427 | Cardiac dysrhythmias | 2259 | 7293 | 19837 | 29122 | 2008 | 4144 | 2032 | 985 | circulatory system |
| 427.1 | Paroxysmal tachycardia, unspecified | 315 | 7293 | 3353 | 29122 | 585 | 4144 | 762 | 985 | circulatory system |
| 427.11 | Paroxysmal supraventricular tachycardia | 133 | 7293 | 1638 | 29122 | 256 | 4144 | 210 | 985 | circulatory system |
| 427.12 | Paroxysmal ventricular tachycardia | 213 | 7293 | 2032 | 29122 | 424 | 4144 | 654 | 985 | circulatory system |
| 427.2 | Atrial fibrillation and flutter | 591 | 7293 | 8812 | 29122 | 729 | 4144 | 1491 | 985 | circulatory system |
| 427.21 | Atrial fibrillation | 567 | 7293 | 8621 | 29122 | 698 | 4144 | 1437 | 985 | circulatory system |
| 427.22 | Atrial flutter | 165 | 7293 | 2285 | 29122 | 229 | 4144 | 471 | 985 | circulatory system |
| 427.4 | Cardiac arrest and ventricular fibrillation | 54 | 7293 | 587 | 29122 | 90 | 4144 | 88 | 985 | circulatory system |
| 427.5 | Arrhythmia (cardiac) NOS | 808 | 7293 | 7739 | 29122 | 96 | 4144 | 96 | 985 | circulatory system |
| 427.6 | Premature beats | 137 | 7293 | 2453 | 29122 | 266 | 4144 | 358 | 985 | circulatory system |
| 427.8 | Sinoatrial node dysfunction (Bradycardia) | 126 | 7293 | 1778 | 29122 | 100 | 4144 | 285 | 985 | circulatory system |
| 427.9 | Palpitations | 771 | 7293 | 6092 | 29122 | 752 | 4144 | 596 | 985 | circulatory system |
| 428 | Congestive heart failure; nonhypertensive | 1219 | 9591 | 7767 | 44594 | 1295 | 5860 | 1253 | 1288 | circulatory system |
| 428.1 | Congestive heart failure (CHF) NOS | 1024 | 9591 | 6071 | 44594 | 861 | 5860 | 1017 | 1288 | circulatory system |
| 428.2 | Heart failure NOS | 510 | 9591 | 3584 | 44594 | 617 | 5860 | 318 | 1288 | circulatory system |
| 428.3 | Heart failure with reduced EF [Systolic or combined heart failure] | 440 | 9591 | 2774 | 44594 | 683 | 5860 | 605 | 1288 | circulatory system |
| 428.4 | Heart failure with preserved EF [Diastolic heart failure] | 385 | 9591 | 2434 | 44594 | 489 | 5860 | 285 | 1288 | circulatory system |
| 429 | Ill-defined descriptions and complications of heart disease | 331 | 9591 | 3428 | 44594 | 327 | 5860 | 305 | 1288 | circulatory system |
| 429.1 | Heart transplant/surgery | 89 | 9591 | 594 | 44594 | 95 | 5860 | 253 | 1288 | circulatory system |
| 429.2 | Abnormal function study of cardiovascular system | 157 | 9591 | 1024 | 44594 | 135 | 5860 | 158 | 1288 | circulatory system |
| 429.3 | Symptoms involving cardiovascular system | 151 | 9591 | 2168 | 44594 | 194 | 5860 | 105 | 1288 | circulatory system |
| 433 | Cerebrovascular disease | 1007 | 10325 | 8055 | 47131 | 774 | 6828 | 568 | 2810 | circulatory system |
| 433.1 | Occlusion and stenosis of precerebral arteries | 272 | 10325 | 4012 | 47131 | 165 | 6828 | 289 | 2810 | circulatory system |
| 433.2 | Occlusion of cerebral arteries | 465 | 10325 | 2663 | 47131 | 420 | 6828 | 172 | 2810 | circulatory system |
| 433.21 | Cerebral artery occlusion, with cerebral infarction | 447 | 10325 | 2524 | 47131 | 419 | 6828 | 171 | 2810 | circulatory system |
| 433.3 | Cerebral ischemia | 473 | 10325 | 3305 | 47131 | 196 | 6828 | 135 | 2810 | circulatory system |
| 433.31 | Transient cerebral ischemia | 427 | 10325 | 3128 | 47131 | 190 | 6828 | 132 | 2810 | circulatory system |
| 433.6 | Acute, but ill-defined cerebrovascular disease | 146 | 10325 | 1088 | 47131 | 65 | 6828 | 54 | 2810 | circulatory system |
| 440 | Atherosclerosis | 389 | 10021 | 4993 | 42478 | 267 | 6550 | 171 | 2384 | circulatory system |
| 440.2 | Atherosclerosis of the extremities | 273 | 10021 | 3046 | 42478 | 202 | 6550 | 82 | 2384 | circulatory system |
| 440.22 | Atherosclerosis of native arteries of the extremities with intermittent claudication | 155 | 10021 | 2161 | 42478 | 138 | 6550 | 60 | 2384 | circulatory system |
| 442.1 | Aortic aneurysm | 109 | 10021 | 3111 | 42478 | 199 | 6550 | 363 | 2384 | circulatory system |
| 442.11 | Abdominal aortic aneurysm | 58 | 10021 | 2053 | 42478 | 79 | 6550 | 118 | 2384 | circulatory system |
| 443 | Peripheral vascular disease | 636 | 10021 | 5389 | 42478 | 512 | 6550 | 378 | 2384 | circulatory system |
| 443.9 | Peripheral vascular disease, unspecified | 554 | 10021 | 4448 | 42478 | 468 | 6550 | 353 | 2384 | circulatory system |
| 444 | Arterial embolism and thrombosis | 69 | 10021 | 900 | 42478 | 136 | 6550 | 50 | 2384 | circulatory system |
| 452 | Other venous embolism and thrombosis | 592 | 8265 | 3816 | 36660 | 411 | 5862 | 169 | 2921 | circulatory system |
| 452.2 | Deep vein thrombosis [DVT] | 355 | 8265 | 2131 | 36660 | 347 | 5862 | 140 | 2921 | circulatory system |
| 454 | Varicose veins | 193 | 8265 | 2385 | 36660 | 146 | 5862 | 55 | 2921 | circulatory system |
| 455 | Hemorrhoids | 516 | 8265 | 3252 | 36660 | 255 | 5862 | 64 | 2921 | circulatory system |
| 456 | Chronic venous insufficiency [CVI] | 247 | 8265 | 1844 | 36660 | 133 | 5862 | 65 | 2921 | circulatory system |
| 458 | Hypotension | 505 | 10150 | 4432 | 46909 | 362 | 6717 | 209 | 2934 | circulatory system |
| 458.9 | Hypotension NOS | 420 | 10150 | 3241 | 46909 | 272 | 6717 | 139 | 2934 | circulatory system |
| 464 | Acute sinusitis | 508 | 6967 | 4292 | 40710 | 274 | 5187 | 77 | 3279 | respiratory |
| 465 | Acute upper respiratory infections of multiple or unspecified sites | 2596 | 6967 | 9159 | 40710 | 1207 | 5187 | 152 | 3279 | respiratory |
| 472 | Chronic pharyngitis and nasopharyngitis | 279 | 6975 | 2169 | 38344 | 302 | 4653 | 64 | 2976 | respiratory |
| 473 | Diseases of the larynx and vocal cords | 240 | 6975 | 1916 | 38344 | 235 | 4653 | 77 | 2976 | respiratory |
| 473.4 | Voice disturbance | 169 | 6975 | 1491 | 38344 | 178 | 4653 | 60 | 2976 | respiratory |
| 475 | Chronic sinusitis | 488 | 6975 | 4029 | 38344 | 472 | 4653 | 142 | 2976 | respiratory |
| 477 | Epistaxis or throat hemorrhage | 235 | 6975 | 1105 | 38344 | 163 | 4653 | 60 | 2976 | respiratory |
| 480 | Pneumonia | 924 | 9101 | 6699 | 44160 | 600 | 6143 | 296 | 2985 | respiratory |
| 480.1 | Bacterial pneumonia | 134 | 9101 | 1288 | 44160 | 85 | 6143 | 80 | 2985 | respiratory |
| 483 | Acute bronchitis and bronchiolitis | 374 | 9101 | 3656 | 44160 | 237 | 6143 | 120 | 2985 | respiratory |
| 496 | Chronic airway obstruction | 690 | 8086 | 5887 | 42542 | 753 | 5389 | 509 | 2614 | respiratory |
| 496.1 | Emphysema | 162 | 8086 | 1695 | 42542 | 275 | 5389 | 123 | 2614 | respiratory |
| 496.2 | Chronic bronchitis | 240 | 8086 | 1621 | 42542 | 285 | 5389 | 84 | 2614 | respiratory |
| 502 | Postinflammatory pulmonary fibrosis | 142 | 9374 | 1723 | 40673 | 81 | 6470 | 130 | 2554 | respiratory |
| 504 | Other alveolar and parietoalveolar pneumonopathy | 72 | 9374 | 678 | 40673 | 99 | 6470 | 133 | 2554 | respiratory |
| 507 | Pleurisy; pleural effusion | 493 | 9374 | 5943 | 40673 | 176 | 6470 | 175 | 2554 | respiratory |
| 508 | Pulmonary collapse; interstitial and compensatory emphysema | 727 | 9374 | 6136 | 40673 | 112 | 6470 | 99 | 2554 | respiratory |
| 509 | Respiratory failure, insufficiency, arrest | 432 | 9374 | 3180 | 40673 | 286 | 6470 | 202 | 2554 | respiratory |
| 509.1 | Respiratory failure | 329 | 9374 | 2463 | 40673 | 267 | 6470 | 181 | 2554 | respiratory |
| 513 | Respiratory abnormalities | 193 | 11361 | 908 | 57665 | 153 | 7507 | 71 | 3504 | respiratory |
| 514 | Abnormal findings examination of lungs | 671 | 10645 | 6549 | 48119 | 350 | 6895 | 151 | 3241 | respiratory |
| 514.2 | Solitary pulmonary nodule | 187 | 10645 | 2270 | 48119 | 397 | 6895 | 145 | 3241 | respiratory |
| 519 | Other diseases of respiratory system, not elsewhere classified | 194 | 11089 | 2056 | 54853 | 161 | 7500 | 90 | 3419 | respiratory |
| 519.9 | Symptoms involving respiratory system and other chest symptoms | 120 | 11089 | 1481 | 54853 | 113 | 7500 | 54 | 3419 | respiratory |
| 530 | Diseases of esophagus | 2636 | 7673 | 16588 | 34622 | 2405 | 4386 | 863 | 2264 | digestive |
| 530.1 | Esophagitis, GERD and related diseases | 2584 | 7673 | 16044 | 34622 | 2349 | 4386 | 824 | 2264 | digestive |
| 530.11 | GERD | 2532 | 7673 | 15047 | 34622 | 2282 | 4386 | 790 | 2264 | digestive |
| 531 | Peptic ulcer (excl. esophageal) | 224 | 11580 | 1561 | 57911 | 155 | 7661 | 56 | 3541 | digestive |
| 532 | Dysphagia | 535 | 7673 | 4673 | 34622 | 499 | 4386 | 156 | 2264 | digestive |
| 536 | Disorders of function of stomach | 403 | 9996 | 2122 | 49832 | 224 | 6495 | 64 | 3356 | digestive |
| 550 | Abdominal hernia | 676 | 10635 | 6743 | 49494 | 688 | 6762 | 263 | 3212 | digestive |
| 550.1 | Inguinal hernia | 119 | 10635 | 2009 | 49494 | 148 | 6762 | 84 | 3212 | digestive |
| 550.2 | Diaphragmatic hernia | 302 | 10635 | 3225 | 49494 | 193 | 6762 | 100 | 3212 | digestive |
| 550.6 | Incisional hernia | 80 | 10635 | 991 | 49494 | 150 | 6762 | 51 | 3212 | digestive |
| 558 | Noninfectious gastroenteritis | 436 | 8744 | 3191 | 36986 | 220 | 5505 | 54 | 2871 | digestive |
| 560 | Intestinal obstruction without mention of hernia | 201 | 8744 | 1858 | 36986 | 157 | 5505 | 57 | 2871 | digestive |
| 561 | Symptoms involving digestive system | 124 | 8744 | 1466 | 36986 | 258 | 5505 | 60 | 2871 | digestive |
| 562 | Diverticulosis and diverticulitis | 683 | 8744 | 7298 | 36986 | 434 | 5505 | 208 | 2871 | digestive |
| 562.1 | Diverticulosis | 647 | 8744 | 6831 | 36986 | 373 | 5505 | 170 | 2871 | digestive |
| 562.2 | Diverticulitis | 123 | 8744 | 1398 | 36986 | 117 | 5505 | 58 | 2871 | digestive |
| 564 | Functional digestive disorders | 218 | 8744 | 3027 | 36986 | 141 | 5505 | 55 | 2871 | digestive |
| 564.1 | Irritable Bowel Syndrome | 165 | 8744 | 2396 | 36986 | 125 | 5505 | 53 | 2871 | digestive |
| 571 | Chronic liver disease and cirrhosis | 405 | 9653 | 3674 | 45302 | 432 | 6351 | 167 | 3013 | digestive |
| 571.5 | Other chronic nonalcoholic liver disease | 381 | 9653 | 3459 | 45302 | 415 | 6351 | 144 | 3013 | digestive |
| 571.51 | Cirrhosis of liver without mention of alcohol | 212 | 9653 | 1025 | 45302 | 263 | 6351 | 100 | 3013 | digestive |
| 571.8 | Liver abscess and sequelae of chronic liver disease | 80 | 9653 | 660 | 45302 | 80 | 6351 | 57 | 3013 | digestive |
| 572 | Ascites (non malignant) | 171 | 9653 | 1236 | 45302 | 115 | 6351 | 73 | 3013 | digestive |
| 573 | Other disorders of liver | 240 | 9653 | 2524 | 45302 | 146 | 6351 | 63 | 3013 | digestive |
| 573.6 | Nonspecific elevation of levels of transaminase or lactic acid dehydrogenase [LDH] | 161 | 9653 | 1054 | 45302 | 146 | 6351 | 59 | 3013 | digestive |
| 573.9 | Abnormal serum enzyme levels | 216 | 9653 | 1391 | 45302 | 247 | 6351 | 52 | 3013 | digestive |
| 574 | Cholelithiasis and cholecystitis | 363 | 11119 | 3105 | 53927 | 288 | 7368 | 84 | 3486 | digestive |
| 574.1 | Cholelithiasis | 323 | 11119 | 2640 | 53927 | 246 | 7368 | 58 | 3486 | digestive |
| 577 | Diseases of pancreas | 249 | 11658 | 1795 | 58743 | 197 | 7706 | 63 | 3580 | digestive |
| 578 | Gastrointestinal hemorrhage | 672 | 9582 | 5275 | 45554 | 616 | 6389 | 186 | 3272 | digestive |
| 578.8 | Hemorrhage of rectum and anus | 302 | 9582 | 1991 | 45554 | 225 | 6389 | 51 | 3272 | digestive |
| 578.9 | Hemorrhage of gastrointestinal tract | 284 | 9582 | 2422 | 45554 | 217 | 6389 | 114 | 3272 | digestive |
| 585 | Renal failure | 2154 | 8502 | 9970 | 41467 | 1993 | 4915 | 955 | 2146 | genitourinary |
| 585.1 | Acute renal failure | 1182 | 8502 | 5336 | 41467 | 1191 | 4915 | 538 | 2146 | genitourinary |
| 585.2 | Renal failure NOS | 357 | 8502 | 1456 | 41467 | 179 | 4915 | 50 | 2146 | genitourinary |
| 585.3 | Chronic renal failure [CKD] | 1744 | 8502 | 7737 | 41467 | 1559 | 4915 | 796 | 2146 | genitourinary |
| 585.32 | End stage renal disease | 755 | 8502 | 1842 | 41467 | 609 | 4915 | 136 | 2146 | genitourinary |
| 585.33 | Chronic Kidney Disease, Stage III | 894 | 8502 | 4812 | 41467 | 818 | 4915 | 336 | 2146 | genitourinary |
| 585.34 | Chronic Kidney Disease, Stage IV | 458 | 8502 | 1855 | 41467 | 377 | 4915 | 123 | 2146 | genitourinary |
| 585.4 | Chronic kidney disease, Stage I or II | 328 | 8502 | 1307 | 41467 | 287 | 4915 | 69 | 2146 | genitourinary |
| 588 | Disorders resulting from impaired renal function | 448 | 8502 | 1105 | 41467 | 365 | 4915 | 66 | 2146 | genitourinary |
| 588.2 | Secondary hyperparathyroidism (of renal origin) | 276 | 8502 | 593 | 41467 | 332 | 4915 | 54 | 2146 | genitourinary |
| 591 | Urinary tract infection | 1675 | 8259 | 10016 | 39711 | 1044 | 5284 | 241 | 2990 | genitourinary |
| 593 | Hematuria | 534 | 8259 | 5044 | 39711 | 570 | 5284 | 140 | 2990 | genitourinary |
| 594 | Urinary calculus | 292 | 11299 | 3445 | 54788 | 232 | 7557 | 114 | 3483 | genitourinary |
| 594.1 | Calculus of kidney | 267 | 11299 | 3053 | 54788 | 206 | 7557 | 107 | 3483 | genitourinary |
| 599.2 | Retention of urine | 223 | 7971 | 3344 | 39277 | 253 | 4920 | 137 | 2985 | genitourinary |
| 599.3 | Dysuria | 860 | 7971 | 3581 | 39277 | 477 | 4920 | 57 | 2985 | genitourinary |
| 599.4 | Urinary incontinence | 675 | 7971 | 4080 | 39277 | 528 | 4920 | 80 | 2985 | genitourinary |
| 599.5 | Frequency of urination and polyuria | 792 | 7971 | 4965 | 39277 | 783 | 4920 | 129 | 2985 | genitourinary |
| 600 | Hyperplasia of prostate | 410 | 11184 | 6077 | 49815 | 471 | 6906 | 378 | 2935 | genitourinary |
| 605 | Erectile dysfunction [ED] | 571 | 10637 | 3372 | 48494 | 710 | 6535 | 148 | 2939 | genitourinary |
| 681 | Superficial cellulitis and abscess | 1019 | 9018 | 6125 | 46342 | 726 | 6223 | 189 | 3231 | dermatologic |
| 681.5 | Cellulitis and abscess of leg, except foot | 276 | 9018 | 2223 | 46342 | 208 | 6223 | 78 | 3231 | dermatologic |
| 687.1 | Rash and other nonspecific skin eruption | 965 | 8609 | 4964 | 42075 | 740 | 5009 | 99 | 3230 | dermatologic |
| 687.4 | Disturbance of skin sensation | 807 | 8609 | 5089 | 42075 | 975 | 5009 | 114 | 3230 | dermatologic |
| 694 | Dyschromia and Vitiligo | 212 | 9760 | 4189 | 44137 | 161 | 6152 | 57 | 3255 | dermatologic |
| 695 | Erythematous conditions | 554 | 9760 | 4210 | 44137 | 464 | 6152 | 99 | 3255 | dermatologic |
| 701.2 | Scar conditions and fibrosis of skin | 97 | 10363 | 1780 | 47831 | 60 | 7108 | 72 | 3481 | dermatologic |
| 702 | Degenerative skin conditions and other dermatoses | 158 | 10050 | 11479 | 39497 | 85 | 6892 | 267 | 3250 | dermatologic |
| 702.2 | Seborrheic keratosis | 113 | 11630 | 8830 | 44485 | 80 | 7701 | 167 | 3316 | dermatologic |
| 706 | Diseases of sebaceous glands | 1228 | 9318 | 5651 | 48182 | 463 | 6441 | 77 | 3423 | dermatologic |
| 707 | Chronic ulcer of skin | 417 | 11284 | 2674 | 56602 | 228 | 7562 | 97 | 3458 | dermatologic |
| 707.1 | Decubitus ulcer | 144 | 11284 | 804 | 56602 | 52 | 7562 | 51 | 3458 | dermatologic |
| 714 | Rheumatoid arthritis and other inflammatory polyarthropathies | 363 | 9887 | 3303 | 46912 | 241 | 6631 | 61 | 3274 | musculoskeletal |
| 714.1 | Rheumatoid arthritis | 308 | 9887 | 2541 | 46912 | 201 | 6631 | 50 | 3274 | musculoskeletal |
| 716 | Other arthropathies | 669 | 9785 | 3763 | 46831 | 451 | 6546 | 106 | 3276 | musculoskeletal |
| 720 | Spinal stenosis | 493 | 9598 | 4606 | 40201 | 420 | 6121 | 124 | 3183 | musculoskeletal |
| 720.1 | Spinal stenosis of lumbar region | 354 | 9598 | 3647 | 40201 | 305 | 6121 | 83 | 3183 | musculoskeletal |
| 721 | Spondylosis and allied disorders | 696 | 9598 | 7316 | 40201 | 598 | 6121 | 122 | 3183 | musculoskeletal |
| 721.1 | Spondylosis without myelopathy | 662 | 9598 | 7114 | 40201 | 527 | 6121 | 112 | 3183 | musculoskeletal |
| 722 | Intervertebral disc disorders | 797 | 9598 | 7458 | 40201 | 760 | 6121 | 145 | 3183 | musculoskeletal |
| 722.1 | Displacement of intervertebral disc | 356 | 9598 | 3010 | 40201 | 297 | 6121 | 58 | 3183 | musculoskeletal |
| 722.6 | Degeneration of intervertebral disc | 553 | 9598 | 5597 | 40201 | 508 | 6121 | 83 | 3183 | musculoskeletal |
| 726 | Peripheral enthesopathies and allied syndromes | 1014 | 8786 | 10210 | 36318 | 857 | 5166 | 136 | 3194 | musculoskeletal |
| 726.1 | Enthesopathy | 532 | 8786 | 5746 | 36318 | 410 | 5166 | 57 | 3194 | musculoskeletal |
| 740 | Osteoarthrosis | 1933 | 9035 | 15971 | 36993 | 1820 | 5352 | 386 | 3027 | musculoskeletal |
| 740.1 | Osteoarthritis; localized | 1066 | 9035 | 11461 | 36993 | 1532 | 5352 | 258 | 3027 | musculoskeletal |
| 740.11 | Osteoarthrosis, localized, primary | 900 | 9035 | 7977 | 36993 | 1339 | 5352 | 214 | 3027 | musculoskeletal |
| 740.2 | Osteoarthrosis, generalized | 281 | 9035 | 2587 | 36993 | 343 | 5352 | 56 | 3027 | musculoskeletal |
| 740.9 | Osteoarthrosis NOS | 1397 | 9035 | 9781 | 36993 | 626 | 5352 | 161 | 3027 | musculoskeletal |
| 741 | Symptoms and disorders of the joints | 894 | 9645 | 6451 | 45245 | 436 | 6678 | 55 | 3513 | musculoskeletal |
| 743 | Osteoporosis, osteopenia and pathological fracture | 1139 | 10012 | 11990 | 43175 | 610 | 7069 | 303 | 3231 | musculoskeletal |
| 743.1 | Osteoporosis | 644 | 10012 | 6732 | 43175 | 227 | 7069 | 172 | 3231 | musculoskeletal |
| 743.11 | Osteoporosis NOS | 512 | 10012 | 5574 | 43175 | 129 | 7069 | 136 | 3231 | musculoskeletal |
| 743.12 | Senile osteoporosis | 194 | 10012 | 3017 | 43175 | 140 | 7069 | 73 | 3231 | musculoskeletal |
| 743.9 | Osteopenia or other disorder of bone and cartilage | 694 | 10012 | 8262 | 43175 | 434 | 7069 | 166 | 3231 | musculoskeletal |
| 747 | Cardiac and circulatory congenital anomalies | 149 | 11761 | 2167 | 57327 | 162 | 7723 | 368 | 3162 | congenital anomalies |
| 747.1 | Cardiac congenital anomalies | 115 | 11761 | 1871 | 57327 | 114 | 7723 | 352 | 3162 | congenital anomalies |
| 747.11 | Cardiac shunt/ heart septal defect | 57 | 11761 | 1037 | 57327 | 62 | 7723 | 173 | 3162 | congenital anomalies |
| 796 | Elevated prostate specific antigen [PSA] | 167 | 11184 | 2175 | 49815 | 324 | 6906 | 118 | 2935 | genitourinary |
