## Supplemental Table 2 for "Multi-ancestry gene-trait connection landscape using electronic health record (EHR) linked biobank data"

Supplementary Table 2. Phecode mappings to PhenomeXcan UK Biobank 3-digit ICD-10 codes

| PhenomeXcan  3-digit ICD-10 | PhenomeXcan  3-digit ICD-10 Description | Phecode | Phecode  Description |
| --- | --- | --- | --- |
| A09 | Diarrhea and gastroenteritis of infectious origin | 008 | Intestinal infection |
| A01 | Typhoid and paratyphoid fevers | 008.5 | Bacterial enteritis |
| A02 | Other salmonella infections | 008.5 | Bacterial enteritis |
| A03 | Shigellosis | 008.5 | Bacterial enteritis |
| A04 | Other bacterial intestinal infections | 008.5 | Bacterial enteritis |
| A05 | Other bacterial foodborne intoxications, not elsewhere classified | 008.5 | Bacterial enteritis |
| A41 | Other septicemia | 038 | Septicemia |
| A42 | Actinomycosis | 038 | Septicemia |
| B00 | Herpesviral [herpes simplex] infections | 038 | Septicemia |
| B02 | Zoster [herpes zoster] | 053 | Herpes zoster |
| B18 | Chronic viral hepatitis | 070 | Viral hepatitis |
| B34 | Viral infection of unspecified site | 079 | Viral infection |
| H81 | Disorders of vestibular function | 079 | Viral infection |
| T81 | Complications of procedures, not elsewhere classified | 080 | Postoperative infection |
| T88 | Other complications of surgical and medical care, not elsewhere classified | 080 | Postoperative infection |
| T82 | Complications of cardiac and vascular prosthetic devices, implants, and grafts | 081 | Infection/inflammation of internal prosthetic device; implant; and graft |
| T83 | Complications of genitourinary prosthetic devices, implants, and grafts | 081 | Infection/inflammation of internal prosthetic device; implant; and graft |
| T84 | Complications of internal orthopedic prosthetic devices, implants, and grafts | 081 | Infection/inflammation of internal prosthetic device; implant; and graft |
| T85 | Complications of other internal prosthetic devices, implants, and grafts | 081 | Infection/inflammation of internal prosthetic device; implant; and graft |
| B37 | Candidiasis | 112 | Candidiasis |
| C45 | Mesothelioma | 165 | Cancer within the respiratory system |
| C34 | Malignant neoplasm of bronchus and lung | 165.1 | Cancer of bronchus; lung |
| D48 | Neoplasm of uncertain or unknown behavior of other and unspecified sites | 173 | Neoplasm of uncertain behavior of skin |
| C61 | Malignant neoplasm of prostate | 185 | Cancer of prostate |
| D07 | Carcinoma in situ of other and unspecified genital organs | 185 | Cancer of prostate |
| D09 | Carcinoma in situ of other and unspecified sites | 189 | Cancer of urinary organs (incl. kidney and bladder) |
| D09 | Carcinoma in situ of other and unspecified sites | 195 | Cancer, suspected or other |
| C78 | Secondary malignant neoplasm of respiratory and digestive organs | 198 | Secondary malignant neoplasm |
| C79 | Secondary malignant neoplasm of other sites | 198 | Secondary malignant neoplasm |
| D20 | Benign neoplasm of soft tissue of retroperitoneum and peritoneum | 199 | Neoplasm of uncertain behavior |
| D37 | Neoplasm of uncertain or unknown behavior of oral cavity and digestive organs | 199 | Neoplasm of uncertain behavior |
| D38 | Neoplasm of uncertain or unknown behavior of middle ear and respiratory and intrathoracic organs | 199 | Neoplasm of uncertain behavior |
| D39 | Neoplasm of uncertain or unknown behavior of female genital organs | 199 | Neoplasm of uncertain behavior |
| D41 | Neoplasm of uncertain or unknown behavior of urinary organs | 199 | Neoplasm of uncertain behavior |
| D44 | Neoplasm of uncertain or unknown behavior of endocrine glands | 199 | Neoplasm of uncertain behavior |
| D48 | Neoplasm of uncertain or unknown behavior of other and unspecified sites | 199 | Neoplasm of uncertain behavior |
| C96 | Other and unspecified malignant neoplasms of lymphoid, hematopoietic, and related tissue | 202 | Cancer of other lymphoid, histiocytic tissue |
| C82 | Follicular [nodular] non-Hodgkin's lymphoma | 202.2 | Non-Hodgkins lymphoma |
| C83 | Diffuse non-Hodgkin's lymphoma | 202.2 | Non-Hodgkins lymphoma |
| C85 | Other and unspecified types of non-Hodgkin's lymphoma | 202.2 | Non-Hodgkins lymphoma |
| D12 | Benign neoplasm of colon, rectum, anus, and anal canal | 208 | Benign neoplasm of colon |
| D22 | Melanocytic nevi | 216 | Benign neoplasm of skin |
| D23 | Other benign neoplasms of skin | 216 | Benign neoplasm of skin |
| E04 | Other nontoxic goiter | 241 | Nontoxic nodular goiter |
| E03 | Other hypothyroidism | 244.2 | Acquired hypothyroidism |
| E03 | Other hypothyroidism | 244.4 | Hypothyroidism NOS |
| E12 | Malnutrition-related diabetes mellitus | 249 | Secondary diabetes mellitus |
| E10 | Insulin-dependent diabetes mellitus | 250.1 | Type 1 diabetes |
| E11 | Non-insulin-dependent diabetes mellitus | 250.2 | Type 2 diabetes |
| E14 | Unspecified diabetes mellitus | 250.2 | Type 2 diabetes |
| R73 | Elevated blood glucose level | 250.41 | Impaired fasting glucose |
| N39 | Other disorders of urinary system | 269 | Proteinuria |
| M10 | Gout | 274.1 | Gout |
| E83 | Disorders of mineral metabolism | 275 | Disorders of mineral metabolism |
| E87 | Other disorders of fluid, electrolyte, and acid-base balance | 276.1 | Electrolyte imbalance |
| E87 | Other disorders of fluid, electrolyte, and acid-base balance | 276.4 | Acid-base balance disorder |
| E86 | Volume depletion | 276.5 | Hypovolemia |
| E87 | Other disorders of fluid, electrolyte, and acid-base balance | 276.6 | Fluid overload |
| E66 | Obesity | 278 | Overweight, obesity and other hyperalimentation |
| D89 | Other disorders involving the immune mechanism, not elsewhere classified | 279 | Disorders involving the immune mechanism |
| D50 | Iron deficiency anemia | 280.1 | Iron deficiency anemias, unspecified or not due to blood loss |
| D63 | Anaemia in chronic diseases classified elsewhere | 285.2 | Anemia of chronic disease |
| D69 | Purpura and other hemorrhagic conditions | 287 | Purpura and other hemorrhagic conditions |
| R59 | Enlarged lymph nodes | 289.4 | Lymphadenitis |
| R41 | Other symptoms and signs involving cognitive functions and awareness | 292 | Neurological disorders |
| G47 | Sleep disorders | 327 | Sleep disorders |
| G43 | Migraine | 340 | Migraine |
| G40 | Epilepsy | 345 | Epilepsy, recurrent seizures, convulsions |
| R26 | Abnormalities of gait and mobility | 350.2 | Abnormality of gait |
| H35 | Other retinal disorders | 362 | Other retinal disorders |
| H40 | Glaucoma | 365 | Glaucoma |
| H26 | Other cataract | 366 | Cataract |
| H25 | Senile cataract | 366.2 | Senile cataract |
| H52 | Disorders of refraction and accommodation | 367 | Disorders of refraction and accommodation; blindness and low vision |
| H53 | Visual disturbances | 368 | Visual disturbances |
| H02 | Other disorders of eyelid | 374 | Other disorders of eyelids |
| H57 | Other disorders of eye and adnexa | 379 | Other disorders of eye |
| H43 | Disorders of vitreous body | 379.2 | Disorders of vitreous body |
| H61 | Other disorders of external ear | 380.4 | Impacted cerumen |
| R42 | Dizziness and giddiness | 386.9 | Dizziness and giddiness (Light-headedness and vertigo) |
| H90 | Conductive and sensorineural hearing loss | 389 | Hearing loss |
| H91 | Other hearing loss | 389 | Hearing loss |
| I08 | Multiple valve diseases | 394 | Rheumatic disease of the heart valves |
| I34 | Nonrheumatic mitral valve disorders | 395.1 | Nonrheumatic mitral valve disorders |
| I35 | Nonrheumatic aortic valve disorders | 395.2 | Nonrheumatic aortic valve disorders |
| I10 | Essential (primary) hypertension | 401.1 | Essential hypertension |
| I12 | Hypertensive renal disease | 401.22 | Hypertensive chronic kidney disease |
| I21 | Acute myocardial infarction | 411.2 | Myocardial infarction |
| I22 | Subsequent myocardial infarction | 411.2 | Myocardial infarction |
| I24 | Other acute ischemic heart diseases | 411.2 | Myocardial infarction |
| I25 | Chronic ischemic heart disease | 411.2 | Myocardial infarction |
| I51 | Complications and ill-defined descriptions of heart disease | 411.2 | Myocardial infarction |
| I20 | Angina pectoris | 411.3 | Angina pectoris |
| I24 | Other acute ischemic heart diseases | 411.4 | Coronary atherosclerosis |
| I25 | Chronic ischemic heart disease | 411.4 | Coronary atherosclerosis |
| I46 | Cardiac arrest | 414 | Other forms of chronic heart disease |
| I51 | Complications and ill-defined descriptions of heart disease | 414 | Other forms of chronic heart disease |
| I26 | Pulmonary embolism | 415 | Pulmonary heart disease |
| I27 | Other pulmonary heart diseases | 415.2 | Chronic pulmonary heart disease |
| I51 | Complications and ill-defined descriptions of heart disease | 416 | Cardiomegaly |
| R07 | Pain in throat and chest | 418 | Nonspecific chest pain |
| I31 | Other diseases of pericardium | 420.2 | Pericarditis |
| B37 | Candidiasis | 420.3 | Endocarditis |
| I33 | Acute and subacute endocarditis | 420.3 | Endocarditis |
| I42 | Cardiomyopathy | 425 | Cardiomyopathy |
| I45 | Other conduction disorders | 426 | Cardiac conduction disorders |
| I44 | Atrioventricular and left bundle-branch block | 426 | Cardiac conduction disorders |
| I47 | Paroxysmal tachycardia | 427.1 | Paroxysmal tachycardia, unspecified |
| I48 | Atrial fibrillation and flutter | 427.2 | Atrial fibrillation and flutter |
| I49 | Other cardiac arrhythmias | 427.5 | Arrhythmia (cardiac) NOS |
| I49 | Other cardiac arrhythmias | 427.6 | Premature beats |
| I49 | Other cardiac arrhythmias | 427.8 | Sinoatrial node dysfunction (Bradycardia) |
| R00 | Abnormalities of heart beat | 427.9 | Palpitations |
| I50 | Heart failure | 428.1 | Congestive heart failure (CHF) NOS |
| I50 | Heart failure | 428.2 | Heart failure NOS |
| R94 | Abnormal results of function studies | 429.2 | Abnormal function study of cardiovascular system |
| R09 | Other symptoms and signs involving the circulatory and respiratory systems | 429.3 | Symptoms involving cardiovascular system |
| G45 | Transient cerebral ischemic attacks and related syndromes | 433 | Cerebrovascular disease |
| I60 | Subarachnoid hemorrhage | 433 | Cerebrovascular disease |
| I67 | Other cerebrovascular diseases | 433 | Cerebrovascular disease |
| I63 | Cerebral infarction | 433.2 | Occlusion of cerebral arteries |
| I64 | Stroke, not specified as hemorrhage or infarction | 433.21 | Cerebral artery occlusion, with cerebral infarction |
| I70 | Atherosclerosis | 440 | Atherosclerosis |
| I71 | Aortic aneurysm and dissection | 442.1 | Aortic aneurysm |
| I73 | Other peripheral vascular diseases | 443.9 | Peripheral vascular disease, unspecified |
| I74 | Arterial embolism and thrombosis | 444 | Arterial embolism and thrombosis |
| I82 | Other venous embolism and thrombosis | 452 | Other venous embolism and thrombosis |
| I86 | Varicose veins of other sites | 454 | Varicose veins |
| I84 | Hemorrhoids | 455 | Hemorrhoids |
| I87 | Other disorders of veins | 456 | Chronic venous insufficiency [CVI] |
| I95 | Hypotension | 458 | Hypotension |
| J01 | Acute sinusitis | 464 | Acute sinusitis |
| J02 | Acute pharyngitis | 465 | Acute upper respiratory infections of multiple or unspecified sites |
| J03 | Acute tonsillitis | 465 | Acute upper respiratory infections of multiple or unspecified sites |
| J06 | Acute upper respiratory infections of multiple and unspecified sites | 465 | Acute upper respiratory infections of multiple or unspecified sites |
| J31 | Chronic rhinitis, nasopharyngitis, and pharyngitis | 472 | Chronic pharyngitis and nasopharyngitis |
| J38 | Diseases of vocal cords and larynx, not elsewhere classified | 473 | Diseases of the larynx and vocal cords |
| R49 | Voice disturbances | 473.4 | Voice disturbance |
| J32 | Chronic sinusitis | 475 | Chronic sinusitis |
| R04 | Hemorrhage from respiratory passages | 477 | Epistaxis or throat hemorrhage |
| J18 | Pneumonia, organism unspecified | 480 | Pneumonia |
| J15 | Bacterial pneumonia, not elsewhere classified | 480.1 | Bacterial pneumonia |
| J05 | Acute obstructive laryngitis [croup] and epiglottitis | 483 | Acute bronchitis and bronchiolitis |
| J20 | Acute bronchitis | 483 | Acute bronchitis and bronchiolitis |
| J44 | Other chronic obstructive pulmonary disease | 496 | Chronic airway obstruction |
| J43 | Emphysema | 496.1 | Emphysema |
| J84 | Other interstitial pulmonary diseases | 502 | Postinflammatory pulmonary fibrosis |
| J84 | Other interstitial pulmonary diseases | 504 | Other alveolar and parietoalveolar pneumonopathy |
| J90 | Pleural effusion, not elsewhere classified | 507 | Pleurisy; pleural effusion |
| R09 | Other symptoms and signs involving the circulatory and respiratory systems | 507 | Pleurisy; pleural effusion |
| J98 | Other respiratory disorders | 508 | Pulmonary collapse; interstitial and compensatory emphysema |
| J96 | Respiratory failure, not elsewhere classified | 509.1 | Respiratory failure |
| R06 | Abnormalities of breathing | 513 | Respiratory abnormalities |
| R91 | Abnormal findings on diagnostic imaging of lung | 514 | Abnormal findings examination of lungs |
| J98 | Other respiratory disorders | 519 | Other diseases of respiratory system, not elsewhere classified |
| K22 | Other diseases of esophagus | 530 | Diseases of esophagus |
| K20 | Esophagitis | 530.1 | Esophagitis, GERD and related diseases |
| K21 | Gastroesophageal reflux disease | 530.11 | GERD |
| K26 | Duodenal ulcer | 531 | Peptic ulcer (excl. esophageal) |
| R13 | Dysphagia | 532 | Dysphagia |
| K31 | Other diseases of stomach and duodenum | 536 | Disorders of function of stomach |
| K43 | Ventral hernia | 550 | Abdominal hernia |
| K46 | Unspecified abdominal hernia | 550 | Abdominal hernia |
| K40 | Inguinal hernia | 550.1 | Inguinal hernia |
| K44 | Diaphragmatic hernia | 550.2 | Diaphragmatic hernia |
| K52 | Other noninfective gastroenteritis and colitis | 558 | Noninfectious gastroenteritis |
| R15 | Fecal incontinence | 561 | Symptoms involving digestive system |
| R19 | Other symptoms and signs involving the digestive system and abdomen | 561 | Symptoms involving digestive system |
| K57 | Diverticular disease of intestine | 562.1 | Diverticulosis |
| K59 | Other functional intestinal disorders | 564 | Functional digestive disorders |
| K58 | Irritable bowel syndrome | 564.1 | Irritable Bowel Syndrome |
| K74 | Fibrosis and cirrhosis of liver | 571.5 | Other chronic nonalcoholic liver disease |
| K76 | Other diseases of liver | 571.5 | Other chronic nonalcoholic liver disease |
| K70 | Alcoholic liver disease | 571.8 | Liver abscess and sequelae of chronic liver disease |
| K75 | Other inflammatory liver diseases | 571.8 | Liver abscess and sequelae of chronic liver disease |
| R18 | Ascites | 572 | Ascites (non malignant) |
| K76 | Other diseases of liver | 573 | Other disorders of liver |
| K80 | Cholelithiasis | 574.1 | Cholelithiasis |
| K86 | Other diseases of pancreas | 577 | Diseases of pancreas |
| K90 | Intestinal malabsorption | 577 | Diseases of pancreas |
| K62 | Other diseases of anus and rectum | 578.8 | Hemorrhage of rectum and anus |
| K92 | Other diseases of digestive system | 578.9 | Hemorrhage of gastrointestinal tract |
| N17 | Acute renal failure | 585.1 | Acute renal failure |
| N19 | Unspecified renal failure | 585.2 | Renal failure NOS |
| N18 | Chronic renal failure | 585.3 | Chronic renal failure [CKD] |
| M10 | Gout | 588 | Disorders resulting from impaired renal function |
| N39 | Other disorders of urinary system | 591 | Urinary tract infection |
| N02 | Recurrent and persistent hematuria | 593 | Hematuria |
| R31 | Unspecified hematuria | 593 | Hematuria |
| N20 | Calculus of kidney and ureter | 594 | Urinary calculus |
| R33 | Retention of urine | 599.2 | Retention of urine |
| R30 | Pain associated with micturition | 599.3 | Dysuria |
| N39 | Other disorders of urinary system | 599.4 | Urinary incontinence |
| R32 | Unspecified urinary incontinence | 599.4 | Urinary incontinence |
| R35 | Polyuria | 599.5 | Frequency of urination and polyuria |
| N40 | Hyperplasia of prostate | 600 | Hyperplasia of prostate |
| F52 | Sexual dysfunction, not caused by organic disorder or disease | 605 | Erectile dysfunction [ED] |
| N48 | Other disorders of penis | 605 | Erectile dysfunction [ED] |
| L03 | Cellulitis | 681 | Superficial cellulitis and abscess |
| L27 | Dermatitis due to substances taken internally | 687.1 | Rash and other nonspecific skin eruption |
| R21 | Rash and other nonspecific skin eruption | 687.1 | Rash and other nonspecific skin eruption |
| R20 | Disturbances of skin sensation | 687.4 | Disturbance of skin sensation |
| L90 | Atrophic disorders of the skin | 701.2 | Scar conditions and fibrosis of skin |
| L82 | Seborrheic keratosis | 702 | Degenerative skin conditions and other dermatoses |
| L97 | Ulcer of lower limb, not elsewhere classified | 707 | Chronic ulcer of skin |
| L89 | Decubitus ulcer | 707.1 | Decubitus ulcer |
| M06 | Other rheumatoid arthritis | 714 | Rheumatoid arthritis and other inflammatory polyarthropathies |
| M05 | Seropositive rheumatoid arthritis | 714.1 | Rheumatoid arthritis |
| M13 | Other arthritis | 716 | Other arthropathies |
| M48 | Other spondylopathies | 720 | Spinal stenosis |
| M47 | Spondylosis | 721 | Spondylosis and allied disorders |
| M48 | Other spondylopathies | 721 | Spondylosis and allied disorders |
| M50 | Cervical disc disorders | 722.1 | Displacement of intervertebral disc |
| M50 | Cervical disc disorders | 722.6 | Degeneration of intervertebral disc |
| M51 | Other intervertebral disc disorders | 722.6 | Degeneration of intervertebral disc |
| M75 | Shoulder lesions | 726 | Peripheral enthesopathies and allied syndromes |
| M77 | Other enthesopathies | 726 | Peripheral enthesopathies and allied syndromes |
| S46 | Injury of muscle and tendon at shoulder and upper arm level | 726 | Peripheral enthesopathies and allied syndromes |
| M25 | Other joint disorders, not elsewhere classified | 726.1 | Enthesopathy |
| M70 | Soft tissue disorders related to use, overuse, and pressure | 726.1 | Enthesopathy |
| M76 | Enthesopathies of lower limb, excluding foot | 726.1 | Enthesopathy |
| M16 | Coxarthrosis [arthrosis of hip] | 740.1 | Osteoarthritis; localized |
| M18 | Arthrosis of first carpometacarpal joint | 740.1 | Osteoarthritis; localized |
| M17 | Gonarthrosis [arthrosis of knee] | 740.11 | Osteoarthrosis, localized, primary |
| M15 | Polyarthrosis | 740.2 | Osteoarthrosis, generalized |
| M13 | Other arthritis | 740.9 | Osteoarthrosis NOS |
| M19 | Other arthrosis | 740.9 | Osteoarthrosis NOS |
| M22 | Disorders of patella | 741 | Symptoms and disorders of the joints |
| M25 | Other joint disorders, not elsewhere classified | 741 | Symptoms and disorders of the joints |
| M53 | Other dorsopathies, not elsewhere classified | 741 | Symptoms and disorders of the joints |
| R29 | Other symptoms and signs involving the nervous and musculoskeletal systems | 741 | Symptoms and disorders of the joints |
| M81 | Osteoporosis without pathological fracture | 743.11 | Osteoporosis NOS |
| M85 | Other disorders of bone density and structure | 743.9 | Osteopenia or other disorder of bone and cartilage |
| M89 | Other disorders of bone | 743.9 | Osteopenia or other disorder of bone and cartilage |
| Q21 | Congenital malformations of cardiac septa | 747.11 | Cardiac shunt/ heart septal defect |
