## Supplemental Table 3 for "Multi-ancestry gene-trait connection landscape using electronic health record (EHR) linked biobank data"

Supplementary Table 3. Statistically significant gene-disease associations in the eMERGE III individuals of European ancestry (P $\leq1.47\times{10}^{-10}$).

^a^ PhenomeXcan validation significance threshold P ≤ 5.49e-10

^b^ PMBB replication significance threshold P ≤ 6.39e-5

^c^ NA: Not applicable; no signal

^d^ NS: Not statistically significant

| Gene | Phecode Description | eMERGE | | PhenomeXcan UK Biobank | |
| --- | --- | --- | --- | --- | --- |
|  |  | Discovery P | Locus RCP | Replication P^a^ | Locus RCP |
| *HLA-DQA2* | Type 1 diabetes | 4.93695E-86 | NA^c^ | 3.3379E-117 | NA |
| *HLA-DRB1* | Rheumatoid arthritis and other inflammatory polyarthropathies | 3.76826E-50 | NA | 2.7532E-114 | 0.007073 |
| *HLA-DQA1* | Rheumatoid arthritis and other inflammatory polyarthropathies | 9.2901E-53 | NA | 4.3968E-110 | NA |
| *HLA-DQA1* | Type 1 diabetes | 8.86832E-42 | NA | 1.8557E-72 | 0.01045 |
| *HLA-DQB2* | Type 1 diabetes | 3.1885E-47 | NA | 1.5758E-69 | 0.191 |
| *HLA-DRA* | Rheumatoid arthritis and other inflammatory polyarthropathies | 2.16757E-27 | NA | 3.5307E-69 | NA |
| *HLA-DRB1* | Type 1 diabetes | 1.36331E-48 | NA | 2.2732E-63 | NA |
| *LPA* | Coronary atherosclerosis | 2.02176E-26 | NA | 1.3923E-62 | NA |
| *LPA* | Myocardial infarction | 4.29515E-18 | NA | 1.3923E-62 | NA |
| *BTNL2* | Rheumatoid arthritis and other inflammatory polyarthropathies | 1.35055E-34 | NA | 1.1812E-58 | NA |
| *HLA-DQB1* | Type 1 diabetes | 3.80849E-46 | NA | 1.2068E-55 | 0.07306 |
| *HLA-DRB1* | Rheumatoid arthritis | 4.75929E-59 | NA | 3.2511E-55 | 0.0002586 |
| *EHMT2* | Type 1 diabetes | 6.04941E-24 | NA | 8.4645E-53 | NA |
| *HLA-DQA1* | Rheumatoid arthritis | 1.98474E-59 | NA | 4.4905E-51 | 0.000575 |
| *PBX2* | Rheumatoid arthritis and other inflammatory polyarthropathies | 1.52324E-17 | NA | 8.1596E-51 | NA |
| *C6orf10* | Rheumatoid arthritis and other inflammatory polyarthropathies | 1.4813E-14 | NA | 2.8972E-46 | NA |
| *PBX2* | Type 1 diabetes | 4.31115E-16 | NA | 3.3048E-45 | NA |
| *PSMB9* | Rheumatoid arthritis and other inflammatory polyarthropathies | 2.24049E-14 | NA | 2.9265E-44 | NA |
| *CDKN2B* | Myocardial infarction | 5.91935E-16 | NA | 3.5621E-43 | 0.1864 |
| *CDKN2B* | Coronary atherosclerosis | 3.3355E-21 | NA | 3.5621E-43 | 0.1864 |
| *C4B* | Type 1 diabetes | 2.58508E-25 | 0.3218 | 1.5254E-39 | NA |
| *AIF1* | Type 1 diabetes | 1.0808E-15 | NA | 5.0306E-39 | NA |
| *HLA-DRA* | Type 1 diabetes | 3.1815E-32 | NA | 1.2836E-37 | NA |
| *HLA-DQB1* | Rheumatoid arthritis and other inflammatory polyarthropathies | 1.28275E-15 | NA | 1.1141E-36 | NA |
| *CSNK2B* | Type 1 diabetes | 6.78096E-17 | NA | 1.2363E-36 | NA |
| *CYP21A2* | Type 1 diabetes | 6.20286E-37 | NA | 1.6187E-36 | NA |
| *ABHD16A* | Type 1 diabetes | 5.32833E-19 | NA | 1.7656E-36 | NA |
| *BTNL2* | Type 1 diabetes | 1.68991E-34 | NA | 1.2343E-34 | NA |
| *MSH5* | Type 1 diabetes | 1.16058E-18 | NA | 1.7127E-34 | NA |
| *C2* | Type 1 diabetes | 2.03224E-19 | NA | 5.3222E-34 | NA |
| *HLA-DQB2* | Rheumatoid arthritis and other inflammatory polyarthropathies | 9.47669E-14 | NA | 2.7037E-33 | 0.005891 |
| *HLA-DQA2* | Rheumatoid arthritis and other inflammatory polyarthropathies | 2.84878E-25 | NA | 4.4199E-33 | NA |
| *TNXB* | Type 1 diabetes | 1.65536E-19 | NA | 4.8232E-32 | NA |
| *CSNK2B* | Rheumatoid arthritis and other inflammatory polyarthropathies | 3.08399E-15 | NA | 5.6164E-32 | 0.09446 |
| *HLA-DRA* | Rheumatoid arthritis | 6.5742E-38 | NA | 1.3649E-31 | NA |
| *GPANK1* | Type 1 diabetes | 3.59207E-16 | NA | 5.5248E-31 | NA |
| *C4B* | Rheumatoid arthritis and other inflammatory polyarthropathies | 2.4356E-18 | NA | 8.5051E-31 | 0.008272 |
| *DDAH2* | Type 1 diabetes | 1.37164E-18 | NA | 1.0361E-30 | NA |
| *PRRC2A* | Type 1 diabetes | 1.40123E-15 | NA | 3.1882E-29 | NA |
| *BTNL2* | Rheumatoid arthritis | 2.53591E-42 | NA | 3.563E-29 | NA |
| *ABO* | Pulmonary heart disease | 1.31221E-13 | NA | 1.5261E-27 | 0.559 |
| *FKBPL* | Rheumatoid arthritis and other inflammatory polyarthropathies | 2.73742E-13 | NA | 1.5884E-27 | NA |
| *MPIG6B* | Type 1 diabetes | 3.47404E-12 | NA | 1.6256E-27 | NA |
| *CFB* | Type 1 diabetes | 6.74546E-11 | NA | 7.7428E-27 | NA |
| *VWA7* | Type 1 diabetes | 1.27608E-14 | NA | 6.7779E-26 | NA |
| *BAG6* | Rheumatoid arthritis and other inflammatory polyarthropathies | 5.50817E-14 | NA | 8.6436E-26 | NA |
| *CLIC1* | Type 1 diabetes | 7.61349E-17 | NA | 1.9317E-25 | NA |
| *ZBTB12* | Type 1 diabetes | 6.06503E-15 | NA | 3.7373E-25 | NA |
| *BAG6* | Type 1 diabetes | 5.82821E-15 | NA | 2.3621E-24 | NA |
| *HLA-DRB5* | Type 1 diabetes | 3.93801E-49 | NA | 1.1692E-23 | NA |
| *POU5F1B* | Cancer of prostate | 5.72323E-12 | NA | 5.3814E-23 | 0.003293 |
| *CYP21A2* | Rheumatoid arthritis and other inflammatory polyarthropathies | 8.74655E-11 | NA | 7.0562E-23 | NA |
| *TAP2* | Type 1 diabetes | 5.68647E-16 | NA | 1.2162E-22 | NA |
| *LY6G6F* | Type 1 diabetes | 6.58481E-13 | NA | 2.1997E-22 | NA |
| *LY6G6C* | Type 1 diabetes | 4.89987E-14 | NA | 2.9938E-22 | NA |
| *C6orf10* | Rheumatoid arthritis | 8.84137E-17 | NA | 6.2762E-22 | NA |
| *ABHD16A* | Rheumatoid arthritis and other inflammatory polyarthropathies | 1.6668E-11 | NA | 1.4658E-21 | NA |
| *PRRC2A* | Rheumatoid arthritis and other inflammatory polyarthropathies | 4.48952E-13 | NA | 1.9497E-21 | NA |
| *PSMB9* | Rheumatoid arthritis | 5.86845E-19 | NA | 2.589E-21 | NA |
| *BRD2* | Rheumatoid arthritis and other inflammatory polyarthropathies | 1.49781E-18 | NA | 4.6294E-21 | NA |
| *PRRT1* | Type 1 diabetes | 2.34788E-11 | NA | 4.6556E-21 | NA |
| *PBX2* | Rheumatoid arthritis | 1.34916E-19 | NA | 1.2887E-19 | NA |
| *C6orf48* | Type 1 diabetes | 6.01858E-16 | NA | 1.9121E-19 | NA |
| *DDR1* | Type 1 diabetes | 4.78629E-11 | NA | 2.7733E-19 | NA |
| *APOM* | Type 1 diabetes | 4.99037E-17 | NA | 5.1645E-19 | NA |
| *AIF1* | Rheumatoid arthritis and other inflammatory polyarthropathies | 1.00708E-14 | NA | 1.9567E-18 | NA |
| *LPA* | Myocardial infarction | 4.29515E-18 | NA | 7.256E-18 | NA |
| *GPANK1* | Rheumatoid arthritis and other inflammatory polyarthropathies | 3.09304E-14 | NA | 3.2026E-17 | NA |
| *C4A* | Type 1 diabetes | 1.46885E-19 | 0.6692 | 4.4814E-17 | NA |
| *AGER* | Type 1 diabetes | 4.66785E-13 | NA | 6.2349E-17 | NA |
| *HLA-DQB2* | Rheumatoid arthritis | 4.59737E-16 | NA | 3.8886E-16 | 0.005961 |
| *HLA-DQA2* | Rheumatoid arthritis | 1.24827E-31 | NA | 5.2876E-16 | 0.002289 |
| *HLA-DQB1* | Rheumatoid arthritis | 2.721E-15 | NA | 8.4273E-16 | NA |
| *LPA* | Angina pectoris | 3.89643E-13 | NA | 1.0842E-15 | 0.004976 |
| *SAPCD1* | Type 1 diabetes | 3.04132E-14 | NA | 1.5149E-15 | NA |
| *DDX39B* | Rheumatoid arthritis and other inflammatory polyarthropathies | 9.43461E-20 | 0.7973 | 5.988E-15 | 0.005139 |
| *PHACTR1* | Coronary atherosclerosis | 1.82278E-11 | 0.9998 | 2.0381E-14 | 1.001 |
| *HLA-DQA2* | Type 2 diabetes | 6.20882E-19 | NA | 3.7509E-14 | NA |
| *AGER* | Rheumatoid arthritis and other inflammatory polyarthropathies | 1.18304E-15 | NA | 7.2246E-14 | NA |
| *NOTCH4* | Type 1 diabetes | 1.07167E-16 | NA | 8.163E-14 | NA |
| *C4B* | Rheumatoid arthritis | 2.76312E-23 | NA | 9.8963E-14 | 0.0003578 |
| *BRD2* | Rheumatoid arthritis | 1.80983E-20 | NA | 5.6827E-13 | NA |
| *FKBPL* | Rheumatoid arthritis | 1.25075E-11 | NA | 8.7031E-13 | NA |
| *VARS* | Type 1 diabetes | 1.74479E-14 | NA | 1.0659E-12 | NA |
| *CDKN2B* | Myocardial infarction | 5.91935E-16 | NA | 1.9644E-12 | 0.1959 |
| *SAPCD1* | Rheumatoid arthritis and other inflammatory polyarthropathies | 8.34151E-12 | NA | 3.7978E-12 | NA |
| *BAG6* | Rheumatoid arthritis | 1.262E-13 | NA | 1.7436E-11 | NA |
| *LTA* | Rheumatoid arthritis and other inflammatory polyarthropathies | 9.81431E-18 | 0.2274 | 1.9487E-11 | 0.04113 |
| *HLA-DMB* | Type 1 diabetes | 8.39696E-11 | NA | 2.1903E-11 | NA |
| *HLA-DQB1* | Type 2 diabetes | 2.37542E-11 | NA | 3.7094E-11 | 0.06962 |
| *FAM13B* | Atrial fibrillation and flutter | 2.59001E-12 | NA | 5.8451E-11 | 0.91 |
| *LST1* | Rheumatoid arthritis and other inflammatory polyarthropathies | 3.87946E-12 | NA | 1.7475E-10 | 0.04971 |
| *NFKBIL1* | Rheumatoid arthritis and other inflammatory polyarthropathies | 2.62927E-20 | 0.2262 | 2.0709E-10 | 0.01361 |
| *CSNK2B* | Rheumatoid arthritis | 2.84052E-13 | NA | 2.4443E-10 | NA |
| *HLA-DQB2* | Type 2 diabetes | 1.65211E-11 | NA | 4.8335E-10 | 0.2059 |
| *LPA* | Ischemic Heart Disease | 8.89759E-24 | NA | NA | NA |
| *IRF4* | Dyschromia and Vitiligo | 1.52613E-11 | 0.2702 | NA | NA |
| *DBNDD1* | Degenerative skin conditions and other dermatoses | 6.8123E-11 | NA | NA | NA |
| *IRF4* | Degenerative skin conditions and other dermatoses | 1.70529E-20 | 0.289 | NA | NA |
| *SPIRE2* | Degenerative skin conditions and other dermatoses | 5.71824E-11 | NA | NA | NA |
| *IRF4* | Seborrheic keratosis | 5.32526E-14 | 0.2823 | NA | NA |
| *TERT* | Seborrheic keratosis | 1.45308E-18 | NA | NA | NA |
| *NOTCH4* | Rheumatoid arthritis and other inflammatory polyarthropathies | 3.0083E-12 | NA | NA | NA |
| *XXbac-BPG181B23.7* | Rheumatoid arthritis and other inflammatory polyarthropathies | 1.63539E-16 | NA | NA | NA |
| *LINC01149* | Rheumatoid arthritis and other inflammatory polyarthropathies | 6.22497E-20 | NA | NA | NA |
| *DDX39B* | Rheumatoid arthritis | 4.66274E-17 | NA | NA | NA |
| *LTA* | Rheumatoid arthritis | 5.85388E-14 | NA | NA | NA |
| *AGER* | Rheumatoid arthritis | 2.80194E-19 | NA | NA | NA |
| *GPANK1* | Rheumatoid arthritis | 2.11406E-13 | NA | NA | NA |
| *HSPA1B* | Rheumatoid arthritis | 9.20928E-11 | NA | NA | NA |
| *XXbac-BPG181B23.7* | Rheumatoid arthritis | 1.34098E-13 | NA | NA | NA |
| *NFKBIL1* | Rheumatoid arthritis | 1.85852E-16 | NA | NA | NA |
| *HLA-DRB5* | Rheumatoid arthritis | 4.44866E-13 | NA | NA | NA |
| *ABHD16A* | Rheumatoid arthritis | 3.32048E-13 | NA | NA | NA |
| *AIF1* | Rheumatoid arthritis | 1.27025E-12 | NA | NA | NA |
| *LINC01149* | Rheumatoid arthritis | 7.71006E-18 | NA | NA | NA |
| *PRRC2A* | Rheumatoid arthritis | 1.53098E-13 | NA | NA | NA |
| *DBNDD1* | Neoplasm of uncertain behavior of skin | 5.11563E-11 | NA | NA | NA |
| *IRF4* | Neoplasm of uncertain behavior of skin | 1.11891E-11 | 0.2024 | NA | NA |
| *SPIRE2* | Neoplasm of uncertain behavior of skin | 2.9904E-12 | NA | NA | NA |
| *HCG22* | Hypothyroidism | 6.97963E-11 | NA | NA | NA |
| *CYP21A2* | Hypothyroidism | 9.89224E-11 | NA | NA | NA |
| *HLA-DQB2* | Hypothyroidism | 4.27167E-22 | NA | NA | NA |
| *HLA-DQA2* | Hypothyroidism | 4.27167E-22 | 0.9323 | NA | NA |
| *TRMO* | Hypothyroidism | 2.36235E-19 | NA | NA | NA |
| *CTLA4* | Hypothyroidism | 2.50859E-14 | NA | NA | NA |
| *HLA-DQB1* | Hypothyroidism | 1.24837E-17 | NA | NA | NA |
| *HLA-DQA1* | Hypothyroidism | 3.57834E-19 | NA | NA | NA |
| *TCF19* | Hypothyroidism | 1.17335E-13 | 0.4857 | NA | NA |
| *PSORS1C1* | Hypothyroidism | 1.36899E-13 | NA | NA | NA |
| *FOXE1* | Hypothyroidism | 8.8031E-21 | NA | NA | NA |
| *HLA-DRB1* | Hypothyroidism | 7.60992E-19 | NA | NA | NA |
| *HLA-DOA* | Hypothyroidism | 1.69429E-11 | NA | NA | NA |
| *HLA-B* | Hypothyroidism | 4.50827E-12 | NA | NA | NA |
| *C4B* | Hypothyroidism | 2.40775E-11 | NA | NA | NA |
| *PSMB9* | Hypothyroidism | 6.42542E-11 | NA | NA | NA |
| *HLA-DRB5* | Hypothyroidism | 5.03682E-12 | NA | NA | NA |
| *HLA-DQA1* | Hypothyroidism NOS | 7.03515E-19 | NA | NA | NA |
| *CYP21A2* | Hypothyroidism NOS | 1.13662E-10 | NA | NA | NA |
| *HLA-DQB2* | Hypothyroidism NOS | 1.04748E-21 | NA | NA | NA |
| *HLA-DQA2* | Hypothyroidism NOS | 1.04748E-21 | 0.9447 | NA | NA |
| *TRMO* | Hypothyroidism NOS | 1.46954E-22 | NA | NA | NA |
| *HLA-DQB1* | Hypothyroidism NOS | 1.5131E-17 | NA | NA | NA |
| *TCF19* | Hypothyroidism NOS | 3.22167E-14 | 0.4348 | NA | NA |
| *PSORS1C1* | Hypothyroidism NOS | 3.89414E-14 | NA | NA | NA |
| *HLA-DOA* | Hypothyroidism NOS | 3.2961E-12 | NA | NA | NA |
| *HLA-DRB1* | Hypothyroidism NOS | 3.15066E-19 | NA | NA | NA |
| *FOXE1* | Hypothyroidism NOS | 1.0784E-21 | NA | NA | NA |
| *HLA-DRB5* | Hypothyroidism NOS | 2.20893E-12 | NA | NA | NA |
| *PSMB9* | Hypothyroidism NOS | 1.12641E-10 | NA | NA | NA |
| *CTLA4* | Hypothyroidism NOS | 9.60218E-14 | NA | NA | NA |
| *HLA-B* | Hypothyroidism NOS | 8.78215E-12 | NA | NA | NA |
| *WNT2* | Secondary diabetes mellitus | 9.08741E-22 | NA | NA | NA |
| *CFTR* | Secondary diabetes mellitus | 7.91921E-15 | NA | NA | NA |
| *HLA-DQB1* | Diabetes mellitus | 1.22008E-15 | NA | NA | NA |
| *HLA-DRB1* | Diabetes mellitus | 2.71065E-12 | NA | NA | NA |
| *HLA-DQA1* | Diabetes mellitus | 4.07896E-13 | NA | NA | NA |
| *CYP21A2* | Diabetes mellitus | 1.30484E-13 | NA | NA | NA |
| *HLA-DQB2* | Diabetes mellitus | 1.88269E-14 | NA | NA | NA |
| *HLA-DQA2* | Diabetes mellitus | 5.73244E-25 | NA | NA | NA |
| *TCF7L2* | Diabetes mellitus | 6.25941E-30 | NA | NA | NA |
| *BTNL2* | Diabetes mellitus | 3.32654E-11 | NA | NA | NA |
| *HLA-DOB* | Type 1 diabetes | 1.27547E-13 | NA | NA | NA |
| *TCF7L2* | Type 1 diabetes | 8.41488E-12 | NA | NA | NA |
| *NEU1* | Type 1 diabetes | 2.42375E-14 | NA | NA | NA |
| *CCND2* | Type 2 diabetes | 1.18899E-10 | NA | NA | NA |
| *CYP21A2* | Type 2 diabetes | 3.09604E-11 | NA | NA | NA |
| *TCF7L2* | Type 2 diabetes | 7.9294E-30 | NA | NA | NA |
| *TCF7L2* | Type 2 diabetes with neurological manifestations | 1.84232E-13 | NA | NA | NA |
| *HLA-DQA2* | Polyneuropathy in diabetes | 9.84904E-16 | NA | NA | NA |
| *WNT2* | severe protein-calorie malnutrition | 9.66283E-11 | NA | NA | NA |
| *PSRC1* | Disorders of lipoid metabolism | 8.36892E-19 | 0.8301 | NA | NA |
| *APOB* | Disorders of lipoid metabolism | 3.98174E-11 | NA | NA | NA |
| *NECTIN2* | Disorders of lipoid metabolism | 1.48759E-11 | NA | NA | NA |
| *APOE* | Disorders of lipoid metabolism | 4.4254E-23 | NA | NA | NA |
| *CELSR2* | Disorders of lipoid metabolism | 7.24943E-14 | NA | NA | NA |
| *TOMM40* | Disorders of lipoid metabolism | 2.00358E-20 | NA | NA | NA |
| *PSMA5* | Disorders of lipoid metabolism | 1.13415E-16 | 0.1118 | NA | NA |
| *LDLR* | Disorders of lipoid metabolism | 1.14156E-15 | 0.6376 | NA | NA |
| *LPA* | Disorders of lipoid metabolism | 3.11952E-14 | NA | NA | NA |
| *PSRC1* | Hyperlipidemia | 4.66068E-19 | 0.8321 | NA | NA |
| *APOB* | Hyperlipidemia | 3.45946E-11 | NA | NA | NA |
| *NECTIN2* | Hyperlipidemia | 1.14513E-11 | NA | NA | NA |
| *APOE* | Hyperlipidemia | 3.29495E-23 | NA | NA | NA |
| *CELSR2* | Hyperlipidemia | 2.89576E-14 | NA | NA | NA |
| *TOMM40* | Hyperlipidemia | 1.85826E-20 | NA | NA | NA |
| *PSMA5* | Hyperlipidemia | 6.5125E-17 | 0.1119 | NA | NA |
| *LDLR* | Hyperlipidemia | 8.43405E-16 | 0.6377 | NA | NA |
| *LPA* | Hyperlipidemia | 3.2591E-14 | NA | NA | NA |
| *APOB* | Hypercholesterolemia | 1.53414E-12 | NA | NA | NA |
| *NECTIN2* | Hypercholesterolemia | 9.51113E-14 | NA | NA | NA |
| *APOE* | Hypercholesterolemia | 1.42873E-19 | NA | NA | NA |
| *PSRC1* | Hypercholesterolemia | 4.16191E-16 | 0.875 | NA | NA |
| *CELSR2* | Hypercholesterolemia | 1.16702E-11 | 0.1088 | NA | NA |
| *TOMM40* | Hypercholesterolemia | 7.64607E-19 | NA | NA | NA |
| *PSMA5* | Hypercholesterolemia | 8.28503E-14 | 0.113 | NA | NA |
| *LDLR* | Hypercholesterolemia | 1.50672E-16 | 0.6401 | NA | NA |
| *LPA* | Hypercholesterolemia | 2.74401E-13 | NA | NA | NA |
| *PSRC1* | Mixed hyperlipidemia | 1.42694E-12 | 0.8451 | NA | NA |
| *PSMA5* | Mixed hyperlipidemia | 4.35257E-11 | NA | NA | NA |
| *WDR1* | Gout and other crystal arthropathies | 2.89077E-23 | NA | NA | NA |
| *SLC2A9* | Gout and other crystal arthropathies | 8.32413E-25 | NA | NA | NA |
| *PKD2* | Gout and other crystal arthropathies | 1.21648E-23 | NA | NA | NA |
| *ABCG2* | Gout and other crystal arthropathies | 4.46819E-12 | NA | NA | NA |
| *SPP1* | Gout and other crystal arthropathies | 4.62267E-13 | NA | NA | NA |
| *WDR1* | Gout | 1.74346E-23 | NA | NA | NA |
| *SLC2A9* | Gout | 1.96481E-25 | NA | NA | NA |
| *PKD2* | Gout | 3.54066E-26 | NA | NA | NA |
| *BICC1* | Gout | 3.06862E-11 | NA | NA | NA |
| *ABCG2* | Gout | 1.62472E-12 | NA | NA | NA |
| *SPP1* | Gout | 1.18579E-13 | NA | NA | NA |
| *WDR1* | Gouty arthropathy | 5.90234E-12 | NA | NA | NA |
| *SLC2A9* | Gouty arthropathy | 9.12332E-11 | 0.1472 | NA | NA |
| *PKD2* | Gouty arthropathy | 1.53869E-15 | NA | NA | NA |
| *ABCG2* | Gouty arthropathy | 1.60587E-11 | NA | NA | NA |
| *FTO* | Overweight, obesity and other hyperalimentation | 2.23357E-13 | NA | NA | NA |
| *FTO* | Obesity | 7.2905E-15 | NA | NA | NA |
| *FTO* | Morbid obesity | 8.06527E-12 | NA | NA | NA |
| *SELE* | Coagulation defects | 3.96987E-14 | NA | NA | NA |
| *RP1-206D15.6* | Coagulation defects | 1.27437E-23 | NA | NA | NA |
| *SLC19A2* | Coagulation defects | 4.92777E-51 | NA | NA | NA |
| *ATP1B1* | Coagulation defects | 3.28165E-24 | NA | NA | NA |
| *CCDC181* | Coagulation defects | 7.33266E-41 | NA | NA | NA |
| *PSMD3* | Elevated white blood cell count | 2.61723E-11 | 0.5802 | NA | NA |
| *APOC1* | Neurological disorders | 1.38217E-11 | NA | NA | NA |
| *TOMM40* | Neurological disorders | 1.64785E-16 | NA | NA | NA |
| *APOC1* | Memory loss | 2.42815E-18 | NA | NA | NA |
| *TOMM40* | Memory loss | 6.72921E-25 | NA | NA | NA |
| *APOE* | Memory loss | 4.15569E-20 | NA | NA | NA |
| *HTRA1* | Other retinal disorders | 4.00144E-22 | NA | NA | NA |
| *CFHR1* | Other retinal disorders | 5.46181E-15 | NA | NA | NA |
| *ARMS2* | Other retinal disorders | 2.17914E-16 | NA | NA | NA |
| *CFH* | Other retinal disorders | 6.66067E-12 | NA | NA | NA |
| *CFHR3* | Degeneration of macula and posterior pole of retina | 8.78802E-14 | NA | NA | NA |
| *HTRA1* | Degeneration of macula and posterior pole of retina | 8.1474E-32 | NA | NA | NA |
| *CFHR1* | Degeneration of macula and posterior pole of retina | 4.80436E-24 | NA | NA | NA |
| *CFH* | Degeneration of macula and posterior pole of retina | 1.64667E-19 | NA | NA | NA |
| *ARMS2* | Degeneration of macula and posterior pole of retina | 1.03954E-24 | NA | NA | NA |
| *LPA* | Heart valve disorders | 5.10915E-11 | NA | NA | NA |
| *LPA* | Nonrheumatic aortic valve disorders | 1.34994E-12 | NA | NA | NA |
| *CDKN2B* | Ischemic Heart Disease | 5.72061E-18 | NA | NA | NA |
| *LPA* | Unstable angina (intermediate coronary syndrome) | 8.1724E-13 | NA | NA | NA |
| *CDKN2B* | Other chronic ischemic heart disease, unspecified | 2.29607E-17 | NA | NA | NA |
| *LPA* | Other chronic ischemic heart disease, unspecified | 3.52299E-20 | NA | NA | NA |
| *ABO* | Acute pulmonary heart disease | 2.433E-16 | 0.1611 | NA | NA |
| *F11* | Acute pulmonary heart disease | 1.2813E-10 | NA | NA | NA |
| *ABO* | Pulmonary embolism and infarction, acute | 1.08195E-16 | 0.162 | NA | NA |
| *FAM13B* | Atrial fibrillation | 4.93093E-12 | NA | NA | NA |
| *LPA* | Occlusion and stenosis of precerebral arteries | 7.39247E-17 | NA | NA | NA |
| *IER3* | Atherosclerosis | 5.03072E-13 | NA | NA | NA |
| *LINC00243* | Atherosclerosis | 5.47461E-12 | NA | NA | NA |
| *FLOT1* | Atherosclerosis | 2.81886E-11 | NA | NA | NA |
| *BTNL2* | Atherosclerosis | 3.23144E-13 | NA | NA | NA |
| *DDR1* | Atherosclerosis | 8.47131E-11 | NA | NA | NA |
| *LPA* | Atherosclerosis | 8.79681E-12 | NA | NA | NA |
| *FLOT1* | Atherosclerosis of the extremities | 4.90906E-15 | NA | NA | NA |
| *IER3* | Atherosclerosis of the extremities | 1.32788E-20 | NA | NA | NA |
| *PPP1R18* | Atherosclerosis of the extremities | 2.54182E-11 | NA | NA | NA |
| *HLA-DQA1* | Atherosclerosis of the extremities | 9.97743E-11 | NA | NA | NA |
| *LINC00243* | Atherosclerosis of the extremities | 1.11463E-18 | NA | NA | NA |
| *CCHCR1* | Atherosclerosis of the extremities | 1.19315E-11 | NA | NA | NA |
| *BTNL2* | Atherosclerosis of the extremities | 3.35744E-17 | NA | NA | NA |
| *DDR1* | Atherosclerosis of the extremities | 3.03206E-17 | NA | NA | NA |
| *HCG20* | Atherosclerosis of the extremities | 1.27587E-13 | NA | NA | NA |
| *TCF19* | Atherosclerosis of the extremities | 7.29805E-11 | NA | NA | NA |
| *TRIM31* | Atherosclerosis of the extremities | 1.45048E-10 | NA | NA | NA |
| *XXbac-BPG13B8.10* | Atherosclerosis of the extremities | 1.44321E-13 | NA | NA | NA |
| *OR2H2* | Atherosclerosis of the extremities | 7.42672E-11 | NA | NA | NA |
| *LPA* | Atherosclerosis of the extremities | 3.56759E-14 | NA | NA | NA |
| *IER3* | Atherosclerosis of native arteries of the extremities with intermittent claudication | 6.51503E-14 | NA | NA | NA |
| *LINC00243* | Atherosclerosis of native arteries of the extremities with intermittent claudication | 2.47691E-17 | NA | NA | NA |
| *DDR1* | Atherosclerosis of native arteries of the extremities with intermittent claudication | 1.06076E-15 | NA | NA | NA |
| *LPA* | Atherosclerosis of native arteries of the extremities with intermittent claudication | 5.56361E-16 | NA | NA | NA |
| *CDKN2B* | Peripheral vascular disease | 3.24239E-12 | NA | NA | NA |
| *DDR1* | Peripheral vascular disease | 7.61794E-12 | NA | NA | NA |
| *CDKN2B* | Peripheral vascular disease, unspecified | 4.97931E-13 | NA | NA | NA |
| *IER3* | Peripheral vascular disease, unspecified | 1.11887E-10 | NA | NA | NA |
| *LPA* | Peripheral vascular disease, unspecified | 2.62006E-13 | NA | NA | NA |
| *ABO* | Other venous embolism and thrombosis | 3.42749E-26 | 0.1594 | NA | NA |
| *RP1-206D15.6* | Other venous embolism and thrombosis | 1.23212E-12 | NA | NA | NA |
| *SLC19A2* | Other venous embolism and thrombosis | 2.92833E-29 | NA | NA | NA |
| *RPL7A* | Other venous embolism and thrombosis | 2.59937E-12 | NA | NA | NA |
| *MED22* | Other venous embolism and thrombosis | 1.44158E-12 | NA | NA | NA |
| *LRAT* | Other venous embolism and thrombosis | 1.49508E-12 | NA | NA | NA |
| *CCDC181* | Other venous embolism and thrombosis | 5.83984E-18 | NA | NA | NA |
| *ABO* | Deep vein thrombosis [DVT] | 9.1287E-16 | 0.1293 | NA | NA |
| *SLC19A2* | Deep vein thrombosis [DVT] | 5.51272E-24 | NA | NA | NA |
| *LRAT* | Deep vein thrombosis [DVT] | 4.01857E-11 | NA | NA | NA |
| *CCDC181* | Deep vein thrombosis [DVT] | 1.67961E-12 | NA | NA | NA |
| *WNT2* | Bacterial pneumonia | 5.49342E-18 | NA | NA | NA |
| *CFTR* | Bacterial pneumonia | 1.68577E-11 | NA | NA | NA |
| *PNPLA3* | Chronic liver disease and cirrhosis | 8.0175E-29 | NA | NA | NA |
| *PNPLA3* | Other chronic nonalcoholic liver disease | 3.59333E-32 | NA | NA | NA |
| *PNPLA3* | Cirrhosis of liver without mention of alcohol | 6.29524E-19 | NA | NA | NA |
| *WNT2* | Diseases of pancreas | 2.72639E-28 | NA | NA | NA |
| *CFTR* | Diseases of pancreas | 5.5527E-17 | NA | NA | NA |
| *PRAME* | Renal failure | 1.08685E-10 | NA | NA | NA |
| *CYP21A2* | Renal failure | 2.15631E-11 | 0.1213 | NA | NA |
| *C4A* | Renal failure | 1.08212E-11 | 0.7481 | NA | NA |
| *C4B* | Renal failure | 9.37572E-11 | 0.2854 | NA | NA |
| *TNXB* | Renal failure | 2.46659E-11 | NA | NA | NA |
| *C4A* | Chronic renal failure [CKD] | 5.95181E-11 | 0.6167 | NA | NA |
| *CYP21A2* | End stage renal disease | 1.21016E-11 | NA | NA | NA |
| *C4A* | End stage renal disease | 3.84406E-11 | 0.1875 | NA | NA |
| *PTCSC2* | Hypothyroidism | 4.58988E-14 | NA | NA | NA |
| *C4B* | Hypothyroidism NOS | 5.79024E-11 | NA | NA | NA |
| *PTCSC2* | Hypothyroidism NOS | 1.60649E-15 | NA | NA | NA |
| *HLA-DRB5* | Diabetes mellitus | 8.20116E-16 | NA | NA | NA |
| *HLA-DRA* | Diabetes mellitus | 1.37161E-10 | NA | NA | NA |
| *HLA-DRB5* | Type 2 diabetes | 9.7195E-13 | NA | NA | NA |
| *CYP2R1* | Vitamin deficiency | 5.1894E-15 | NA | NA | NA |
| *CYP2R1* | Vitamin D deficiency | 3.21904E-19 | NA | NA | NA |
| *APOE* | Neurological disorders | 1.46113E-11 | NA | NA | NA |
| *POU5F1* | Atherosclerosis of the extremities | 1.17319E-10 | NA | NA | NA |
| *DDR1* | Arterial embolism and thrombosis | 3.30567E-12 | NA | NA | NA |
| *SAPCD1* | Rheumatoid arthritis | 2.68511E-11 | NA | NA | NA |
