## Supplemental Table 4 for "Multi-ancestry gene-trait connection landscape using electronic health record (EHR) linked biobank data"

Supplementary Table 4. Statistically significant gene-disease associations in the eMERGE III individuals of African ancestry (P $\leq1.47\times{10}^{-10}$).

^a^ PMBB replication significance threshold P ≤ 6.39e-5

^b^ NA: not applicable

^c^ NS: not statistically significant

| Gene | Phecode Description | eMERGE | |
| --- | --- | --- | --- |
|  |  | Discovery P | Locus RCP |
| *GOLGA6B* | Acute pulmonary heart disease | 3.1062E-11 | NA^b^ |
| *CTC-304I17.5* | Acute bronchitis and bronchiolitis | 4.22106E-13 | NA |
| *CTC-304I17.5* | Osteopenia or other disorder of bone and cartilage | 2.07372E-12 | NA |
| *GOLGA6B* | Essential hypertension | 1.51053E-11 | NA |
| *GOLGA6B* | Hypertension | 4.15418E-12 | NA |
| *GOLGA6B* | Atherosclerosis | 1.97951E-13 | NA |
| *LINC00563* | Cancer of urinary organs (incl. kidney and bladder) | 8.07359E-11 | NA |
| *AC109826.1* | Respiratory failure | 5.06708E-13 | NA |
| *CTC-304I17.5* | Intestinal infection | 4.35733E-12 | NA |
| *OBP2B* | Inflammation of the eye | 4.00499E-14 | NA |
| *TGM3* | Frequency of urination and polyuria | 2.92387E-23 | NA |
| *NPHS2* | Nonspecific chest pain | 3.30326E-17 | NA |
| *LINC01001* | Disturbance of skin sensation | 1.27903E-12 | NA |
| *CTC-304I17.5* | Symptoms and disorders of the joints | 3.86918E-13 | NA |
| *HLA-DRB5* | Type 1 diabetes | 1.56428E-11 | NA |
| *OBP2B* | Osteoarthritis; localized | 5.12247E-11 | NA |
| *GOLGA6B* | End stage renal disease | 4.90752E-13 | NA |
| *ASH1L* | Acute pulmonary heart disease | 9.06293E-13 | NA |
| *RP11-552E20.4* | Nonspecific chest pain | 1.37427E-16 | NA |
| *ASH1L* | Pulmonary embolism and infarction, acute | 7.32764E-13 | NA |
| *CTC-304I17.5* | Pleurisy; pleural effusion | 8.54197E-11 | NA |
| *CTC-304I17.5* | Dysphagia | 1.45612E-11 | NA |
| *CTC-304I17.5* | Viral hepatitis C | 1.25681E-20 | NA |
| *GOLGA6B* | Dysphagia | 8.64035E-13 | NA |
| *GOLGA6B* | Urinary tract infection | 4.05532E-13 | NA |
| *RP11-49P4.7* | Cardiomegaly | 1.32037E-18 | NA |
| *HIST1H1B* | Occlusion and stenosis of precerebral arteries | 6.60188E-12 | NA |
| *OBP2B* | Insomnia | 2.22448E-26 | NA |
| *CTD-3116E22.8* | Decubitus ulcer | 1.54019E-11 | 0.8058 |
| *ZMYM6* | Other arthropathies | 1.06203E-13 | NA |
| *TTK* | Vitamin deficiency | 6.04353E-12 | NA |
| *TOMM40* | Disorders of lipoid metabolism | 2.05239E-13 | NA |
| *TOMM40* | Hyperlipidemia | 7.3749E-13 | NA |
| *TRIM41* | Abnormal coagulation profile | 5.17872E-15 | NA |
| *BATF3* | Other retinal disorders | 6.91858E-15 | NA |
| *RACK1* | Disorders of refraction and accommodation; blindness and low vision | 5.59766E-11 | NA |
| *RBM27* | Other disorders of eyelids | 9.99326E-12 | NA |
| *PDCD7* | Hypertensive chronic kidney disease | 1.36818E-23 | NA |
| *MED15* | Primary pulmonary hypertension | 4.8913E-14 | 0.1798 |
| *DRD2* | Cerebrovascular disease | 4.73135E-14 | NA |
| *MPLKIP* | Varicose veins | 2.53219E-13 | NA |
| *LINC00989* | Pneumonia | 6.47648E-17 | NA |
| *GATA6-AS1* | Intestinal infection | 8.61626E-12 | NA |
| *OBP2B* | Septicemia | 1.39597E-14 | NA |
| *CAMK2N2* | Viral hepatitis | 3.85433E-14 | NA |
| *CTC-304I17.5* | Viral hepatitis | 2.61493E-25 | NA |
| *EIF2S2* | Viral hepatitis C | 1.67798E-11 | NA |
| *MPHOSPH9* | Viral infection | 1.093E-12 | NA |
| *SLC12A7* | Viral infection | 1.44698E-11 | NA |
| *RFESD* | Viral infection | 3.05695E-11 | NA |
| *LINC01785* | Viral infection | 3.50764E-12 | NA |
| *RP11-173P15.9* | Viral infection | 1.35953E-13 | NA |
| *GOLGA6B* | Postoperative infection | 1.2689E-10 | NA |
| *GOLGA6B* | Infection/inflammation of internal prosthetic device; implant; and graft | 5.42177E-47 | NA |
| *S100A6* | Candidiasis | 7.88203E-11 | NA |
| *RP11-167N5.5* | Candidiasis | 3.43467E-11 | NA |
| *MCHR2* | Cancer of prostate | 1.66647E-11 | NA |
| *GATA6-AS1* | Cancer of urinary organs (incl. kidney and bladder) | 1.19858E-10 | NA |
| *ALDH3A1* | Cancer, suspected or other | 6.54364E-12 | NA |
| *CCR9* | Cancer, suspected or other | 1.69277E-11 | NA |
| *LINC02009* | Non-Hodgkins lymphoma | 2.70152E-11 | NA |
| *PDCD7* | Benign neoplasm of colon | 6.21749E-11 | NA |
| *STRA6* | Benign neoplasm of skin | 5.60431E-11 | NA |
| *AEBP1* | Diabetes mellitus | 1.54402E-14 | NA |
| *GOLGA6B* | Diabetes mellitus | 1.13119E-32 | NA |
| *GOLGA6B* | Type 1 diabetes | 1.26104E-17 | NA |
| *LINC02009* | Type 1 diabetes | 6.71711E-13 | NA |
| *AEBP1* | Type 2 diabetes | 5.78296E-13 | NA |
| *UNC119* | Type 2 diabetes | 2.72991E-12 | NA |
| *GOLGA6B* | Type 2 diabetes | 1.41922E-33 | NA |
| *SLC12A7* | Type 2 diabetes with renal manifestations | 6.60536E-11 | NA |
| *HIST1H1B* | Type 2 diabetes with renal manifestations | 7.80328E-19 | NA |
| *SPA17* | Type 2 diabetes with neurological manifestations | 6.75785E-13 | NA |
| *UBA3* | Abnormal glucose | 7.30887E-12 | NA |
| *SLC6A19* | Abnormal glucose | 8.95993E-11 | NA |
| *SLC12A7* | Polyneuropathy in diabetes | 6.8043E-13 | NA |
| *GOLGA6B* | Polyneuropathy in diabetes | 1.37503E-38 | NA |
| *HSPB11* | severe protein-calorie malnutrition | 1.15026E-10 | NA |
| *NPHS2* | severe protein-calorie malnutrition | 9.06715E-14 | NA |
| *TTK* | Vitamin D deficiency | 1.19966E-10 | NA |
| *RP11-552E20.4* | Vitamin D deficiency | 2.30406E-12 | NA |
| *CTC-338M12.9* | Vitamin D deficiency | 7.8619E-12 | NA |
| *TMEM252* | Proteinuria | 4.12432E-17 | NA |
| *ST6GAL1* | Disorders of protein plasma/amino-acid transport and metabolism | 1.60262E-12 | NA |
| *DNAJC10* | Disorders of protein plasma/amino-acid transport and metabolism | 1.04785E-12 | NA |
| *PDCD7* | Mixed hyperlipidemia | 1.34661E-15 | NA |
| *MLLT11* | Mixed hyperlipidemia | 3.96183E-13 | NA |
| *GATA6-AS1* | Mixed hyperlipidemia | 5.53584E-12 | NA |
| *PPOX* | Gout and other crystal arthropathies | 9.16373E-12 | NA |
| *CTC-304I17.5* | Gout and other crystal arthropathies | 2.59087E-14 | NA |
| *ITGA11* | Gout | 1.3097E-11 | NA |
| *PPOX* | Gout | 9.44627E-12 | NA |
| *CTC-304I17.5* | Gout | 1.53411E-13 | NA |
| *EIF3K* | Disorders of mineral metabolism | 9.4621E-12 | NA |
| *NPHS2* | Disorders of calcium/phosphorus metabolism | 2.40072E-12 | NA |
| *OBP2B* | Disorders of fluid, electrolyte, and acid-base balance | 5.89278E-22 | NA |
| *CTC-304I17.5* | Disorders of fluid, electrolyte, and acid-base balance | 1.78349E-28 | NA |
| *OBP2B* | Electrolyte imbalance | 1.36418E-17 | NA |
| *CTC-304I17.5* | Electrolyte imbalance | 3.22951E-20 | NA |
| *LIPA* | Hyposmolality and/or hyponatremia | 2.34053E-11 | NA |
| *CTC-304I17.5* | Hyposmolality and/or hyponatremia | 4.30071E-12 | NA |
| *CTC-304I17.5* | Hypopotassemia | 3.10613E-31 | NA |
| *CSDE1* | Hypovolemia | 1.31581E-10 | NA |
| *METTL5* | Hypovolemia | 6.66851E-13 | NA |
| *OBP2B* | Hypovolemia | 8.1791E-18 | NA |
| *CTC-304I17.5* | Hypovolemia | 1.21092E-28 | NA |
| *GOLGA6B* | Fluid overload | 4.72468E-12 | NA |
| *RP11-552E20.4* | Fluid overload | 4.08504E-20 | NA |
| *KAT6A* | Dysmetabolic syndrome X | 1.10234E-10 | NA |
| *CD8A* | Dysmetabolic syndrome X | 2.15924E-11 | NA |
| *CDC26* | Overweight, obesity and other hyperalimentation | 9.40505E-11 | NA |
| *MLLT11* | Overweight, obesity and other hyperalimentation | 2.63794E-13 | NA |
| *MLLT11* | Obesity | 4.686E-11 | NA |
| *LINC00563* | Obesity | 8.68468E-12 | NA |
| *PDCD7* | Morbid obesity | 1.42426E-24 | NA |
| *MAP1B* | Morbid obesity | 1.67547E-13 | NA |
| *PRKCSH* | Iron deficiency anemias | 3.87902E-11 | NA |
| *DUT* | Iron deficiency anemias, unspecified or not due to blood loss | 1.04945E-10 | NA |
| *PDCD7* | Acute posthemorrhagic anemia | 9.71237E-14 | NA |
| *AARS2* | Coagulation defects | 1.05707E-11 | NA |
| *CTC-304I17.5* | Coagulation defects | 3.11633E-16 | NA |
| *CTC-471J1.9* | Coagulation defects | 2.92277E-16 | NA |
| *ATRAID* | Purpura and other hemorrhagic conditions | 1.60515E-11 | NA |
| *FREM2* | Purpura and other hemorrhagic conditions | 6.15345E-11 | NA |
| *CTC-304I17.5* | Purpura and other hemorrhagic conditions | 3.79623E-13 | NA |
| *GATA6-AS1* | Purpura and other hemorrhagic conditions | 9.11132E-12 | NA |
| *ATRAID* | Thrombocytopenia | 1.00003E-11 | NA |
| *PDCD7* | Decreased white blood cell count | 1.1119E-13 | NA |
| *SPINT1* | Decreased white blood cell count | 1.80341E-13 | NA |
| *NELFA* | Lymphadenitis | 1.80998E-11 | NA |
| *RP4-781K5.4* | Lymphadenitis | 1.00984E-11 | NA |
| *GOLGA6B* | Neurological disorders | 1.02575E-11 | NA |
| *NPHS2* | Altered mental status | 2.13165E-11 | NA |
| *CTC-471J1.9* | Altered mental status | 4.75568E-15 | NA |
| *OBP2B* | Sleep disorders | 1.15638E-16 | NA |
| *CAND1* | Migraine | 7.90594E-12 | NA |
| *RP11-49P4.7* | Epilepsy, recurrent seizures, convulsions | 7.68217E-12 | NA |
| *STAT5A* | Abnormality of gait | 1.18227E-11 | NA |
| *LINC01833* | Abnormality of gait | 8.92606E-13 | NA |
| *VAX1* | Hereditary and idiopathic peripheral neuropathy | 8.79195E-13 | NA |
| *P3H3* | Degeneration of macula and posterior pole of retina | 5.86354E-12 | NA |
| *GATA6-AS1* | Glaucoma | 3.39247E-12 | NA |
| *SLC39A4* | Cataract | 2.19034E-11 | NA |
| *COX8A* | Senile cataract | 1.58864E-11 | NA |
| *LINC01001* | Visual disturbances | 1.11628E-10 | NA |
| *RP11-223C24.1* | Visual disturbances | 1.20613E-12 | NA |
| *GATA6-AS1* | Visual disturbances | 1.58497E-12 | NA |
| *CSDE1* | Inflammation of the eye | 9.43757E-12 | NA |
| *LINC01001* | Inflammation of the eye | 1.75384E-12 | NA |
| *GBE1* | Disorders of vitreous body | 9.92834E-11 | NA |
| *CDH5* | Impacted cerumen | 4.09742E-17 | NA |
| *CAND1* | Hearing loss | 9.54902E-11 | NA |
| *RP11-662M24.2* | Hearing loss | 1.39906E-12 | NA |
| *GATA6-AS1* | Hearing loss | 2.81505E-19 | NA |
| *PDCD7* | Rheumatic disease of the heart valves | 1.40765E-13 | NA |
| *MED15* | Rheumatic disease of the heart valves | 3.04022E-12 | NA |
| *BLK* | Rheumatic disease of the heart valves | 7.68912E-12 | NA |
| *MED15* | Disease of tricuspid valve | 8.00147E-15 | NA |
| *GATA6-AS1* | Disease of tricuspid valve | 1.0764E-13 | NA |
| *ZNF836* | Heart valve disorders | 5.74416E-11 | NA |
| *HBZ* | Nonrheumatic aortic valve disorders | 4.32608E-12 | NA |
| *ZNF836* | Nonrheumatic aortic valve disorders | 3.31872E-11 | NA |
| *PDCD7* | Hypertensive heart and/or renal disease | 3.06723E-17 | NA |
| *P2RX3* | Hypertensive heart disease | 3.62361E-11 | NA |
| *PPP1R3A* | Hypertensive heart disease | 4.53587E-12 | NA |
| *LIPE* | Hypertensive chronic kidney disease | 8.62661E-11 | NA |
| *CTC-304I17.5* | Hypertensive chronic kidney disease | 2.61131E-11 | NA |
| *ORC6* | Ischemic Heart Disease | 4.73601E-14 | NA |
| *PPP1R3A* | Ischemic Heart Disease | 1.69721E-13 | NA |
| *CTC-304I17.5* | Ischemic Heart Disease | 7.79435E-13 | NA |
| *CTC-304I17.5* | Myocardial infarction | 7.0399E-14 | NA |
| *KBTBD3* | Angina pectoris | 5.05824E-11 | NA |
| *CSDE1* | Coronary atherosclerosis | 4.57199E-13 | NA |
| *ORC6* | Coronary atherosclerosis | 9.17014E-12 | NA |
| *PPP1R3A* | Coronary atherosclerosis | 3.7291E-13 | NA |
| *RFESD* | Coronary atherosclerosis | 3.03901E-12 | NA |
| *CTC-304I17.5* | Coronary atherosclerosis | 8.84706E-14 | NA |
| *HBZ* | Other chronic ischemic heart disease, unspecified | 9.06371E-15 | NA |
| *FREM2* | Other chronic ischemic heart disease, unspecified | 1.38399E-11 | NA |
| *LINC00284* | Other chronic ischemic heart disease, unspecified | 8.68454E-12 | NA |
| *CTC-471J1.9* | Other chronic ischemic heart disease, unspecified | 2.2297E-11 | NA |
| *ASH1L* | Pulmonary heart disease | 3.92201E-11 | NA |
| *CTC-304I17.5* | Pulmonary heart disease | 3.54605E-14 | NA |
| *TMEM132A* | Acute pulmonary heart disease | 8.85632E-11 | NA |
| *CTC-304I17.5* | Acute pulmonary heart disease | 1.04512E-16 | NA |
| *CTC-304I17.5* | Pulmonary embolism and infarction, acute | 1.65494E-17 | NA |
| *MED15* | Chronic pulmonary heart disease | 7.88003E-12 | NA |
| *RP11-320M2.1* | Primary pulmonary hypertension | 7.82067E-13 | NA |
| *RP11-552E20.4* | Cardiomegaly | 4.91399E-12 | NA |
| *GOLGA6B* | Nonspecific chest pain | 2.77311E-16 | NA |
| *CSDE1* | Precordial pain | 1.56737E-14 | NA |
| *CTC-304I17.5* | Precordial pain | 3.60417E-14 | NA |
| *CTC-471J1.9* | Carditis | 7.54822E-13 | NA |
| *SLC39A4* | Cardiomyopathy | 1.81381E-11 | NA |
| *MORC1* | Secondary/extrinsic cardiomyopathies | 1.39735E-11 | NA |
| *NPHS2* | Secondary/extrinsic cardiomyopathies | 8.56891E-12 | NA |
| *CTC-304I17.5* | Secondary/extrinsic cardiomyopathies | 4.73432E-11 | NA |
| *OBP2B* | Cardiac conduction disorders | 2.66974E-15 | NA |
| *GOLGA6B* | Cardiac conduction disorders | 2.66847E-12 | NA |
| *LINC01122* | Cardiac conduction disorders | 1.74504E-11 | NA |
| *CTC-304I17.5* | Cardiac conduction disorders | 4.86704E-11 | NA |
| *TRIM41* | Bundle branch block | 6.20479E-11 | NA |
| *BSDC1* | Bundle branch block | 3.61156E-11 | NA |
| *KDM4E* | Right bundle branch block | 8.633E-13 | NA |
| *BSDC1* | Left bundle branch block | 1.76017E-13 | NA |
| *CTC-304I17.5* | Cardiac dysrhythmias | 8.13742E-12 | NA |
| *GOLGA6B* | Paroxysmal tachycardia, unspecified | 3.27803E-11 | NA |
| *SERPINH1* | Paroxysmal ventricular tachycardia | 1.06126E-11 | NA |
| *OBP2B* | Paroxysmal ventricular tachycardia | 2.09959E-11 | NA |
| *CTC-304I17.5* | Paroxysmal ventricular tachycardia | 2.06691E-13 | NA |
| *CDH5* | Atrial flutter | 1.22018E-13 | NA |
| *CTC-304I17.5* | Cardiac arrest and ventricular fibrillation | 1.94754E-11 | NA |
| *TMEM252* | Arrhythmia (cardiac) NOS | 2.90219E-13 | NA |
| *HIST1H1B* | Palpitations | 4.2665E-13 | NA |
| *GOLGA6B* | Congestive heart failure; nonhypertensive | 5.86967E-13 | NA |
| *RP11-202G18.1* | Congestive heart failure; nonhypertensive | 6.60526E-14 | NA |
| *CTC-304I17.5* | Congestive heart failure; nonhypertensive | 5.68156E-18 | NA |
| *CTC-471J1.9* | Congestive heart failure; nonhypertensive | 1.50839E-16 | NA |
| *RP11-202G18.1* | Congestive heart failure (CHF) NOS | 1.4973E-11 | NA |
| *CTC-304I17.5* | Congestive heart failure (CHF) NOS | 7.68626E-13 | NA |
| *CTC-471J1.9* | Congestive heart failure (CHF) NOS | 2.76102E-18 | NA |
| *FREM2* | Heart failure NOS | 1.4718E-14 | NA |
| *CTC-304I17.5* | Heart failure NOS | 3.17806E-15 | NA |
| *CTC-471J1.9* | Heart failure with reduced EF [Systolic or combined heart failure] | 2.03032E-12 | NA |
| *RP11-675F6.4* | Heart failure with preserved EF [Diastolic heart failure] | 6.33822E-11 | NA |
| *GATA6-AS1* | Heart failure with preserved EF [Diastolic heart failure] | 3.54358E-14 | NA |
| *GATA6-AS1* | Ill-defined descriptions and complications of heart disease | 1.66321E-13 | NA |
| *CDH5* | Heart transplant/surgery | 2.09513E-17 | NA |
| *GOLGA6B* | Abnormal function study of cardiovascular system | 4.23168E-15 | NA |
| *GPATCH8* | Symptoms involving cardiovascular system | 7.87914E-16 | NA |
| *NPHS2* | Cerebrovascular disease | 1.1115E-15 | NA |
| *FREM2* | Occlusion and stenosis of precerebral arteries | 7.08756E-11 | NA |
| *GOLGA6B* | Occlusion and stenosis of precerebral arteries | 3.66431E-11 | NA |
| *BSDC1* | Occlusion of cerebral arteries | 2.31911E-11 | NA |
| *RFESD* | Occlusion of cerebral arteries | 1.05158E-10 | NA |
| *BSDC1* | Cerebral artery occlusion, with cerebral infarction | 1.08171E-11 | NA |
| *RFESD* | Cerebral artery occlusion, with cerebral infarction | 1.33786E-12 | NA |
| *NPHS2* | Cerebral ischemia | 4.52475E-11 | NA |
| *C2CD4A* | Cerebral ischemia | 2.98289E-13 | NA |
| *GOLGA6B* | Acute, but ill-defined cerebrovascular disease | 1.52938E-18 | NA |
| *ZNF836* | Atherosclerosis | 1.01047E-11 | NA |
| *CTC-471J1.9* | Atherosclerosis | 7.12277E-11 | NA |
| *LINC00563* | Aortic aneurysm | 4.23954E-13 | NA |
| *LINC00563* | Abdominal aortic aneurysm | 1.74458E-11 | NA |
| *ELFN1* | Peripheral vascular disease | 1.38753E-11 | NA |
| *ELFN1* | Peripheral vascular disease, unspecified | 6.47852E-11 | NA |
| *ZNF688* | Peripheral vascular disease, unspecified | 5.63146E-11 | NA |
| *CTC-471J1.9* | Other venous embolism and thrombosis | 5.56866E-11 | NA |
| *CTC-304I17.5* | Deep vein thrombosis [DVT] | 2.06127E-12 | NA |
| *NPHS2* | Varicose veins | 1.33E-12 | NA |
| *MORC2* | Varicose veins | 9.63626E-11 | NA |
| *GPRC5C* | Chronic venous insufficiency [CVI] | 1.44544E-12 | NA |
| *NPHS2* | Acute sinusitis | 4.56816E-26 | NA |
| *HOXD10* | Acute sinusitis | 4.24247E-13 | NA |
| *MPHOSPH8* | Acute sinusitis | 1.80246E-12 | NA |
| *LIPA* | Acute upper respiratory infections of multiple or unspecified sites | 1.10261E-10 | NA |
| *HOXD10* | Acute upper respiratory infections of multiple or unspecified sites | 6.24909E-11 | NA |
| *CPN1* | Chronic pharyngitis and nasopharyngitis | 5.82915E-14 | NA |
| *CTC-304I17.5* | Diseases of the larynx and vocal cords | 4.02696E-16 | NA |
| *GATA6-AS1* | Diseases of the larynx and vocal cords | 3.95153E-13 | NA |
| *ZMYND10* | Chronic sinusitis | 1.06915E-11 | NA |
| *TMEM115* | Chronic sinusitis | 5.14607E-12 | NA |
| *GOLGA6B* | Epistaxis or throat hemorrhage | 2.75646E-15 | NA |
| *PDCD7* | Pneumonia | 2.28762E-12 | NA |
| *RP11-49P4.7* | Pneumonia | 1.61712E-15 | NA |
| *UNC93A* | Acute bronchitis and bronchiolitis | 1.09535E-10 | NA |
| *GOLGA6B* | Chronic airway obstruction | 1.10577E-19 | NA |
| *RP11-552E20.4* | Chronic airway obstruction | 3.56439E-15 | NA |
| *GPRC5C* | Postinflammatory pulmonary fibrosis | 5.83962E-17 | NA |
| *PDCD7* | Pleurisy; pleural effusion | 5.91297E-27 | NA |
| *QRFPR* | Pleurisy; pleural effusion | 1.55784E-12 | NA |
| *RP11-552E20.4* | Pulmonary collapse; interstitial and compensatory emphysema | 4.48853E-15 | NA |
| *CTC-304I17.5* | Pulmonary collapse; interstitial and compensatory emphysema | 1.8269E-12 | NA |
| *PCDH17* | Respiratory abnormalities | 7.83837E-12 | NA |
| *LINC01001* | Respiratory abnormalities | 4.0292E-14 | NA |
| *SLC39A4* | Abnormal findings examination of lungs | 1.07293E-11 | NA |
| *CTC-471J1.9* | Abnormal findings examination of lungs | 3.6287E-12 | NA |
| *GATA6-AS1* | Solitary pulmonary nodule | 4.65883E-13 | NA |
| *ATP4A* | Other diseases of respiratory system, not elsewhere classified | 8.89462E-14 | NA |
| *FREM2* | Other diseases of respiratory system, not elsewhere classified | 1.23966E-12 | NA |
| *ATP4A* | Symptoms involving respiratory system and other chest symptoms | 1.43548E-11 | NA |
| *SLC12A7* | Symptoms involving respiratory system and other chest symptoms | 7.93842E-15 | NA |
| *FREM2* | Symptoms involving respiratory system and other chest symptoms | 9.88555E-12 | NA |
| *NPHS2* | Diseases of esophagus | 5.44424E-13 | NA |
| *GOLGA6B* | Diseases of esophagus | 9.07675E-17 | NA |
| *NPHS2* | Esophagitis, GERD and related diseases | 6.92005E-15 | NA |
| *GOLGA6B* | Esophagitis, GERD and related diseases | 4.6298E-17 | NA |
| *NPHS2* | GERD | 1.25932E-14 | NA |
| *GOLGA6B* | GERD | 2.0102E-16 | NA |
| *MED23* | Dysphagia | 2.11203E-11 | NA |
| *HIST1H1B* | Disorders of function of stomach | 1.08548E-12 | NA |
| *RP11-324I22.2* | Disorders of function of stomach | 1.78048E-11 | NA |
| *CTC-304I17.5* | Disorders of function of stomach | 1.0715E-13 | NA |
| *NPHS2* | Abdominal hernia | 1.42316E-14 | NA |
| *GATA6-AS1* | Abdominal hernia | 3.54679E-15 | NA |
| *GATA6-AS1* | Inguinal hernia | 1.51166E-14 | NA |
| *SAMD9L* | Incisional hernia | 1.35161E-10 | NA |
| *CTC-471J1.9* | Intestinal obstruction without mention of hernia | 6.81669E-11 | NA |
| *CDH5* | Symptoms involving digestive system | 5.1299E-17 | NA |
| *KAT6A* | Diverticulosis and diverticulitis | 1.50044E-11 | NA |
| *EIF3K* | Diverticulosis and diverticulitis | 4.41934E-12 | NA |
| *EIF3K* | Diverticulosis | 4.19955E-16 | NA |
| *MED23* | Functional digestive disorders | 1.13085E-13 | NA |
| *COX8A* | Chronic liver disease and cirrhosis | 5.48929E-11 | NA |
| *METTL21C* | Other chronic nonalcoholic liver disease | 1.16434E-10 | NA |
| *COX8A* | Cirrhosis of liver without mention of alcohol | 4.86231E-11 | NA |
| *CTC-471J1.9* | Liver abscess and sequelae of chronic liver disease | 2.55581E-12 | NA |
| *CTC-304I17.5* | Ascites (non malignant) | 1.28288E-23 | NA |
| *SLC12A7* | Nonspecific elevation of levels of transaminase or lactic acid dehydrogenase [LDH] | 3.99286E-11 | NA |
| *RP11-320M2.1* | Nonspecific elevation of levels of transaminase or lactic acid dehydrogenase [LDH] | 2.19048E-14 | NA |
| *SLC39A4* | Diseases of pancreas | 1.66993E-12 | NA |
| *CTC-304I17.5* | Diseases of pancreas | 2.3132E-19 | NA |
| *PDCD7* | Renal failure | 2.77682E-15 | NA |
| *GOLGA6B* | Renal failure | 7.19557E-15 | NA |
| *KIF5C* | Acute renal failure | 5.61125E-12 | NA |
| *OBP2B* | Acute renal failure | 5.76981E-11 | NA |
| *PRR34* | Acute renal failure | 2.65852E-11 | NA |
| *CTC-304I17.5* | Acute renal failure | 2.39505E-16 | NA |
| *INHBA* | Renal failure NOS | 3.38698E-11 | NA |
| *HIST1H1B* | Renal failure NOS | 6.01066E-11 | NA |
| *CTC-304I17.5* | Renal failure NOS | 1.38151E-12 | NA |
| *PDCD7* | Chronic renal failure [CKD] | 1.99722E-13 | NA |
| *SLC12A7* | Chronic renal failure [CKD] | 4.89442E-11 | NA |
| *HIST1H1B* | Chronic renal failure [CKD] | 2.52664E-13 | NA |
| *GOLGA6B* | Chronic renal failure [CKD] | 9.3864E-12 | NA |
| *SLC12A7* | End stage renal disease | 1.71429E-11 | NA |
| *HIST1H1B* | End stage renal disease | 1.22673E-14 | NA |
| *NDUFA7* | Chronic kidney disease, Stage I or II | 1.12543E-10 | NA |
| *OBP2B* | Disorders resulting from impaired renal function | 2.41431E-11 | NA |
| *AC109826.1* | Disorders resulting from impaired renal function | 8.97702E-12 | NA |
| *PDCD7* | Secondary hyperparathyroidism (of renal origin) | 8.29923E-11 | NA |
| *SLC12A7* | Secondary hyperparathyroidism (of renal origin) | 1.9293E-11 | NA |
| *PDCD7* | Urinary tract infection | 7.55036E-15 | NA |
| *RP11-167N5.5* | Hematuria | 1.04394E-11 | NA |
| *RP11-439E19.9* | Urinary calculus | 3.46396E-15 | NA |
| *SCNN1A* | Calculus of kidney | 1.88587E-11 | NA |
| *RP11-439E19.9* | Calculus of kidney | 1.69651E-15 | NA |
| *TGM5* | Retention of urine | 1.49663E-12 | NA |
| *EMC2* | Urinary incontinence | 3.70237E-12 | NA |
| *MBOAT2* | Urinary incontinence | 5.07969E-12 | NA |
| *HIST1H1B* | Urinary incontinence | 4.17904E-13 | NA |
| *RP11-307C19.2* | Frequency of urination and polyuria | 1.03344E-11 | NA |
| *DHRS3* | Hyperplasia of prostate | 1.51374E-11 | NA |
| *OLFM2* | Erectile dysfunction [ED] | 8.3818E-12 | NA |
| *CTC-471J1.9* | Superficial cellulitis and abscess | 1.05031E-14 | NA |
| *PDCD1* | Rash and other nonspecific skin eruption | 1.02667E-10 | NA |
| *FREM2* | Disturbance of skin sensation | 2.27223E-13 | NA |
| *SHISA9* | Disturbance of skin sensation | 3.65095E-11 | NA |
| *ZNF790* | Dyschromia and Vitiligo | 9.00945E-11 | NA |
| *GOLGA8R* | Erythematous conditions | 6.32333E-11 | NA |
| *PDCD7* | Scar conditions and fibrosis of skin | 1.40321E-10 | NA |
| *COX8A* | Scar conditions and fibrosis of skin | 8.93609E-11 | NA |
| *CTC-304I17.5* | Scar conditions and fibrosis of skin | 8.17297E-12 | NA |
| *SLC6A19* | Diseases of sebaceous glands | 8.48457E-13 | NA |
| *CDH5* | Chronic ulcer of skin | 6.24278E-11 | NA |
| *ZNF836* | Chronic ulcer of skin | 3.08371E-11 | NA |
| *CTC-471J1.9* | Chronic ulcer of skin | 6.95346E-13 | NA |
| *TGM3* | Rheumatoid arthritis and other inflammatory polyarthropathies | 5.01088E-12 | NA |
| *ADCY5* | Rheumatoid arthritis and other inflammatory polyarthropathies | 9.08933E-12 | NA |
| *LINC01001* | Rheumatoid arthritis and other inflammatory polyarthropathies | 6.55042E-13 | NA |
| *ADCY5* | Rheumatoid arthritis | 1.13875E-12 | NA |
| *MLLT11* | Spinal stenosis | 3.26565E-14 | NA |
| *CAND1* | Spinal stenosis of lumbar region | 8.77316E-14 | NA |
| *MLLT11* | Spinal stenosis of lumbar region | 1.20193E-10 | NA |
| *MLLT11* | Spondylosis and allied disorders | 3.41071E-13 | NA |
| *PPOX* | Spondylosis without myelopathy | 5.81088E-12 | NA |
| *MLLT11* | Spondylosis without myelopathy | 7.8188E-12 | NA |
| *GOLGA6B* | Spondylosis without myelopathy | 1.13328E-11 | NA |
| *EPB41L4A* | Degeneration of intervertebral disc | 2.34623E-11 | NA |
| *GOLGA6B* | Degeneration of intervertebral disc | 2.52798E-12 | NA |
| *LINC01001* | Peripheral enthesopathies and allied syndromes | 1.82316E-13 | NA |
| *UNC93A* | Enthesopathy | 6.10293E-11 | NA |
| *LINC01001* | Enthesopathy | 1.01067E-11 | NA |
| *ZMYND10* | Osteoarthrosis | 3.2332E-11 | NA |
| *PON2* | Osteoarthrosis | 3.75448E-14 | NA |
| *TMEM115* | Osteoarthrosis | 4.41364E-13 | NA |
| *KIF5C* | Osteoarthrosis | 5.16828E-11 | NA |
| *OBP2B* | Osteoarthrosis | 9.07289E-13 | NA |
| *GFRA4* | Osteoarthritis; localized | 2.92154E-11 | NA |
| *AC114730.2* | Osteoarthritis; localized | 9.2815E-11 | NA |
| *CTC-304I17.5* | Osteoarthrosis, localized, primary | 2.01188E-13 | NA |
| *HBZ* | Osteoarthrosis, generalized | 9.80334E-11 | NA |
| *CRMP1* | Osteoarthrosis NOS | 1.48069E-11 | NA |
| *PON2* | Osteoarthrosis NOS | 6.97234E-11 | NA |
| *EIF2S2* | Osteoarthrosis NOS | 2.23864E-12 | NA |
| *OBP2B* | Osteoarthrosis NOS | 3.36397E-12 | NA |
| *RP5-1125A11.6* | Osteoarthrosis NOS | 1.96465E-13 | NA |
| *OBP2B* | Symptoms and disorders of the joints | 1.54171E-13 | NA |
| *RP11-87G24.6* | Symptoms and disorders of the joints | 6.5801E-13 | NA |
| *OBP2B* | Osteoporosis, osteopenia and pathological fracture | 6.6297E-11 | NA |
| *GOLGA6B* | Osteoporosis | 8.36345E-14 | NA |
| *GOLGA6B* | Osteoporosis NOS | 2.38598E-16 | NA |
| *PRKCSH* | Senile osteoporosis | 3.72913E-11 | NA |
| *CARD11* | Senile osteoporosis | 5.65798E-11 | NA |
| *NPHS2* | Osteopenia or other disorder of bone and cartilage | 1.38555E-13 | NA |
| *HIST1H1B* | Osteopenia or other disorder of bone and cartilage | 3.9256E-12 | NA |
| *GATA6-AS1* | Cardiac congenital anomalies | 5.53482E-13 | NA |
