## Supplemental Table 5 for "Multi-ancestry gene-trait connection landscape using electronic health record (EHR) linked biobank data"

Supplementary Table 5. Statistically significant gene-disease associations that were exclusively found in the eMERGE III cross-ancestry analysis (P $\leq1.47\times{10}^{-10}$).

^a^ NS: not statistically significant

| Gene | Phecode Description | eMERGE III  Cross-ancestry | PhenomeXcan  European Ancestry |
| --- | --- | --- | --- |
|  |  | Discovery P | Validation P^a^ |
| XXbac-BPG181B23.7 | Type 1 diabetes | 4.17E-12 | 8.94E-24 |
| ATP6V1G2 | Type 1 diabetes | 6.52E-20 | 1.43E-22 |
| HLA-B | Type 1 diabetes | 9.57E-13 | 1.64E-20 |
| MICB | Type 1 diabetes | 1.46E-10 | 4.12E-17 |
| TAP1 | Type 1 diabetes | 1.18E-14 | 3.46E-15 |
| AGPAT1 | Rheumatoid arthritis and other inflammatory polyarthropathies | 4.33E-17 | 1.35E-13 |
| RP11-325L7.2 | Atrial fibrillation and flutter | 7.03E-11 | 3.14E-11 |
| DXO | Type 1 diabetes | 1.87E-17 | 7.04E-11 |
| MICA | Type 1 diabetes | 4.72E-11 | 9.51E-11 |
| FGF7 | Nontoxic multinodular goiter | 1.27E-10 | NS^a^ |
| XPA | Hypothyroidism | 2.84E-20 | NS |
| RP11-356N1.2 | Hypothyroidism | 1.65E-12 | NS |
| NOTCH4 | Hypothyroidism | 1.28E-10 | NS |
| XPA | Hypothyroidism NOS | 1.73E-21 | NS |
| RP11-356N1.2 | Hypothyroidism NOS | 1.33E-12 | NS |
| ATF6B | Type 1 diabetes | 1.44E-29 | NS |
| CCHCR1 | Type 1 diabetes | 2.44E-11 | NS |
| NELFE | Type 1 diabetes | 2.12E-15 | NS |
| HLA-DRB1 | Type 2 diabetes | 3.16E-11 | NS |
| HLA-DQA1 | Type 2 diabetes | 4.09E-15 | NS |
| RP11-395N3.2 | Type 2 diabetes | 1.33E-10 | NS |
| TCF7L2 | Type 2 diabetes with renal manifestations | 7.79E-17 | NS |
| HLA-DQB1 | Polyneuropathy in diabetes | 8.40E-11 | NS |
| HLA-DQB2 | Polyneuropathy in diabetes | 2.55E-12 | NS |
| SYPL2 | Disorders of lipoid metabolism | 2.71E-18 | NS |
| SORT1 | Disorders of lipoid metabolism | 7.63E-16 | NS |
| SYPL2 | Hyperlipidemia | 1.65E-18 | NS |
| SORT1 | Hyperlipidemia | 5.82E-16 | NS |
| ATXN7L2 | Hypercholesterolemia | 1.07E-15 | NS |
| SORT1 | Hypercholesterolemia | 4.29E-14 | NS |
| RPGRIP1L | Overweight, obesity and other hyperalimentation | 1.78E-12 | NS |
| GSDMA | Elevated white blood cell count | 2.03E-12 | NS |
| CSF3 | Elevated white blood cell count | 2.93E-11 | NS |
| RP11-387H17.6 | Elevated white blood cell count | 8.19E-11 | NS |
| PHACTR1 | Ischemic Heart Disease | 1.27E-10 | NS |
| RP11-325L7.2 | Atrial fibrillation | 5.87E-11 | NS |
| HLA-DQA2 | Atherosclerosis | 1.17E-10 | NS |
| HLA-B | Atherosclerosis | 4.30E-15 | NS |
| HLA-DQA2 | Atherosclerosis of the extremities | 2.37E-11 | NS |
| C6orf136 | Atherosclerosis of the extremities | 1.30E-10 | NS |
| TRIM26 | Atherosclerosis of native arteries of the extremities with intermittent claudication | 5.30E-12 | NS |
| CDKN2B | Abdominal aortic aneurysm | 6.33E-11 | NS |
| LPA | Peripheral vascular disease | 3.59E-11 | NS |
| HLA-B | Peripheral vascular disease, unspecified | 2.19E-11 | NS |
| BTNL2 | Arterial embolism and thrombosis | 2.08E-11 | NS |
| ATP1B1 | Other venous embolism and thrombosis | 3.20E-28 | NS |
| STKLD1 | Other venous embolism and thrombosis | 2.93E-11 | NS |
| CACFD1 | Other venous embolism and thrombosis | 7.03E-16 | NS |
| C1orf112 | Other venous embolism and thrombosis | 3.17E-17 | NS |
| ATP1B1 | Deep vein thrombosis [DVT] | 4.62E-27 | NS |
| CACFD1 | Deep vein thrombosis [DVT] | 6.89E-12 | NS |
| WNT2 | Chronic sinusitis | 4.65E-11 | NS |
| IL12RB2 | Solitary pulmonary nodule | 5.84E-11 | NS |
| SAMM50 | Chronic liver disease and cirrhosis | 3.31E-24 | NS |
| SAMM50 | Other chronic nonalcoholic liver disease | 8.78E-29 | NS |
| NOTCH4 | Chronic renal failure [CKD] | 2.57E-11 | NS |
| C4B | Chronic renal failure [CKD] | 1.06E-10 | NS |
| CYP21A2 | Chronic renal failure [CKD] | 9.56E-12 | NS |
| TNXB | Chronic renal failure [CKD] | 1.19E-11 | NS |
| ATF6B | End stage renal disease | 1.32E-10 | NS |
| HLA-DQB1 | End stage renal disease | 1.88E-11 | NS |
| TNXB | End stage renal disease | 1.32E-10 | NS |
| TAS2R43 | Chronic ulcer of skin | 4.95E-11 | NS |
| AGPAT1 | Rheumatoid arthritis | 2.14E-20 | NS |
| TMEM59L | Hypertension | 2.50E-11 | NS |
| TMEM59L | Essential hypertension | 6.95E-12 | NS |
| RP11-543C4.1 | Paroxysmal supraventricular tachycardia | 6.12E-13 | NS |
| TMEM45A | Heart failure NOS | 3.97E-11 | NS |
| RP11-543C4.1 | Diseases of the larynx and vocal cords | 1.20E-12 | NS |
| RP11-543C4.1 | Voice disturbance | 1.59E-11 | NS |
| KIF5C | Pleurisy; pleural effusion | 4.81E-11 | NS |
| CYP4F12 | Chronic Kidney Disease, Stage IV | 5.68E-14 | NS |
| TAS2R46 | Secondary hyperparathyroidism (of renal origin) | 2.58E-11 | NS |
| ATP1B2 | Osteoarthrosis NOS | 1.19E-10 | NS |
